# The impact of vaccine products, schedules and geographic regions on immunogenicity of pneumococcal vaccines in children under 2 years old: a systematic review and meta-analysis

**DOI:** 10.1101/2025.10.10.25337708

**Authors:** Xinghui Chen, Sarah Tavlian, Kylie S. Carville, Nefel Tellioglu, Violeta Spirkoska, Natalie Carvalho, David J. Price, Patricia T. Campbell, Jodie McVernon

## Abstract

**Background:** Pneumococcal conjugate vaccines (PCVs) cover only a proportion of disease-causing serotypes. In some settings, population-level introduction of PCVs has resulted in an increase in “non-vaccine” serotype incidence. Higher-valency PCVs were developed to address shifting disease-causing serotypes. We aim to systematically define the trends in vaccine immunogenicity and likely protection over time.

**Methods:** We conducted a systematic review and meta-analysis of studies published to Jan 7, 2025, reporting immunoglobulin G (IgG) responses after PCV vaccination in healthy children <2 years. Outcomes were serotype-specific IgG geometric mean concentration (GMC) and seroresponse rate. We performed random-effects meta-analyses using log-transformed GMCs and logit-transformed seroresponse rates to generate pooled estimates by vaccine product, dosing schedule, and World Health Organization (WHO) region. This study was registered with PROSPERO (CRD42024484824).

**Findings:** We included 250 articles from 138 study groups involving 244 study arms. Pooled IgG GMCs for vaccine-included serotypes post-childhood-schedule exceeded the WHO-defined protective threshold (0·35 μg/mL), but varied by serotype, lowest for serotype 3-PCV20 (0·84 μg/mL; 95% confidence interval: 0·60–1·17). Post-childhood-schedule seroresponse rates were >95% for all serotypes except serotype 3 (84–92%). A general “downward trend” in IgG GMCs was observed with the increasing vaccine valency. IgG responses increased with the number of primary doses, and were further enhanced by a booster, although magnitude varied by serotype and vaccine. IgG responses post 1-primary dose were low, whereas GMCs post 2- or 3-primary doses exceeded protective thresholds for most serotypes. Booster-containing schedules (3+1, 2+1, 1+1) generally elicited higher post-childhood-schedule IgG response than primary-only schedule (3+0). We observed substantial regional variation of post-childhood-schedule serotype-specific IgG GMCs, with highest GMCs in the Western Pacific Region.

**Interpretation:** Vaccine immunogenicity varied by serotype, vaccine product, schedule and WHO region, and should be carefully considered when evaluating potential vaccination programs.

**Funding:** PhD Scholarship; Australian Department of Foreign Affairs and Trade.

**Research in context Evidence before this study:** We implemented a targeted literature review strategy to identify prior evidence in PubMed on August 14, 2023, without time and language restrictions, using the following search strategies: ((streptococcus pneumoniae[Title]) OR (pneumococc*[Title]) AND (immun*[Title] OR antibod*[Title]) AND (review[Filter])). We identified six systematic reviews quantitatively evaluating the serotype-specific immune responses following pneumococcal conjugate vaccine (PCV) in infants. All reviews focused on PCV7, PCV10-GSK, and PCV13; none included the newer higher-valency PCVs (PCV15, PCV20) or the novel PCV10-SII. Two early reviews (2011) compared 2-dose versus 3-dose primary series in randomized clinical trials (RCTs), reporting differences in seroresponse rates — greatest for serotypes 6B and 23F — without accounting for specific vaccine products. Two reviews in 2014 further explored the impact of dosing schedules, vaccine product, and region on immune responses, but only a subset of vaccine-included serotypes was analysed. A 2020 review investigated regional variation in post-primary responses but was restricted to RCTs and did not assess post-booster immune responses. A 2023 review of head-to-head RCTs in infants comparing PCV7, PCV10-GSK and PCV13 found that 1-month post-primary serotype-specific IgG geometric mean ratios favoured PCV7 over either PCV13 or PCV10 for serotypes 4, 6B, 9V, 14, and 23F.

*Added value of this study:* PCV15 and PCV20 were developed in response to changes in patterns of disease-causing serotypes, whereas PCV10-SII was designed for developing countries with distinct serotype distribution. To date, however, no systematic review has synthesised all available PCV data and compared the immunogenicity of newer vaccines relative to earlier ones. As immunogenicity remains the cornerstone of vaccination recommendations in the absence of representative effectiveness studies, our study quantifies and compares immune responses to five widely used PCVs across alternative dosing regimens, populations, and epidemiological contexts.

*Implications of all the available evidence:* Vaccine immunogenicity varied by serotype, vaccine product, schedule and World Health Organization (WHO) region. Downward trend of serotype-specific IgG response was generally observed with increasing vaccine valency for most serotypes, indicating the importance of balancing vaccine serotype coverage and the magnitude of immune response for sustained population protection. This evidence should be considered when evaluating potential vaccination programs.

## Introduction

*Streptococcus pneumoniae* (the pneumococcus) is a multi-strain bacterial pathogen, with more than 100 immunologically distinct serotypes,^1^ that causes a substantial global disease burden.^2^ Pneumococcal conjugate vaccines (PCVs) cover only a small proportion of disease-causing serotypes. Following widespread introduction of PCVs in several populations, immunological selection resulted in shifts in the pneumococcal serotype distribution.^3^ Emergence of “non-vaccine” serotypes has been associated with replacement disease in many settings, a phenomenon termed “serotype replacement”, eroding vaccine impact.^4^

In response to changes in the distribution of disease-causing serotypes, higher-valency PCVs have been developed to target a broader spectrum of serotypes emerging as a cause of disease. Despite the transition from 7-valent PCV (PCV7) to 10-valent PCV (PCV10-GSK) and 13-valent PCV (PCV13), serotype replacement continues to occur.^5^ Recently, newer PCVs with even higher valency such as 15-valent PCV (PCV15), 20-valent PCV (PCV20) and 21-valent PCV (V116, for use in adults only) have been licensed. In 2019, a novel 10-valent PCV (PCV10-SII), designed to protect against serotypes causing most diseases in low- and middle-income countries, received prequalification status by the World Health Organization (WHO).

However, these newer vaccines, starting with PCV10-GSK, are evaluated indirectly, using immunogenicity studies to infer vaccine efficacy. This process, known as immunobridging, compares immune responses from new candidate vaccines with those of licensed comparators whose efficacy has already been established.^6,7^ WHO has defined a post-immunisation immunoglobulin G (IgG) geometric mean concentration (GMC) of 0·35 μg/mL as a correlate of protection against invasive pneumococcal disease (IPD)^8^, while this is an average measure that may differ for individual serotypes, it remains a widely accepted threshold for regulatory approval.

Regulatory agencies such as the U.S. Food and Drug Administration^9^ and European Medicines Agency^10^ accept immunobridging for licensure in infants in lieu of efficacy studies if immune responses for shared serotypes meet non-inferiority criteria compared to licensed vaccines. These assessments are typically based on both IgG GMCs and seroresponse rates (i.e., proportion of participants meeting a serotype-specific protective threshold, usually 0·35 μg/mL). The downward trend in immune responses associated with expanded serotype coverage, and the successive use of non-inferiority comparisons, means that this “bridge to a bridge” process has sequentially lowered the bar needed to meet non-inferiority.^11^ There is potential for real-world vaccine effectiveness to be compromised if antibody levels induced by approved vaccines do not maintain a protective threshold.

Currently, no study has comprehensively compared the immunogenicity of PCV formulations, in combination with different dosing schedules and regional settings, which is important to design effective pneumococcal vaccination strategies. In this study, we conducted a systematic review and meta-analysis of post-vaccination immune responses in infants by serotype, vaccine product, dosing schedule and geographic region to define trends in vaccine immunogenicity and likely protection over time.

## Methods

This systematic review and meta-analysis, reported in accordance with PRISMA guidelines, investigated the immunogenicity of pneumococcal vaccination in children under 2 years. The study protocol was registered in the PROSPERO database (ID CRD42024484824).

### Search strategy and selection criteria

We formulated a search strategy using relevant keywords and medical subject heading terms for pneumococcal vaccination and immunogenicity. Five databases — EMBASE, MEDLINE, Web of Science Core Collection, Global Health, and Cochrane Central Register of Controlled Trials — were searched on May 09, 2024, with no date or study design restrictions, except for English language. An updated search was performed on Jan 07, 2025. Reference lists of included articles were screened manually for relevant studies. The full search strategy and results are provided in the appendix pp 1–5.

All identified citations were imported to online systematic review software Covidence, and duplicate removal was performed. Title and abstract screening were undertaken independently by two reviewers (XC and ST) for assessment against the eligibility criteria (appendix p 5, this includes exclusion of PCV10-GSK as the carrier proteins differ). The full texts of potentially eligible studies were retrieved, and the articles were uploaded to Covidence. Full texts were assessed by one reviewer (XC) according to the predefined eligibility criteria (appendix p 5), and a second reviewer (ST) independently checked a random subset of the full-text articles to ensure consistency. Differences at any stage were resolved through discussion with a third reviewer (PTC, DJP or JMcV).

Citations meeting inclusion criteria were grouped into “study groups”, defined as publications or abstracts arising from a single protocol, population, or surveillance system. Within each study group, we defined “study arms” as cohorts of children receiving a distinct immunisation schedule and/or PCV product. A study group could therefore include one or multiple study arms (see appendix pp 11–23 for full listings).

### Data extraction, outcomes, and quality analysis

A standardized data extraction form was developed to collect study and participant characteristics, vaccine product, schedule-related information, laboratory assay, and immunogenicity outcomes from each included study arm (see appendix pp 5–7 for definitions and p 24 and the full variable list and completeness analysis). One reviewer (XC) extracted data for each study arm, a second reviewer (ST) checked a random subset for accuracy; and discrepancies were resolved through discussion or consultation with a third reviewer (PTC, DJP or JMcV). Authors were contacted to clarify any ambiguities or discrepancies.

Immunogenicity data were categorised by blood sample collection time in relation to administered vaccine doses. Our primary outcome was serotype-specific IgG GMCs at 30 days post-childhood-schedule (“post-childhood-schedule”). Additionally, serotype-specific IgG GMCs at 30 days post-primary series (“post 1-, 2-, 3-primary dose”) and at 30 days post-booster (“post-booster”) were also collected to allow comparisons across different vaccine schedules. Each study arm could have multiple immunogenicity outcomes if data were reported at more than one timepoint. The secondary outcome was serotype-specific IgG seroresponse rates at 30 days post-childhood-schedule, post-primary series, and post-booster, defined as the proportion of participants meeting the assay-specific protective threshold against IPD. Definitions of timepoints, assay cut-off values, details of data preparation and processing, are provided in the appendix pp 6–8.

The quality of included study groups was assessed by two reviewers (XC and PTC) using modified Joanna Briggs Institute (JBI) critical appraisal tools,^12^ with each study group classified as low-, moderate- or high-risk of bias (appendix pp 8–9 and pp 25–28).

### Data analysis

Study arms were included in the main analysis if they met the following criteria: participants received their first primary dose at ≤4 months of age and last primary dose at ≤6 months of age; vaccines were administered intramuscularly; outcomes were measured using second- or third-generation ELISA (WHO ELISA), or assays bridged to WHO ELISA (see appendix p 7); study arms were from study groups with low or moderate risk of bias.

Pooled mean serotype-specific IgG GMCs with 95% confidence intervals (CIs) and seroresponse rates were estimated across vaccine products, schedules and WHO regions. Random-effects models were used to analyse log-transformed (base-2) serotype-specific IgG GMCs and logit-transformed seroresponse rates, and weights were assigned to each study arm based on the inverse variance weighting method. Between-study variance was estimated using the Paule-Mandel method, with 95% CIs obtained by the Q-profile method. For the overall random effects estimates, 95% CIs were calculated using a Wald-type normal distribution. Heterogeneity between study arms was assessed using the inconsistency index (I^2^) statistic. Sensitivity analysis is described in the appendix p 10. Publication bias was assessed using funnel plots. Data were analysed with R statistical software (version 4.3.3).

### Role of the funding source

The funder had no role in study design, data collection, data analysis, interpretation, or writing of the report.

## Results

After screening and full-text review, a total of 138 study groups from 250 articles were included (figure 1). The included study groups comprised 110 randomized controlled trials (RCTs), 14 quasi-experimental studies, and 14 prospective cohort studies. Risk of bias was low in 110 study groups, moderate in 12, and high in 16. The characteristics of the individual study groups and the modified risk of bias assessments are reported in the appendix pp 11–23.

**Figure 1.**
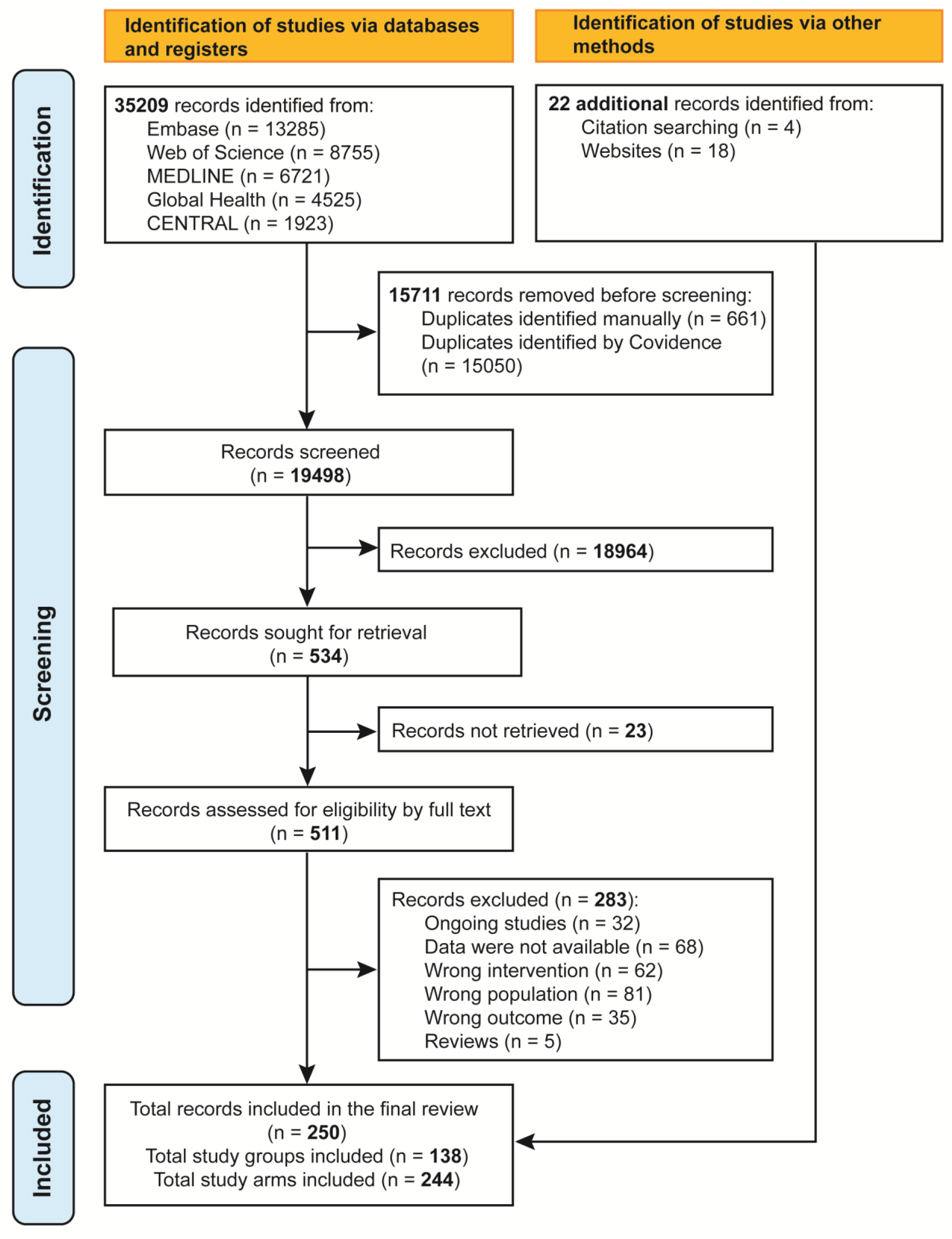
PRISMA flow diagram to show study selection process. These 138 study groups included 244 study arms. Their main characteristics and spatiotemporal distributions are summarized in table 1 and appendix (pp 29–30, 42). Most study arms investigated the immunogenicity after PCV13 (n=122) and PCV7 (n=101) vaccination, while fewer evaluated PCV15 (n=13), PCV20 (n=5) and PCV10-SII (n=3). The most common vaccine schedules were the 3+1 (n=168) and 2+1 schedule (n=49), while 3+0 (n=19), 1+1 (n=7) and 0+1 (n=1) schedules were only investigated in study arms using PCV7 and/or PCV13. Most study arms used either a 2, 4 and 6 (n=104) or 2, 3, 4 months (n=48) primary series, with booster most commonly administered at 12–15 months (n=182). Despite the highest disease burden in African and Asian countries, most study arms were conducted in the European (n=77) and Americas (n=61) regions, particularly the United States (n=42), with relatively few in the African (n=20) and South-East Asia (n=15) regions. An exception was PCV10-SII, all study arms were conducted in Gambia, African region (n=3).

There were 140 study arms that reported IgG GMCs and 88 that reported IgG seroresponse rates post-childhood-schedule (appendix p 30). Overall, pooled IgG GMCs for all vaccine-included serotypes (VTs) post-childhood-schedule were all above the WHO-defined protective threshold of 0·35 μg/mL (figure 2, and appendix pp 31–32, 43). Pooled seroresponse rates were >95% for all VTs except serotype 3 (84–92%) (appendix pp 32, 45). However, pooled IgG GMCs varied considerably across serotypes, with serotypes 14 and 6B consistently high and serotype 3 consistently low across different vaccine products, with the lowest IgG GMC observed for serotype 3 (PCV20) at 0·84 μg/mL (95%CI: 0·60–1·17). A general “downward trend” in IgG GMCs was observed with increasing vaccine valency. For PCV7-included serotypes, the IgG GMCs post PCV7 were generally higher than those post PCV13 and PCV15, except for serotype 19F. A similar trend of decreasing IgG GMCs with increasing valency was found for serotypes shared between PCV13 and higher-valency vaccines. In contrast, PCV20 did not always follow this trend, noting that CIs were wide. When data are adjusted for laboratory methods, the declining trend across increasing valency becomes apparent (appendix p 44). However, PCV10-SII was an exception, exhibiting a distinct IgG response profile, with serotypes showing high or low IgG responses differing from those in other PCVs.

**Table 1.** Characteristics of all included study arms by vaccine product.

| Characteristics | Total<br>(n=244) | PCV7<br>(n=101) | PCV13<br>(n=122) | PCV15<br>(n=13) | PCV20<br>(n=5) | PCV10-SII<br>(n=3) |
| --- | --- | --- | --- | --- | --- | --- |
| **Study period (year)** |  |  |  |  |  |  |
| 1995-1999 | 8/244 (3.3%) | 8/101 (7.9%) | - | - | - | - |
| 2000-2009 | 107/244 (43.9%) | 83/101 (82.2%) | 24/122 (19.7%) | - | - | - |
| 2010-2019 | 106/244 (43.4%) | 10/101 (9.9%) | 84/122 (68.9%) | 8/13 (61.5%) | 1/5 (20.0%) | 3/3 (100%) |
| 2020-2025 | 23/244 (9.4%) | - | 14/122 (11.5%) | 5/13 (38.5%) | 4/5 (80.0%) | - |
| **WHO region** |  |  |  |  |  |  |
| Africa | 20/244 (8.2%) | 4/101 (4.0%) | 13/122 (10.7%) | - | - | 3/3 (100%) |
| Americas | 61/244 (25.0%) | 36/101 (35.6%) | 22/122 (18.0%) | 1/13 (7.7%) | 2/5 (40.0%) | - |
| Europe | 77/244 (31.6%) | 38/101 (37.6%) | 38/122 (31.1%) | 1/13 (7.7%) | - | - |
| South-East Asia | 15/244 (6.1%) | 4/101 (4.0%) | 11/122 (9.0%) | - | - | - |
| Western Pacific | 41/244 (16.8%) | 15/101 (14.9%) | 20/122 (16.4%) | 4/13 (30.8%) | 2/5 (40.0%) | - |
| Multiple regions | 30/244 (12.3%) | 4/101 (4.0%) | 18/122 (14.8%) | 7/13 (53.8%) | 1/5 (20.0%) | - |
| **Income group** |  |  |  |  |  |  |
| Low income | 11/244 (4.5%) | - | 8/122 (6.6%) | - | - | 3/3 (100%) |
| Lower middle income | 18/244 (7.4%) | 6/101 (5.9%) | 12/122 (9.8%) | - | - | - |
| Upper middle income | 32/244 (13.1%) | 15/101 (14.9%) | 17/122 (13.9%) | - | - | - |
| Upper middle income, High income | 16/244 (6.6%) | 5/101 (5.0%) | 8/122 (6.6%) | 3/13 (23.1%) | - | - |
| High income | 167/244 (68.4%) | 75/101 (74.3%) | 77/122 (63.1%) | 10/13 (76.9%) | 5/5 (100%) | - |
| **Population type** |  |  |  |  |  |  |
| General population | 241/244 (98.8%) | 100/101 (99.0%) | 120/122 (98.4%) | 13/13 (100%) | 5/5 (100%) | 3/3 (100%) |
| Indigenous peoples | 3/244 (1.2%) | 1/101 (1.0%) | 2/122 (1.6%) | - | - | - |
| **Vaccine schedule** |  |  |  |  |  |  |
| 3+1 | 168/244 (68.9%) | 82/101 (81.2%) | 70/122 (57.4%) | 10/13 (76.9%) | 4/5 (80.0%) | 2/3 (66.7%) |
| 2+1 | 49/244 (20.1%) | 11/101 (10.9%) | 33/122 (27.0%) | 3/13 (23.1%) | 1/5 (20.0%) | 1/3 (33.3%) |
| 3+0 | 19/244 (7.8%) | 8/101 (7.9%) | 11/122 (9.0%) | - | - | - |
| 1+1 | 7/244 (2.9%) | - | 7/122 (5.7%) | - | - | - |
| 0+1 | 1/244 (0.4%) | - | 1/122 (0.8%) | - | - | - |
| **Administration of vaccine** |  |  |  |  |  |  |
| Intramuscular injection | 234/244 (95.9%) | 99/101 (98.0%) | 117/122 (95.9%) | 11/13 (84.6%) | 4/5 (80.0%) | 3/3 (100%) |
| Subcutaneous injection | 10/244 (4.1%) | 2/101 (2.0%) | 5/122 (4.1%) | 2/13 (15.4%) | 1/5 (20.0%) | - |
| **Age of first dose (Month)** |  |  |  |  |  |  |
| <2 | 25/244 (10.2%) | 7/101 (6.9%) | 16/122 (13.1%) | - | - | 2/3 (66.7%) |
| 2-4 | 213/244 (87.3%) | 92/101 (91.1%) | 103/122 (84.4%) | 12/13 (92.3%) | 5/5 (100%) | 1/3 (33.3%) |
| >4 | 5/244 (2.0%) | 2/101 (2.0%) | 2/122 (1.6%) | 1/13 (7.7%) | - | - |
| No primary series | 1/244 (0.4%) | - | 1/122 (0.8%) | - | - | - |
| **Interval between primary doses (Month)** |  |  |  |  |  |  |
| 1 | 80/244 (32.8%) | 34/101 (33.7%) | 38/122 (31.1%) | 4/13 (30.8%) | 2/5 (40.0%) | 2/3 (66.7%) |
| 2* | 156/244 (63.9%) | 67/101 (66.3%) | 76/122 (62.3%) | 9/13 (69.2%) | 3/5 (60.0%) | 1/3 (33.3%) |
| No interval | 8/244 (3.3%) | - | 8/122 (6.6%) | - | - | - |
| **Age at booster (Month)** |  |  |  |  |  |  |
| 9-11 | 32/244 (13.1%) | 10/101 (9.9%) | 20/122 (16.4%) | - | - | 2/3 (66.7%) |
| 12-15 | 182/244 (74.6%) | 76/101 (75.2%) | 87/122 (71.3%) | 13/13 (100%) | 5/5 (100%) | 1/3 (33.3%) |
| 16-18 | 11/244 (4.5%) | 7/101 (6.9%) | 4/122 (3.3%) | - | - | - |
| No booster | 19/244 (7.8%) | 8/101 (7.9%) | 11/122 (9.0%) | - | - | - |
| **Laboratory method** |  |  |  |  |  |  |
| First-generation ELISA | 2/244 (0.8%) | 2/101 (2.0%) | - | - | - | - |
| Second-generation ELISA | 14/244 (5.7%) | 13/101 (12.9%) | 1/122 (0.8%) | - | - | - |
| Third-generation ELISA | 131/244 (53.7%) | 57/101 (56.4%) | 71/122 (58.2%) | - | - | 3/3 (100%) |
| GSK 22F-ELISA | 19/244 (7.8%) | 11/101 (10.9%) | 8/122 (6.6%) | - | - | - |
| Pfizer dLIA platform | 14/244 (5.7%) | - | 9/122 (7.4%) | - | 5/5 (100%) | - |
| Pn ECL assay | 33/244 (13.5%) | 1/101 (1.0%) | 19/122 (15.6%) | 13/13 (100%) | - | - |
| FMIA | 9/244 (3.7%) | - | 9/122 (7.4%) | - | - | - |
| ELISA without detail | 22/244 (9.0%) | 17/101 (16.8%) | 5/122 (4.1%) | - | - | - |

\*Interval between primary doses was 1.5 months for one study arm and was grouped into 2 months group.

**Figure 2.**
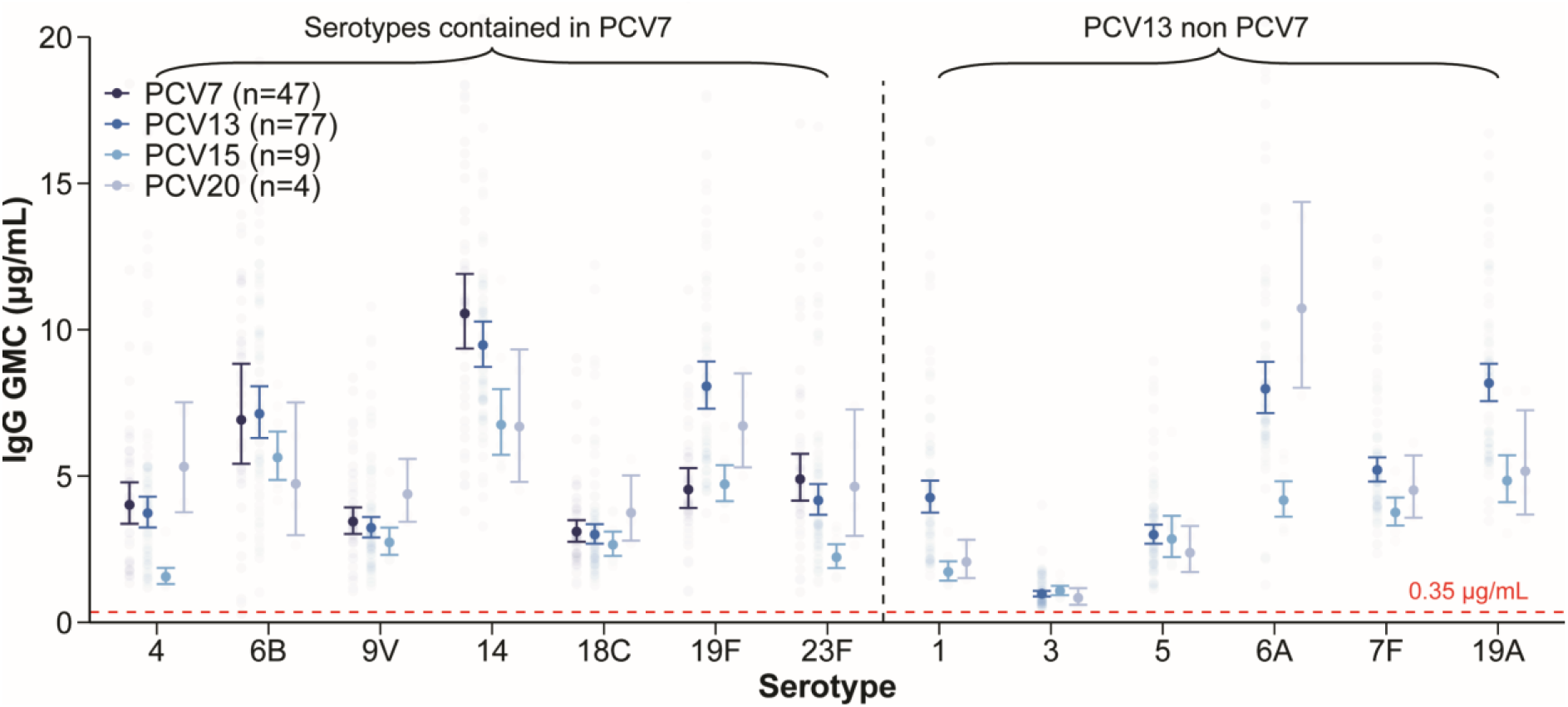
Pneumococcal IgG GMCs (μg/mL) post-childhood-schedule by serotype and vaccine product. Serotypes shared by most vaccine products are shown. The full panel of all included serotypes, as well as those unique to PCV15/20, is provided in the appendix (pp 31–32, 43). Numbers in parentheses following each vaccine name in the legend indicate the number of study arms included in the meta-analysis contributing to the pooled estimates.

Serotype-specific IgG GMCs and seroresponse rates measured post-childhood-schedule varied by vaccine schedule (see appendix p 30 for study arm numbers by schedule). Overall, vaccine schedules that included a booster dose (3+1, 2+1 or 1+1) generally elicited higher post-childhood-schedule IgG responses (both IgG GMCs and seroresponse rates; hereafter IgG responses) compared to a primary-only (3+0) schedule (figure 3a-b and appendix pp 33–35, 46–47). The 3+1 schedule generally induced immune responses that were higher than or comparable to those induced by the 2+1 schedule across most serotypes. The 1+1 schedule—although less commonly used and only tested in PCV13—produced even higher point estimates of IgG responses than 3+1/2+1 schedules for specific serotypes (e.g. serotypes 1, 14, 19F) following the booster dose.

**Figure 3.**
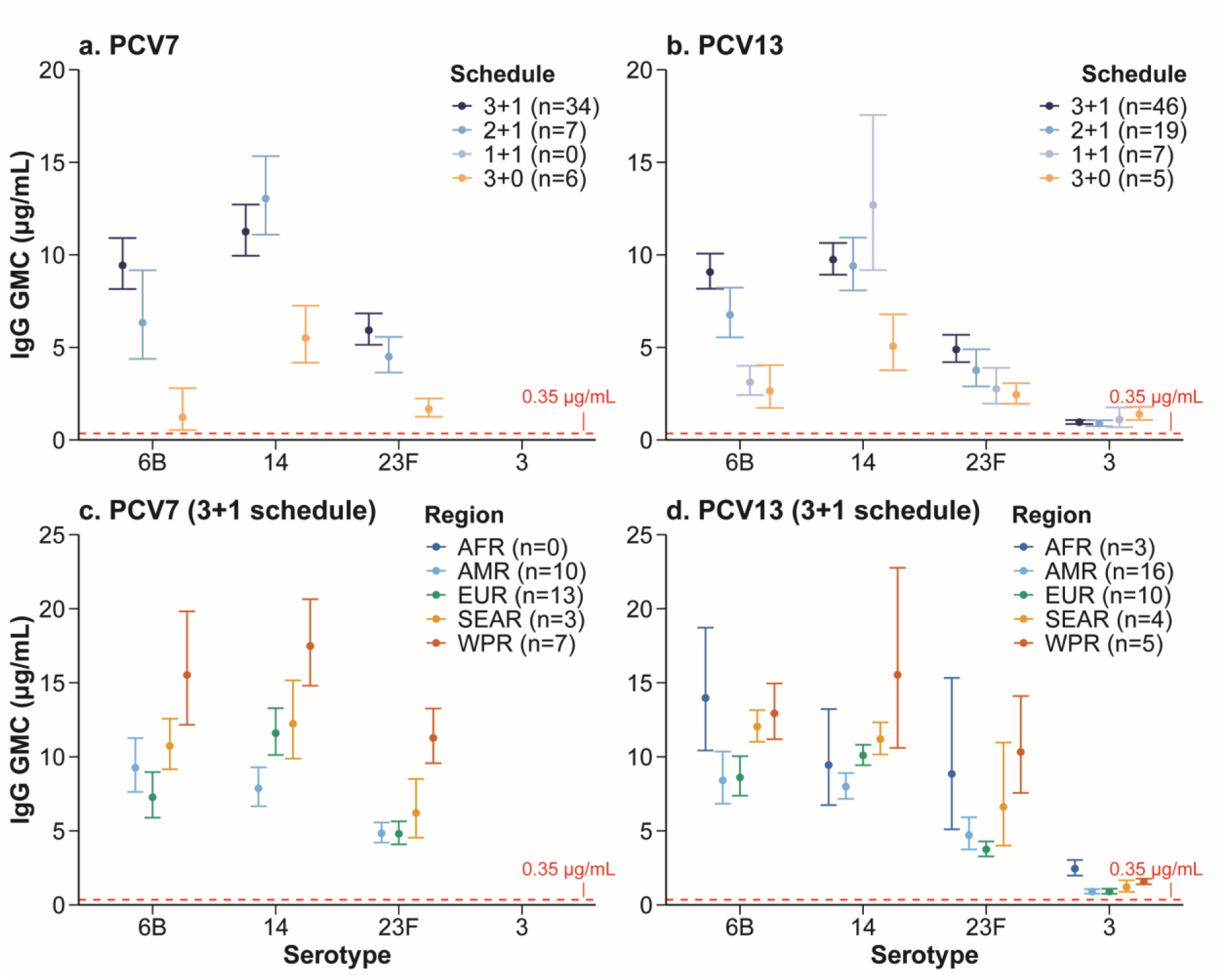
Exemplar serotype-specific pneumococcal IgG GMCs (μg/mL) post-childhood-schedule, by schedule and region. Panel (a) PCV7 stratified by schedule; (b) PCV13 stratified by schedule; (c) PCV7 stratified by region (3+1 schedule); (d) PCV13 stratified by region (3+1 schedule). Full panels are provided in the appendix (pp 46 and 48). Numbers in parentheses following each vaccine name in the legend indicate the number of study arms included in the meta-analysis contributing to the pooled estimates. Abbreviations: AFR=African Region; AMR=Region of the Americas; EUR=European Region; SEAR=South-East Asia Region; WPR=Western Pacific Region.

Given the impact of vaccine schedules on immune responses, regional analyses were restricted to study arms administering a 3+1 schedule—the most commonly used regimen—to reduce heterogeneity and improve comparability (see appendix pp 30–31 for study arm numbers by WHO region). We observed substantial regional variation of serotype-specific IgG GMCs post-childhood-schedule (figure 3c-d and appendix pp 35–37, 48). IgG GMCs were generally highest in the Western Pacific Region (WPR), followed by the African Region (AFR) and South-East Asia Region (SEAR), and lowest in the Region of the Americas (AMR), and the European Region (EUR), regardless of vaccine product. In contrast, seroresponse rates were >95% for most serotypes, except serotype 3, with minimal regional differences (appendix pp 37–39, 49).

The overall immune response post-childhood-schedule provides only part of the picture. We further assessed both serotype-specific IgG GMCs and seroresponse rates at post 1-, 2-, 3-primary doses, and post-booster (see appendix p 31 for study arm numbers by timepoint). Overall, IgG responses generally increased with the number of primary doses, and were further enhanced by a booster, although magnitude of this increase varied by serotype and vaccine; downward trends in IgG GMC with increasing valency persisted across timepoints (figure 4 and appendix pp 39–41, 50–51). Post 1-dose data (PCV7/PCV13 only) showed low IgG GMCs and seroresponse rates for most serotypes (<80%). After 2-primary doses, PCV10-SII IgG GMCs exceeded the protective threshold for VTs, as did PCV7, PCV13, and PCV15 for most serotypes (except serotypes 6B/23F and PCV15-specific 6A/33F), while PCV20 remained low (six serotypes below threshold and 13 serotypes with seroresponse rates <80%). Post 3-dose primary vaccination, all PCVs achieved GMCs above threshold, with only serotype 3 showing seroresponse rate <80%. Post-booster IgG GMCs and seroresponse rates exceeded primary series levels for all serotypes (except serotype 3). Pooled estimates across four sensitivity analyses were consistent with the main meta-analysis (appendix pp 83– 84), confirming the robustness of our findings. We found no evidence of publication bias (appendix pp 85–86).

**Figure 4.**
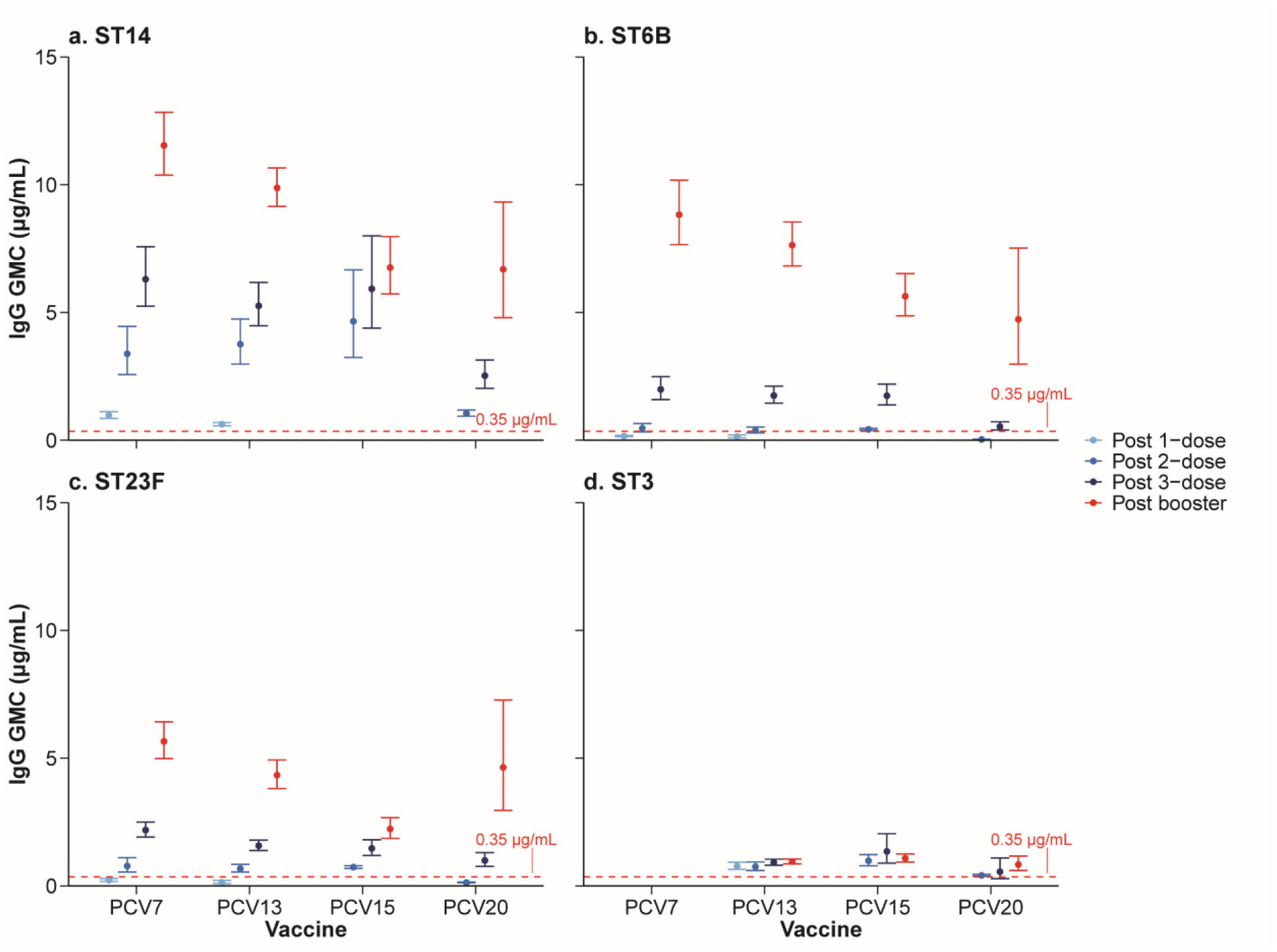
Exemplar serotype-specific pneumococcal IgG GMCs (μg/mL) by vaccine product and time point of blood sample collection. The full panel is provided in the appendix (p 50).Numbers in parentheses following each vaccine name in the legend indicate the number of study arms included in the meta-analysis contributing to the pooled estimates.

## Discussion

Our study is the largest study to systematically and quantitatively evaluate immune responses in young children for five commonly used PCVs following alternative dosing regimens across different populations and epidemiologic contexts. Our findings demonstrate that IgG responses post pneumococcal vaccination vary by serotype, PCV product, vaccine schedule, and WHO region. These immunogenicity differences should be incorporated into transmission models and health economic evaluations, especially as countries adapt vaccination programs based on their local vaccine choices, schedules, serotype distributions and disease burden.

Overall, pooled serotype-specific IgG GMCs post-childhood-schedule for all VTs were above the WHO-defined protective threshold against IPD, consistent with previous reported overall vaccine effectiveness estimates of 80·7–86·0% against VT-IPD.^13,14^ However, IgG responses varied considerably across serotypes. For some serotypes, such as serotype 3, lower IgG responses may partly explain its over-representation among vaccine failure cases,^15^ suggesting a higher correlate of protection may be required, as demonstrated by Andrews and colleagues.^16^ Consistently, multiple real-world studies have concluded little to no impact of PCV13 on reducing IPD attributed to serotype 3, with a pooled effectiveness estimated at 63·5% (95% CI: 37·3–89·7%).^17^ In contrast, serotype like 19A and 19F exhibit relatively high IgG levels that substantially exceed the correlates of protection proposed by Andrews and colleagues,^16^ yet are still frequently linked with vaccine failure,^15^ implying factors beyond antibody levels may contribute to protection. This finding suggests that vaccine effectiveness against disease outcomes varies by serotype, requiring definition of serotype-specific protective thresholds to accurately infer vaccine effectiveness.

A downward trend in IgG GMCs was generally observed with increasing valency of vaccine formulations for most serotypes, except for PCV10-SII. This exception may be related to the novel conjugation method used for PCV10-SII,^18^ which could better preserve capsular polysaccharide (CPS) integrity and enhance immune responses. Previous studies similarly found that, compared with PCV7, higher-valency PCVs (PCV10-GSK and PCV13) had lower GMCs for most serotypes.^19,20^ Given that CPS amounts per serotype were largely consistent across vaccines, this pattern is unlikely to be explained by differences in antigen amounts. This trend may reflect a potential trade-off between serotype coverage and immunogenicity, potentially due to antigenic competition and carrier-induced epitope suppression.^21^ However, as no studies used the same laboratory methods to measure immune responses after PCV15 and PCV20 vaccination, these comparisons should be interpreted with caution. Overall, this downward trend in IgG GMCs with increasing valency highlights the importance of balancing broader vaccine serotype coverage and the magnitude of immune response for sustained population protection.

Vaccine schedules that included a booster dose (3+1, 2+1 or 1+1) consistently elicited higher IgG GMCs and seroresponse rates following completion of the childhood vaccination compared to a primary-only (3+0) schedule, regardless of serotypes and vaccine products. Deloria and colleagues^22^ also reported that booster doses substantially increased GMCs for all serotypes, with 2+1 schedule yielding higher IgG response than 3+0.

Given the evidence that a 3+0 schedule, even with high coverage, provided limited population-level immunity beyond the first year of life,^23^ along with increased rates of vaccine failures in cohorts of primary-only schedule.^15,24^ This finding underscores the critical role of a booster dose in achieving optimal protection during and beyond the second year of life.

Regarding the primary series, a 3-dose schedule generally induced higher IgG responses than 2- or 1-dose schedule, although the difference between 3-dose and 2-dose was generally small, but was substantial for PCV20 recipients. Our findings partly align with an earlier review favouring 3-doses,^22^ however that review only included limited serotypes. We further included post 1-dose responses and found that a single dose of PCV7/PCV13 produced IgG GMCs lower than the WHO-defined protective threshold of 0·35 ug/mL for most serotypes. Overall, at least a 2-dose primary schedule is likely needed to induce sufficient antibody levels for short-term protection against most VTs. Notably, the largest differences were observed in serotypes 6B and 23F, for which two primary doses may not provide sufficient protection during the first year of life based on observed post-immunisation IgG responses, consistent with previous reviews.^25,26^ These findings suggest that a 3-dose primary series may be required in settings where these serotypes predominate. For PCV20 recipients, the difference between 2- and 3-doses was remarkable, indicating suboptimal immunogenicity with a 2-dose schedule and supporting the need for three doses to achieve optimal protection with PCV20.

The 1+1 schedule — one primary dose followed by a booster — presents a unique immunological profile. Our findings indicate that post-booster IgG responses following a 1+1 schedule were higher or equivalent to those after 2- or 3-dose primary series (3+1 or 2+1), except for serotype 6B, but only limited to few studies. Real-world studies have shown that dropping a primary dose from a 2+1 schedule did not increase VT carriage and vaccine failure in the second year of life.^27,28^ However, IgG responses were low after a single primary dose for most serotypes based on PCV7 and PCV13 data. Given the observed “downward trend” in immunogenicity with higher valency PCVs, post-primary responses after 1+1 schedules with PCV15 and PCV20 may be even lower, this requires confirmation. This divergence — low early immunogenicity but strong booster responses — may partly result from insufficient early protection that allows continued pneumococcal carriage during infancy, thereby enhancing the subsequent booster response. While the 1+1 schedule is promising in terms of simplification and affordability, its suitability should be carefully evaluated using transmission models that incorporate immunological parameters,^29^ particularly in settings where early-life pneumococcal exposure is common.

We observed substantial regional variation of serotype-specific IgG responses post-childhood-schedule. IgG responses were generally highest in the WPR, followed by AFR and SEAR, and lowest in AMR and EUR, regardless of vaccine product. These findings are consistent with previous studies assessing the impact of geographic region on post-primary immunogenicity: Park and colleagues^19^ reported higher GMCs in Asia, Africa, and Latin America, while Choe and colleagues^30^ similarly found enhanced responses in WPR, SEAR, and AFR. Our study extends this evidence by including newer PCVs — PCV15, PCV20, and PCV10-SII — and observed similar trends, although evidence is limited to certain regions, with PCV10-SII only being evaluated in AFR. The regional variations may reflect differences in circulating serotypes and force of infection.^2^ These findings suggest that regions with high IgG GMCs, such as WPR, may require fewer vaccine doses to achieve a clinically adequate immune response, potentially resulting in substantial cost savings. The observed regional differences also highlight the importance of reviewing immunogenicity studies in the context of their specific settings as findings may not be routinely generalisable across all global regions.

Several limitations should be noted. First, PCV10-GSK was not included because it has different carrier proteins, which improves comparability but limits coverage of all globally used PCVs. Second, functional opsonophagocytic activity (OPA), a potentially better predictor of protection than serum IgG, was not assessed in our study because substantial variability in laboratory methods across studies would make direct comparisons unreliable. Third, given large number of studies included, full text review and data extraction were performed by a single reviewer and independently checked by a second reviewer, which may introduce a small risk of error but was considered a practical and commonly accepted approach. Fourth, only English-language studies were included to ensure accurate data extraction, as English is the common language used by our team. This may have excluded relevant studies published in other languages, particularly from Asia and Africa where pneumococcal burden is highest, potentially introducing language bias.

Despite these limitations, our systematic review provided quantitative estimates of variations in vaccine immunogenicity by serotype, vaccine type, schedule and WHO region. These differences should be appropriately considered when evaluating vaccine programs. The observed downward trends in both IgG GMCs and seroresponse rates with increasing vaccine valency challenge the rationale of continually expanding PCV valency. Rather than accepting suboptimal immune responses as an inevitable trade-off, future vaccine development could benefit from novel approaches, such as optimizing conjugation strategies to enhance immunogenicity. Developing a standalone vaccine targeting problematic serotypes (e.g., serotypes 3 and 19A) for use alongside routine immunization schedules may also be a feasible strategy in settings with a substantial disease burden of these problematic serotypes.

## Data Availability

All data produced in the present study are available upon reasonable request to the authors

## Contributors

XC, PTC, DJP, KSC, and JMcV contributed to the conceptualization of the study. XC, and ST curated the data. XC performed the formal analysis. JMcV acquired funding and provided resources. XC, VS, KSC, NT, NC, PTC, DJP, and JMcV carried out the investigation. XC, PTC, DJP, and JMcV developed the methodology. XC managed the project administration. XC, PTC, and DJP contributed to software development. PTC, DJP, NC, and JMcV supervised the study. ST, and PTC performed validation and verification. XC prepared the visualizations. XC wrote the original draft, and all authors contributed to review and editing of the manuscript. All authors interpreted the data and approved the final version for submission. The corresponding author had full access to all data and final responsibility for submission.

## Declaration of interests

We declare no competing interests.

## Data sharing

All data used in this study were obtained from publicly available sources, including population-level immune response data. No new individual participant data were collected for this research. Since the data are publicly accessible, any qualified researcher can access the datasets directly from the original sources.

## Acknowledgements

This work received no dedicated funding. XC is supported by the China Scholarship Council – University of Melbourne PhD Scholarship; PTC receives salary support from SPARKLE, funded by the Australian Department for Foreign Affairs and Trade under the Partnerships for a Healthy Region Initiative grant to the Peter Doherty Institute for Infection and Immunity.

## Notes

### Competing Interest Statement

The authors have declared no competing interest.

### Clinical Protocols

https://www.crd.york.ac.uk/PROSPERO/view/CRD42024484824

### Author Declarations

The study used only openly available human data identified through searches of EMBASE, MEDLINE, Web of Science Core Collection, Global Health, and the Cochrane Central Register of Controlled Trials. No new human data were collected, and no interaction with human subjects occurred.

