## Supplementary appendix for "The impact of vaccine products, schedules and geographic regions on immunogenicity of pneumococcal vaccines in children under 2 years old: a systematic review and meta-analysis"

### Supplementary Methods

#### Supplementary Methods 1. Search strategy and corresponding results

In collaboration with biomedical reference librarians, we developed a search strategy guided by the population, intervention, comparator, outcome, study design (PICOS) framework. Our search strategy focused primarily on the intervention and outcome components. Population criteria related to age were not incorporated into the initial search, as age information is often inconsistently indexed and difficult to capture through keywords. To maintain sensitivity, a broad search strategy was employed, with age-related eligibility applied during the screening phase. Additionally, as immunogenicity is an objective outcome measure typically measured using standardized laboratory assays, the inclusion of studies without a comparator and non-randomised quasi-experimental or observational studies was considered acceptable and did not compromise the interpretability of our findings. The intervention of interest was pneumococcal conjugate vaccine, and the main outcome parameters were anti-polysaccharide immunoglobulin G (IgG) geometric mean concentration (GMC) and seroresponse rate.

**Search strategy and results in Embase**

| ID | Embase | Results up to 2024-05-09 | Results up to 2025-01-07 |
|---|---|---|---|
| 1 | Streptococcus pneumoniae/ or pneumococcal infection/ or ("streptococcus pneumonia*" or pneumococc* or "s pneumonia*" or "strep pneumonia*" or "streptococcal pneumonia*").ti,ab,kf. | 85476 | 88173 |
| 2 | exp vaccine/ or exp immunization/ or (vaccin* or immuniz* or immunis*).ti,ab,kf. | 842499 | 876156 |
| 3 | **1 and 2** | 28767 | 29916 |
| 4 | Pneumococcus vaccine/ or (pneumococc* adj5 vaccin*).ti,ab,kf. | 28822 | 29865 |
| 5 | (7vpcv or "7v pcv" or pcv7 or "pcv 7" or heptavalent or "7 valent" or 7valent or pncrm7 or "pncrm 7" or 7vpnc or 7vcrm or "seven valent" or prevnar or prevenar or prevnar7 or prevenar7).ti,ab,kf. | 3894 | 3965 |
| 6 | (10vpcv or "10v pcv" or pcv10 or "pcv 10" or "10 valent" or 10valent or "ten valent" or pneumosil or "SIIPL PCV" or "SII Pneumosil").ti,ab,kf. | 1144 | 1192 |
| 7 | (13vpcv or "13v pcv" or pcv13 or "pcv 13" or 13vcrm or "13 valent" or 13valent or "thirteen valent" or prevenar13 or prevnar13).ti,ab,kf. | 4049 | 4269 |
| 8 | (15vpcv or "15v pcv" or pcv15 or "pcv 15" or "15 valent" or 15valent or "fifteen valent" or vaxneuvance or v114).ti,ab,kf. | 318 | 385 |
| 9 | (20vpcv or "20v pcv" or pcv20 or "pcv 20" or "20 valent" or 20valent or "twenty valent" or prevnar20 or prevenar20 or apexxnar).ti,ab,kf. | 237 | 340 |
| 10 | (21vpcv or "21v pcv" or pcv21 or "pcv 21" or "21 valent" or 21valent or "twenty one valent" or v116).ti,ab,kf. | 57 | 85 |
| 11 | (ppv23 or "ppv 23" or 23vppv or "23v ppv" or ppsv23 or "ppsv 23" or "23 valent" or 23valent or pneumovax or "pneumovax 23" or pneumovax23 or "pneumo 23").ti,ab,kf. | 3184 | 3286 |
| 12 | **3 or 4 or 5 or 6 or 7 or 8 or 9 or 10 or 11** | 35885 | 37277 |
| 13 | exp immunity/ or antibody/ or immunoglobulin G/ or (immun* or antibod* or "immunoglobulin G" or IgG or opsonophagocyt* or OPA).ti,ab,kf. | 5228712 | 5423693 |
| 14 | **12 and 13** | 19951 | 20743 |
| 15 | (exp in vitro study/ or exp animal experiment/ or exp animal/ or exp juvenile animal/ or adult animal/ or animal cell/ or animal tissue/ or nonhuman/ or animal model/ or exp invertebrate/ or exp plant/ or exp fungus/) not (exp human/ or human experiment/) | 9195544 | 9406874 |
| 16 | (("in vitro" or animal or animals or canine* or dog or dogs or cat or cats or feline or hamster* or lamb or lambs or mice or ferret* or primate* or macaque* or monkey or monkeys or mouse or murine or swine or pig or pigs or piglet* or porcine or rabbit* or rat or rats or rodent* or sheep* or bovine or cow or cows or horse or horses or poultry or chick or chicken* or turkey* or avian* or vertebrate* or veterinary* or plant* or fung*) not (human* or patient* or people or mankind*)).ti,ab,kf. | 6175695 | 6325725 |
| 17 | **15 or 16** | 10347740 | 10601649 |

| ID | Embase | Results up to 2024-05-09 | Results up to 2025-01-07 |
|---|---|---|---|
| 18 | **14 not 17** | 17417 | 18143 |
| 19 | limit 18 to ("review" or editorial) | 3891 | 3993 |
| 20 | **18 not 19** | 13526 | 14150 |
| 21 | Language: English | 12702 | 13316 |
| | limit 21 to dc=20240510-20250107 (for update search only) | - | 583 |

### Search strategy and results in Medline

| ID | Medline (Ovid) | Results up to 2024-05-09 | Results up to 2025-01-07 |
|---|---|---|---|
| 1 | Streptococcus pneumoniae/ or Pneumococcal Infections/ or ("streptococcus pneumonia*" or pneumococc* or "s pneumonia*" or "strep pneumonia*" or "streptococcal pneumonia*").ti,ab,kf. | 53363 | 54390 |
| 2 | exp Vaccines/ or exp Immunization/ or (vaccin* or immuniz* or immunis*).ti,ab,kf. | 630284 | 650985 |
| 3 | **1 and 2** | 18252 | 18767 |
| 4 | Pneumococcal Vaccines/ or (pneumococc* adj5 vaccin*).ti,ab,kf. | 14557 | 14971 |
| 5 | (7vpcv or "7v pcv" or pcv7 or "pcv 7" or heptavalent or "7 valent" or 7valent or pncrm7 or "pncrm 7" or 7vpnc or 7vcrm or "seven valent" or prevnar or prevenar or prevnar7 or prevenar7).ti,ab,kf. | 2938 | 2971 |
| 6 | (10vpcv or "10v pcv" or pcv10 or "pcv 10" or "10 valent" or 10valent or "ten valent" or pneumosil or "SIIPL PCV" or "SII Pneumosil").ti,ab,kf. | 893 | 929 |
| 7 | (13vpcv or "13v pcv" or pcv13 or "pcv 13" or 13vcrm or "13 valent" or 13valent or "thirteen valent" or prevenar13 or prevnar13).ti,ab,kf. | 2823 | 2955 |
| 8 | (15vpcv or "15v pcv" or pcv15 or "pcv 15" or "15 valent" or 15valent or "fifteen valent" or vaxneuvance or v114).ti,ab,kf. | 216 | 259 |
| 9 | (20vpcv or "20v pcv" or pcv20 or "pcv 20" or "20 valent" or 20valent or "twenty valent" or prevnar20 or prevenar20 or apexxnar).ti,ab,kf. | 180 | 231 |
| 10 | (21vpcv or "21v pcv" or pcv21 or "pcv 21" or "21 valent" or 21valent or "twenty one valent" or v116).ti,ab,kf. | 30 | 47 |
| 11 | (ppv23 or "ppv 23" or 23vppv or "23v ppv" or ppsv23 or "ppsv 23" or "23 valent" or 23valent or pneumovax or "pneumovax 23" or pneumovax23 or "pneumo 23").ti,ab,kf. | 2075 | 2138 |
| 12 | **3 or 4 or 5 or 6 or 7 or 8 or 9 or 10 or 11** | 19162 | 19693 |
| 13 | exp Immunity/ or Immunoglobulin G/ or Antibodies/ or (immun* or antibod* or "immunoglobulin G" or IgG or opsonophagocyt* or OPA).ti,ab,kf. | 3545895 | 3663361 |
| 14 | **12 and 13** | 10196 | 10488 |
| 15 | (exp Animals/ or Disease Models, Animal/ or exp Animal Experimentation/ or exp Plants/ or exp Fungi/) not (exp Humans/ or Human Experimentation/) | 5659448 | 5746803 |
| 16 | (("in vitro" or animal or animals or canine* or dog or dogs or cat or cats or feline or hamster* or lamb or lambs or mice or ferret* or primate* or macaque* or monkey or monkeys or mouse or murine or swine or pig or pigs or piglet* or porcine or rabbit* or rat or rats or rodent* or sheep* or bovine or cow or cows or horse or horses or poultry or chick or chicken* or turkey* or avian* or vertebrate* or veterinary* or plant* or fung*) not (human* or patient* or people or mankind*)).ti,ab,kf. | 5135215 | 5261822 |
| 17 | **15 or 16** | 7264974 | 7425755 |
| 18 | **14 not 17** | 8560 | 8830 |
| 19 | limit 18 to ("review" or editorial) | 1643 | 1690 |
| 20 | **18 not 19** | 6917 | 7140 |
| 21 | limit 20 to english language | 6507 | 6721 |

| ID | Medline (Ovid) | Results up to 2024-05-09 | Results up to 2025-01-07 |
|---|---|---|---|
| | limit 21 to dt=20240510-20250107 (for update search only) | - | 214 |

#### Search strategy and results in Web of Science

| ID | Web of Science | Results up to 2024-05-13 | Results up to 2025-01-07 |
|---|---|---|---|
| 1 | TS=("streptococcus pneumonia*" or pneumococc* or "s pneumonia*" or "strep pneumonia*" or "streptococcal pneumonia*") | 60335 | 62504 |
| 2 | TS=(vaccin* or immuniz* or immunis*) | 598169 | 632305 |
| 3 | **#1 and #2** | 20934 | 21774 |
| 4 | TS=(pneumococc* NEAR/5 vaccin*) | 15298 | 15900 |
| 5 | TS=(7vpcv or "7v pcv" or pcv7 or "pcv 7" or heptavalent or "7 valent" or 7valent or pncrm7 or "pncrm 7" or 7vpnc or 7vcrm or "seven valent" or prevnar or prevenar or prevnar7 or prevenar7) | 3122 | 3164 |
| 6 | TS=(10vpcv or "10v pcv" or pcv10 or "pcv 10" or "10 valent" or 10valent or "ten valent" or pneumosil or "SIIPL PCV" or "SII Pneumosil") | 920 | 965 |
| 7 | TS=(13vpcv or "13v pcv" or pcv13 or "pcv 13" or 13vcrm or "13 valent" or 13valent or "thirteen valent" or prevenar13 or prevnar13) | 2971 | 3126 |
| 8 | TS=(15vpcv or "15v pcv" or pcv15 or "pcv 15" or "15 valent" or 15valent or "fifteen valent" or vaxneuvance or v114) | 222 | 263 |
| 9 | TS=(20vpcv or "20v pcv" or pcv20 or "pcv 20" or "20 valent" or 20valent or "twenty valent" or prevnar20 or prevenar20 or apexxnar) | 180 | 236 |
| 10 | TS=(21vpcv or "21v pcv" or pcv21 or "pcv 21" or "21 valent" or 21valent or "twenty one valent" or v116) | 27 | 41 |
| 11 | TS=(ppv23 or "ppv 23" or 23vppv or "23v ppv" or ppsv23 or "ppsv 23" or "23 valent" or 23valent or pneumovax or "pneumovax 23" or pneumovax23 or "pneumo 23") | 2047 | 2146 |
| 12 | **#3 or #4 or #5 or #6 or #7 or #8 or #9 or #10 or #11** | 21608 | 22480 |
| 13 | TS=(immun* or antibod* or "immunoglobulin G" or IgG or opsonophagocyt* or OPA) | 3885130 | 4101149 |
| 14 | **#12 and #13** | 12309 | 12844 |
| 15 | TS=(("in vitro" or animal or animals or canine* or dog or dogs or cat or cats or feline or hamster* or lamb or lambs or mice or ferret* or primate* or macaque* or monkey or monkeys or mouse or murine or swine or pig or pigs or piglet* or porcine or rabbit* or rat or rats or rodent* or sheep* or bovine or cow or cows or horse or horses or poultry or chick or chicken* or turkey* or avian* or vertebrate* or veterinary* or plant* or fung*) not (human* or patient* or people or mankind*)) | 8352238 | 8729305 |
| 16 | **#14 not #15** | 10742 | 11198 |
| 17 | **#14 not #15** and **Review Article** or **Editorial Material** (Exclude – Document Types) and **English** (Languages) | 8498 | 8905 |
| | **#17 and LD=(2024-05-14 to 2025-01-07)** (for update search only) | - | 257 |

#### Search strategy and results in Global Health

| ID | Global Health (since 1973) | Results up to 2024-05-13 | Results up to 2025-01-07 |
|---|---|---|---|
| 1 | Streptococcus pneumoniae/ or ("streptococcus pneumonia*" or pneumococc* or "s pneumonia*" or "strep pneumonia*" or "streptococcal pneumonia*").ti,ab,id. | 21041 | 22042 |
| 2 | exp vaccines/ or exp immunization/ or (vaccin* or immuniz* or immunis*).ti,ab,id. | 208091 | 209029 |

| ID | Global Health (since 1973) | Results up to 2024-05-13 | Results up to 2025-01-07 |
|---|---|---|---|
| 3 | **1 and 2** | 9489 | 9512 |
| 4 | (pneumococc* adj5 vaccin*).ti,ab,id. | 7157 | 7227 |
| 5 | (7vpcv or "7v pcv" or pcv7 or "pcv 7" or heptavalent or "7 valent" or 7valent or pncrm7 or "pncrm 7" or 7vpnc or 7vcrm or "seven valent" or prevnar or prevenar or prevnar7 or prevenar7).ti,ab,id. | 2198 | 2224 |
| 6 | (10vpcv or "10v pcv" or pcv10 or "pcv 10" or "10 valent" or 10valent or "ten valent" or pneumosil or "SIIPL PCV" or "SII Pneumosil").ti,ab,id. | 642 | 683 |
| 7 | (13vpcv or "13v pcv" or pcv13 or "pcv 13" or 13vcrm or "13 valent" or 13valent or "thirteen valent" or prevenar13 or prevnar13).ti,ab,id. | 1982 | 2082 |
| 8 | (15vpcv or "15v pcv" or pcv15 or "pcv 15" or "15 valent" or 15valent or "fifteen valent" or vaxneuvance or v114).ti,ab,id. | 118 | 138 |
| 9 | (20vpcv or "20v pcv" or pcv20 or "pcv 20" or "20 valent" or 20valent or "twenty valent" or prevnar20 or prevenar20 or apexxnar).ti,ab,id. | 109 | 138 |
| 10 | (21vpcv or "21v pcv" or pcv21 or "pcv 21" or "21 valent" or 21valent or "twenty one valent" or v116).ti,ab,id. | 13 | 17 |
| 11 | (ppv23 or "ppv 23" or 23vppv or "23v ppv" or ppsv23 or "ppsv 23" or "23 valent" or 23valent or pneumovax or "pneumovax 23" or pneumovax23 or "pneumo 23").ti,ab,id. | 1198 | 1233 |
| 12 | **3 or 4 or 5 or 6 or 7 or 8 or 9 or 10 or 11** | 9517 | 9637 |
| 13 | exp immunity/ or antibodies/ or IgG/ or (immun* or antibod* or "immunoglobulin G" or IgG or opsonophagocyt* or OPA).ti,ab,id. | 670571 | 671674 |
| 14 | **12 and 13** | 5049 | 5143 |
| 15 | (in vitro/ or animal experiments/ or animals/ or animal tissues/ or animal models/ or invertebrates/ or plants/ or fungi/) not (man/ or human diseases/) | 1272514 | 1274215 |
| 16 | (("in vitro" or animal or animals or canine* or dog or dogs or cat or cats or feline or hamster* or lamb or lambs or mice or ferret* or primate* or macaque* or monkey or monkeys or mouse or murine or swine or pig or pigs or piglet* or porcine or rabbit* or rat or rats or rodent* or sheep* or bovine or cow or cows or horse or horses or poultry or chick or chicken* or turkey* or avian* or vertebrate* or veterinary* or plant* or fung*) not (human* or patient* or people or mankind*)).ti,ab,id. | 1003594 | 1005224 |
| 17 | **15 or 16** | 1563115 | 1564815 |
| 18 | **14 not 17** | 4703 | 4812 |
| 19 | limit 18 to editorial | 7 | 3 |
| 20 | **18 not 19** | 4696 | 4809 |
| 21 | limit 20 to english language | 4425 | 4525 |
| | limit 21 to yr="2024 -Current" (for update search only) | - | 115 |

### Search strategy and results in Cochrane Central Register of Controlled Trials

| ID | Cochrane Central Register of Controlled Trials | Results up to 2024-05-13 | Results up to 2025-01-07 |
|---|---|---|---|
| 1 | Streptococcus pneumoniae/ or Pneumococcal Infections/ or ("streptococcus pneumonia*" or pneumococc* or "s pneumonia*" or "strep pneumonia*" or "streptococcal pneumonia*").ti,ab,kf. | 3627 | 3751 |
| 2 | exp Vaccines/ or exp Immunization/ or (vaccin* or immuniz* or immunis*).ti,ab,kf. | 34575 | 36105 |
| 3 | **1 and 2** | 2340 | 2443 |
| 4 | Pneumococcal Vaccines/ or (pneumococc* adj5 vaccin*).ti,ab,kf. | 2105 | 2192 |

| ID | Cochrane Central Register of Controlled Trials | Results up to 2024-05-13 | Results up to 2025-01-07 |
|---|---|---|---|
| 5 | (7vpcv or "7v pcv" or pcv7 or "pcv 7" or heptavalent or "7 valent" or 7valent or pncrm7 or "pncrm 7" or 7vpnc or 7vcrm or "seven valent" or prevnar or prevenar or prevnar7 or prevenar7).ti,ab,kf. | 681 | 696 |
| 6 | (10vpcv or "10v pcv" or pcv10 or "pcv 10" or "10 valent" or 10valent or "ten valent" or pneumosil or "SIIPL PCV" or "SII Pneumosil").ti,ab,kf. | 203 | 213 |
| 7 | (13vpcv or "13v pcv" or pcv13 or "pcv 13" or 13vcrm or "13 valent" or 13valent or "thirteen valent" or prevenar13 or prevnar13).ti,ab,kf. | 580 | 610 |
| 8 | (15vpcv or "15v pcv" or pcv15 or "pcv 15" or "15 valent" or 15valent or "fifteen valent" or vaxneuvance or v114).ti,ab,kf. | 118 | 125 |
| 9 | (20vpcv or "20v pcv" or pcv20 or "pcv 20" or "20 valent" or 20valent or "twenty valent" or prevnar20 or prevenar20 or apexxnar).ti,ab,kf. | 50 | 65 |
| 10 | (21vpcv or "21v pcv" or pcv21 or "pcv 21" or "21 valent" or 21valent or "twenty one valent" or V116).ti,ab,kf. | 23 | 30 |
| 11 | (ppv23 or "ppv 23" or 23vppv or "23v ppv" or ppsv23 or "ppsv 23" or "23 valent" or 23valent or pneumovax or "pneumovax 23" or pneumovax23 or "pneumo 23").ti,ab,kf. | 544 | 562 |
| 12 | **3 or 4 or 5 or 6 or 7 or 8 or 9 or 10 or 11** | 2487 | 2594 |
| 13 | exp Immunity/ or Antibodies/ or Immunoglobulin G/ or (immun* or antibod* or "immunoglobulin G" or IgG or opsonophagocyt* or OPA).ti,ab,kf. | 138967 | 146528 |
| 14 | **12 and 13** | 1898 | 1982 |
| 15 | (exp Animals/ or Disease Models, Animal/ or exp Animal Experimentation/ or exp Plants/ or exp Fungi/) not (exp Humans/ or Human Experimentation/) | 3871 | 3871 |
| 16 | (("in vitro" or animal or animals or canine* or dog or dogs or cat or cats or feline or hamster* or lamb or lambs or mice or ferret* or primate* or macaque* or monkey or monkeys or mouse or murine or swine or pig or pigs or piglet* or porcine or rabbit* or rat or rats or rodent* or sheep* or bovine or cow or cows or horse or horses or poultry or chick or chicken* or turkey* or avian* or vertebrate* or veterinary* or plant* or fung*) not (human* or patient* or people or mankind*)).ti,ab,kf. | 26716 | 28034 |
| 17 | **15 or 16** | 28331 | 29650 |
| 18 | **14 not 17** | 1869 | 1953 |
| 19 | Language: English | 1851 | 1935 |
| | limit 19 to yr="2024 -Current" (for update search only) | | 72 |

### Supplementary Methods 2. Inclusion and exclusion criteria

Eligible studies met the following inclusion criteria: (1) Studies investigating any of the five licensed pneumococcal conjugate vaccines (PCV7, PCV13, PCV10-SII, PCV15, and PCV20; detailed characteristics are provided in the Supplementary Methods 3, pp 8–9) in healthy paediatric populations aged 2 years and under; (2) Studies reporting either antibody responses assessed by anti-polysaccharide IgG geometric mean antibody concentration (GMC) and 95% confidence intervals (CIs) in micrograms per millilitre (µg/mL) or IgG seroresponse rates for at least one time point of a) between 4 and 6 weeks after the primary vaccination series, b) and/or 4 and 6 weeks after a booster vaccination.

Exclusion criteria: Studies that measured antibody responses by radioimmunoassay (RIA) or hemagglutination; Studies focused on immunocompromised patients; Studies in preterm infants only; Studies related to maternal vaccination and related infant immune response; Studies on infants born to HIV-infected mothers, whether prenatally infected or exposed but uninfected; Studies with only opsonophagocytic assay (OPA) results; Studies with only salivary IgG response results.

### Supplementary Methods 3. Included pneumococcal conjugate vaccines and their characteristics

| Vaccine | Licensed (FDA) | Serotypes contained in the vaccine | Carrier protein | Amounts of antigens (components) |
|---|---|---|---|---|
| PCV7 | February 2000 | 4, 6B, 9V, 14, 18C, 19F, 23F | Diphtheria toxoid-derived recombinant Cross-Reactive-Material 197 (CRM197) | Each 0·5mL dose contains 2 µg of pneumococcal purified capsular polysaccharides (CPS) for serotypes (STs) 4, 9V, 14, 18C, 19F, 23F, 4 µg of pneumococcal purified CPS for ST 6B; 20 µg CRM197; |

| Vaccine | Licensed (FDA) | Serotypes contained in the vaccine | Carrier protein | Amounts of antigens (components) |
|---|---|---|---|---|
| | | | | adsorbed on aluminium phosphate adjuvant (0·125 mg) |
| PCV13 | February 2010 | 4, 6B, 9V, 14, 18C, 19F, 23F, 1, 3, 5, 6A, 7F, 19A | CRM197 | Each 0·5mL dose contains 2·2 µg of pneumococcal purified CPS for STs 1, 3, 4, 5, 6A, 7F, 9V, 14, 18C, 19A, 19F, and 23F, 4·4 µg of pneumococcal purified CPS for ST 6B; 32 µg CRM197; adsorbed on aluminium phosphate adjuvant (0·565 mg) |
| PCV10-SII* | December 2019□ | 6B, 9V, 14, 19F, 23F, 1, 5, 6A, 7F, 19A | CRM197 | Each 0·5mL dose contains 2 µg of pneumococcal purified CPS for STs 1, 5, 9V, 14, 19A, 19F, 23F, 7F, and 6A, and 4 µg of pneumococcal purified CPS for ST 6B; each ST is individually conjugated to the CRM197 (19 to 48 µg); adsorbed on aluminium phosphate adjuvant (0·125 mg) |
| PCV20 | June 2021 | 4, 6B, 9V, 14, 18C, 19F, 23F, 1, 3, 5, 6A, 7F, 19A, 22F, 33F, 8, 10A, 11A, 12F, 15B | CRM197 | Each 0·5mL dose contains 2·2 µg of pneumococcal purified capsular polysaccharides (CPS) for serotypes (STs) 1, 3, 4, 5, 8, 6A, 7F, 9V, 10A, 11A, 12F, 15B, 14, 18C, 19A, 19F 22F,23F, 33F, 4·4 µg of pneumococcal purified CPS for ST 6B; 65 µg CRM197; adsorbed on aluminium phosphate adjuvant (0·565 mg) |
| PCV15 | July 2021 | 4, 6B, 9V, 14, 18C, 19F, 23F, 1, 3, 5, 6A, 7F, 19A, 22F, 33F | CRM197 | Each 0·5mL dose contains 2 µg of pneumococcal purified CPS for STs 1, 3, 4, 5, 6A, 7F, 9V, 14, 18C, 19A, 19F, 22F, 23F, and 33F, 4 µg of pneumococcal purified CPS for ST 6B; 30 µg CRM197; adsorbed on aluminium phosphate adjuvant (0·125 mg) |

*PCV10-SII (PNEUMOSIL) received the World Health Organization (WHO) prequalification in December 2019.

### Supplementary Methods 4. Pneumococcal vaccine schedules

In our study, vaccination schedules were classified as follows: "1+1" referred to one primary dose with one PCV booster dose, respectively; "2+1" referred to two primary doses with a booster; "3+0" and "3+1" referred to three primary doses without and with a booster; "0+1" represented one included study arm with only a single PCV dose given at 12 months of age. Additionally, "1-dose", "2-dose", and "3-dose" primary schedules referred to vaccination schedules consisting of one, two, and three primary doses, respectively.

### Supplementary Methods 5. Definitions of study timepoints

#### Childhood schedule

In our study, "childhood schedule" was defined as any study arm with immunogenicity data after the completion of pneumococcal childhood vaccination schedule. For schedules that include a booster dose (e.g., "2+1" or "3+1"), post-childhood-schedule refers to immunogenicity data obtained after the administration of the booster dose. For schedules without a booster dose (e.g., "3+0"), post-childhood-schedule refers to immunogenicity data collected after completion of the final primary dose.

#### Primary series

In our study, "1 primary dose" was defined as any study arm with immunogenicity data after the first primary dose, including those that later received a second or third dose. Similarly, "2 primary doses" and "3 primary doses" were defined as study arms with immunogenicity data following the second and third primary doses, respectively, regardless of whether a booster dose was administered. For infants receiving a three dose primary series measures reported after each dose were described as "1 primary dose", "2 primary doses", and "3 primary doses" respectively.

#### Booster

In our study, a booster dose was defined as a PCV dose administered between 9 and 18 months of age in infants who had completed a 1-, 2-, or 3-dose primary series.

#### Supplementary Methods 6. Included assays for the quantitation of pneumococcal IgG antibody and assay-specific protective thresholds

| Assay | Preadsorption | Aligned with WHO protective threshold* | Main analysis | Key features |
|---|---|---|---|---|
| First-generation ELISA[1] | None | - | No | Overestimates antibody levels due to detection of both anti-capsular and C-PS antibodies |
| Second-generation ELISA[2] | Pneumococcal C-PS | Yes, 0·35 µg/mL | Yes | Used in PCV7 pivotal trials<br>Specificity improved vs 1st-gen ELISA, but still suboptimal |
| Third-generation ELISA (WHO reference ELISA)[3-7] | C-PS and 22F PS | 0·35 µg/mL | Yes | WHO gold standard assay |
| GSK 22F-ELISA[8] | C-PS and 22F PS | No, 0·20 µg/mL | No | Used 0·20 µg/mL cut-off (vs WHO's 0·35 µg/mL) |
| Pfizer direct Luminex-based immunoassay (dLIA)[9,10] | C-PS and 22F PS | Partial yes. Serotype-specific: mostly 0·35 µg/mL; 5 = 0·23, 6B = 0·10, 19A = 0·12 | Yes (bridged to WHO ELISA) | - |
| Pneumococcal electrochemiluminescence (Pn ECL) assay[11,12] | C-PS and serotypes 25 and 72 | Yes, 0·35 µg/mL | Yes (bridged to WHO ELISA) | - |
| Fluorescent multiplex immunoassay (FMIA)[13-15] | C-PS and 22F PS | No, 0·35 µg/mL (threshold not formally validated) | No | Good agreement with WHO ELISA ($R^2 > 0·8$);<br>0·35 µg/mL threshold not formally validated for FMIA<br>Cross-reactivity present |

*The WHO has defined a protective IgG concentration cut-off against IPD of 0·35 µg/mL for all serotypes based on model results from three major clinical trials. The WHO-defined protective threshold of 0·35 µg/mL for all serotypes was used for second-generation ELISA, third-generation ELISA (WHO ELISA was derived from third-generation ELISA), corresponding to 0·20 µg/mL for GSK's 22F-ELISA. For newly developed assays, a threshold of 0·35 µg/mL was applied for both the Pneumococcal (Pn) electrochemiluminescence (ECL)-based detection assay and FMIA methods. For the Pfizer Luminex-based direct immunoassay (dLIA), most serotypes used the 0·35 µg/mL cut-off, except for serotype 5 (0·23 µg/mL), 6B (0·10 µg/mL), and 19A (0·12 µg/mL), based on bridging studies to the WHO reference ELISA.

#### Supplementary Methods 7. Data preparation and processing

Each unique study group and study arm were assigned a distinct identifier to facilitate data processing and analysis. Specifically, a unique study ID and a unique study arm ID were generated to ensure accurate tracking of data from different sources and intervention arms.

If the study start year was missing, we used the year of publication instead. We then categorized the study period into four intervals: 1995–1999, 2000–2009, 2010–2019, and 2020–2025. For studies that spanned across multiple time intervals, we calculated the mean of the start and end years and classified the study based on this average. For example, a study conducted from 2005 to 2011 would have a midpoint of 2008 and thus be categorized under 2000–2009.

The WHO region codelist used in our study was retrieved from WHO Global Health Observatory metadata, and the country income group was retrieved from The World Bank. To incorporate contextual information, we used study site locations to match their corresponding WHO region and country income group. For multisite studies that spanned multiple WHO regions, we classified them as "multi-region".

Of the 138 included study groups, 47 (34·1%) did not report any information on race or ethnicity. Among the remaining 91 study groups, 41 study groups (29·7%) reported categories of race (primarily Asian, Black, White, and Other). Eight study groups (5·8%) reported categories of nationality or ethnicity (e.g., Chinese, Japanese), while other study groups (n = 20, 14·5%) combined race and ethnicity into a single categorization (e.g., White, Hispanic American, Multiracial, Black, Asian, Native American, Other; or African heritage/African American, Asian-East Asian heritage, White-Arabic/North African heritage, White-Caucasian/European heritage, Other). Only 22 study groups (n = 15·9%) reported race and ethnicity separately. Due to the heterogeneity in how race/ethnicity was defined and reported across studies, we did not further group participants by race or ethnicity but instead presented the data as reported in the original sources. For studies specifically designed for Indigenous populations, we used a binary indicator of Indigenous status (yes = 3, 2·2%) to distinguish these study groups.

If the timing of the vaccine dosing schedule was not reported, we referred to the recommended national immunisation schedule of the country for the year in which the study was conducted. The timing of each dose was determined based on the reported age at vaccination. Specifically, the age at first dose and age at booster were extracted directly from the study if available. If age-specific data were not reported, we inferred the timing of each dose using the stated dosing schedule. For example, for participants receiving a “3+1” schedule with doses administered at 2, 4, 6, and 12 months, the age at first dose was assumed to be 2 months, and the age at booster 12 months.

We further categorised the age at first dose into three groups: <6 weeks, 6–14 weeks, and >14 weeks; and the age at booster into three groups: 9–11 months, 12–15 months, and 16–18 months. If the reported age was given as a range, the midpoint of the range was used to determine the appropriate category. For example, if the age at booster was reported as 10–14 months, the midpoint (12 months) was used, and the data were classified into the 12–15 months group. The dosing schedule was also used to determine the interval between primary doses, which was defined as the number of months between the first and second primary doses. If the intervals between the first and second doses and between the second and third doses differed, their average was used. To standardise categorisation across studies, both the timing of each dose and the interval between doses were rounded to the nearest two weeks.

If the population size of a certain intervention arm was reported as a range, we calculated the mean of the range and used it as the final population size.

With respect to sampling time, we included only results measured approximately 30 days post-vaccination (i.e., 4–6 weeks after immunisation). Specifically, this included results reported at 30 days following the first, second, or third primary dose; 30 days post-booster dose; or, more generally, 30 days after completion of the infant primary series. Data collected at other time points, such as 1 week, 6 months, or 12 months post-vaccination, were excluded from the analysis.

To ensure consistency across results, 95%CIs for seroresponse rates were calculated using Fisher’s exact test, based on the reported number of seropositive participants and the total number of participants.

### Supplementary Methods 8. Quality assessment tools

The original Joanna Briggs Institute (JBI) critical appraisal tools for randomized controlled trials (RCTs) [16], quasi-experimental studies[17], and cohort studies[18] were modified to align with the specific objectives of our review, focusing on key domains most relevant to each study design. For RCTs, we retained seven questions across five domains from the original 13-question tool. For quasi-experimental studies, we included six questions covering five domains from the original nine-question tool. For cohort studies, we selected six questions across five domains from the original 11-question tool.

Taking the JBI tool for RCTs as an example, we excluded questions deemed unrelated to our study outcomes. Given that immunogenicity is an objective outcome typically measured using standardized laboratory assays, factors such as blinding of participants, treatment providers, and outcome assessors—as well as randomization and allocation concealment—were considered unlikely to influence the measurement of outcomes. Therefore, we retained only the items deemed relevant to the internal validity of immunogenicity results. A similar approach was applied to JBI tools of quasi-experimental studies and cohort studies. Details of the modifications are provided in following tables (pp 13–16).

We did not apply numerical cut-off scores to categorize study quality. Instead, each study was assessed individually using the modified JBI tools and classified as having low-, moderate-, or high-risk of bias based on a case-by-case evaluation. Detailed risk of bias assessments are presented in Table S3–5. Quality assessment was conducted jointly by two reviewers, and any disagreements were resolved through discussion until consensus was reached.

**a. The original and modified JBI tools for randomized clinical trials (RCTs)**

| Domains | Questions in JBI tool | Questions in modified JBI tool |
|---|---|---|
| Bias related to selection and allocation | Q1: Was true randomization used for assignment of participants to treatment groups? | - |
| | Q2: Was allocation to treatment groups concealed? | - |
| | Q3: Were treatment groups similar at the baseline? | Yes |
| Bias related to administration of intervention/exposure | Q4: Were participants blind to treatment assignment? | - |
| | Q5: Were those delivering the treatment blind to treatment assignment? | - |

| Domains | Questions in JBI tool | Questions in modified JBI tool |
|---|---|---|
| | Q6: Were treatment groups treated identically other than the intervention of interest? | Yes |
| Bias related to assessment, detection, and measurement of the outcome | Q7: Were outcome assessors blind to treatment assignment? | - |
| | Q8: Were outcomes measured in the same way for treatment groups? | Yes |
| | Q9: Were outcomes measured in a reliable way? | Yes |
| Bias related to participant retention | Q10: Was follow-up complete and, if not, were differences between groups in terms of their follow-up adequately described and analyzed? | Yes |
| Statistical conclusion validity | Q11: Were participants analyzed in the groups to which they were randomized? | Yes, with modified Q11: Were participants analyzed according to the vaccines they received?". |
| | Q12: Was appropriate statistical analysis used? | Yes |
| | Q13: Was the trial design appropriate and any deviations from the standard RCT design (individual randomization, parallel groups) accounted for in the conduct and analysis of the trial? | - |

#### b. The original and modified JBI tools for quasi experimental study

| Domains | Questions in JBI tool | Questions in modified JBI tool |
|---|---|---|
| Bias related to temporal precedence | Q1: Is it clear in the study what is the "cause" and what is the "effect" (ie, there is no confusion about which variable comes first)? | - |
| Bias related to selection and allocation | Q2: Was there a control group? | - |
| Bias related to confounding factors | Q3: Were participants included in any comparisons similar? | Yes |
| Bias related to administration of intervention/exposure | Q4: Were the participants included in any comparisons receiving similar treatment/care, other than the exposure or intervention of interest? | Yes |
| Bias related to assessment, detection, and measurement of the outcome | Q5: Were there multiple measurements of the outcome, both pre and post the intervention/exposure? | - |
| | Q6: Were the outcomes of participants included in any comparisons measured in the same way? | Yes |
| | Q7: Were outcomes measured in a reliable way? | Yes |
| Bias related to participant retention | Q8: Was follow-up complete and, if not, were differences between groups in terms of their follow-up adequately described and analyzed? | Yes |
| Statistical conclusion validity | Q9: Was appropriate statistical analysis used? | Yes |

#### c. The original and modified JBI tools for cohort study

| Domains | Questions in JBI tool | Questions in modified JBI tool |
|---|---|---|
| Bias related to selection and allocation | Q1: Were the two groups similar and recruited from the same population? | Yes |
| Bias related to classification of the exposure | Q2: Were the exposures measured similarly to assign people to both the exposed and unexposed groups? | Yes |
| | Q3: Were the exposures measured similarly to assign people to both the exposed and unexposed groups? | - |
| Bias related to confounding factors | Q4: Were confounding factors identified? | - |
| | Q5: Were strategies to deal with confounding factors stated? | - |
| Bias related to temporal precedence | Q6: Were the groups/participants free of the outcome at the start of the study (or at the moment of the exposure)? | - |
| Bias related to assessment, detection and measurement of the outcome | Q7: Were the outcomes measured in a valid and reliable way? | Yes |
| | Q8: Was the follow-up time reported and sufficient to be long enough for outcomes to occur? | Yes |
| Bias related to participant retention | Q9: Was follow up complete, and if not, were the reasons to loss to follow-up described and explored? | Yes |
| | Q10: Were strategies to address incomplete follow-up utilized? | - |
| Statistical conclusion validity | Q11: Was appropriate statistical analysis used? | Yes |

#### Supplementary Methods 9. Sensitivity analysis

Four sensitivity analyses were conducted to evaluate the robustness of the results: 1) including study arms from studies with a high risk of bias; 2) including study arms that used first-generation ELISA, GSK's 22F ELISA, ELISA without details and fluorescent multiplex immunoassay (FMIA); 3) including study arms with subcutaneous vaccine administration; 4) including study arms with late primary dosing (>4 months for first dose or >6 months for last dose).

### Supplementary Tables

#### Table S1. Summary of studies included in immunogenicity analysis

| ID | Author & Year | Study design | Vaccine type | Study arm | Setting | | | Outcome reported | | | |
|---|---|---|---|---|---|---|---|---|---|---|---|
| | | | | | Population | Country/region | Schedule | Type† | Time points* | Assay | Serotype |
| Randomized controlled trials (RCTs) | | | | | | | | | | | |
| 1 | Rennels, 1998[19] | Phase III RCT | PCV7 | 1 | Infants | United States | 3+1 (2-4-6-12 months) | GMC | post dose 2, post dose 3, post-booster | 2nd-gen ELISA | 4, 6B, 9V, 14, 18C, 19F, 23F |
| 2 | Shinefield, 1999[20] | Phase III RCT | PCV7 | 1 | Infants | United States | 3+1 (2-4-6-12 months) | GMC | post dose 3, post-booster | 2nd-gen ELISA | 4, 6B, 9V, 14, 18C, 19F, 23F |
| 3 | Black, 2000[21], US FDA Package Insert: Prevnar for Prevnar Study D118-P8[22] | Phase III RCT | PCV7 | 1 | Infants | United States | 3+1 (2-4-6-12 months) | GMC | Post dose 3, post-booster | 2nd-gen ELISA | 4, 6B, 9V, 14, 18C, 19F, 23F |
| 4 | Eskola, 2001[23], Ekström, 2005[24], Ekström, 2007[25] | Phase III RCT | PCV7 | 1 | Infants | Finland | 3+1 (2-4-6-12 months) | GMC | Post dose 3, post-booster | 1st-gen ELISA | 4, 6B, 9V, 14, 18C, 19F, 23F |
| 5 | Tichmann-Schumann, 2005[26] | Open-labeled RCT | PCV7 | 1 | Infants | Germany | 3+1 (2-3-4-13 months) | GMC | Post dose 3, post-booster | 2nd-gen ELISA | 4, 6B, 9V, 14, 18C, 19F, 23F |
| 6 | Scheifele, 2006[27], Scheifele, 2007[28] | Phase IV RCT | PCV7 | 3 | Infants | Canada | 3+1 (2-4-6-15 or 2-4-6-18 or 3-5-7-15 months) | GMC | post dose 3, post-booster | ELISA without details, conducted in Wyeth Lab using published ELISA | 4, 6B, 9V, 14, 18C, 19F, 23F |
| 7 | Knuf, 2006[29] | Open-labeled RCT | PCV7 | 1 | Infants | Germany | 3+1 (2-3-4-12~15 months) | GMC | post dose 3, post-booster | 2nd-gen ELISA | 4, 6B, 9V, 14, 18C, 19F, 23F |
| 8 | Pichichero, 2007[30] | Phase III RCT | PCV7 | 3 | Infants | United States | 3+0 (2-4-6 or 2.5-4.5-6.5 months) | GMC | Post dose 3 | 3rd-gen ELISA (WHO reference ELISA) | 4, 6B, 9V, 14, 18C, 19F, 23F |
| 9 | O'Brien, 2007[31], Millar, 2007[32] | Phase III RCT | PCV7 | 1 | Navajo and White Mountain Apache children | United States | 3+1 (2-4-6-12 months) | GMC | post dose 1, post dose 2, post dose 3, post-booster | 2nd-gen ELISA | 4, 6B, 9V, 14, 18C, 19F, 23F |
| 10 | Li, 2008[33], NCT00488826[34], Li, 2016[35] | Phase III RCT | PCV7 | 2 | Infants | China | 3+1 (3-4-5-12~15 months) | Both | post dose 3, post-booster | 3rd-gen ELISA (WHO reference ELISA) | 4, 6B, 9V, 14, 18C, 19F, 23F |
| 11 | Olivier, 2008[36] | Phase III RCT | PCV7 | 1 | Infants | France, Germany | 3+1 (2-3-4-12~15 months) | GMC | post dose 3, post-booster | 3rd-gen ELISA (WHO reference ELISA) | 4, 6B, 9V, 14, 18C, 19F, 23F |
| 12 | Dennehy, 2008[37] | Phase III RCT | PCV7 | 2 | Infants | United States | 3+1 (2-4-6-12~15 months) | GMC | Post dose 3 | 3rd-gen ELISA (WHO reference ELISA) | 4, 6B, 9V, 14, 18C, 19F, 23F |
| 13 | Trofa, 2008[38], NCT00197002[39] | Phase III RCT | PCV7 | 2 | Infants | United States | 3+1 (2-4-6-12~15 months) | GMC | Post-booster | GSK 22F-ELISA | 4, 6B, 9V, 14, 18C, 19F, 23F |
| 14 | Vesikari, 2009[40], NCT00370396[41] | Phase III RCT, PCV7 as control | PCV7 | 1 | Infants | Finland, France, Poland | 3+1 (2-3-4-12~18 months) | Both | Post dose 3, post-booster | GSK 22F-ELISA | 4, 6B, 9V, 14, 18C, 19F, 23F |

| ID | Author & Year | Study design | Vaccine type | Study arm | Setting | | | Outcome reported | | | |
|---|---|---|---|---|---|---|---|---|---|---|---|
| | | | | | Population | Country/region | Schedule | Type† | Time points* | Assay | Serotype |
| 15 | Wysocki, 2009[42] | Phase III RCT, PCV7 as control | PCV7 | 1 | Infants | Germany, Poland, Spain | 3+1 (2-4-6-11~18 months) | Both | Post dose 3, post-booster | GSK 22F-ELISA | 4, 6B, 9V, 14, 18C, 19F, 23F |
| 16 | Bermal, 2009[43], Bermal, 2011[44], NCT00344318[45], NCT00547248[46] | Phase III RCT, PCV7 as control | PCV7 | 2 | Infants | Philippines and Poland | 3+1 (6-10-14 weeks and 12~18 months for Philippines or 2-4-6-12~18 months for Poland) | Both | Post dose 3, post-booster | GSK 22F-ELISA | 4, 6B, 9V, 14, 18C, 19F, 23F |
| 17 | Givon-Lavi, 2010[47], Dagan, 2010[48], Dagan, 2012[49], Dagan, 2018[50] | Phase III RCT | PCV7 | 3 | Infants | Israel | 3+1 (2-4-6-12 months) or 3+0 (2-4-6 months) or 2+1 (4-6-12 months) | Both | Post dose 2 (for 2+1 only), post dose 3 (for 3+1 and 3+0), post-booster | 3rd-gen ELISA (WHO reference ELISA) | 4, 6B, 9V, 14, 18C, 19F, 23F |
| 18 | Wysocki, 2010[51] | Phase III RCT | PCV7 | 2 | Infants | Poland | 3+1 (2-3.5-6-12 months) | Both | Post dose 3, post-booster | 3rd-gen ELISA (WHO reference ELISA) | 4, 6B, 9V, 14, 18C, 19F, 23F |
| 19 | Goldblatt, 2010[52] | Phase IV RCT | PCV7 | 3 | Infants | United Kingdom | 2+1 (2-3-12 months) or 2+1 (2-4-12 months) | Both | Post dose 2, post-booster | 3rd-gen ELISA (WHO reference ELISA) | 4, 6B, 9V, 14, 18C, 19F, 23F |
| 20 | Grimprel, 2011[53] | Phase III RCT | PCV7 | 2 | Infants | France, Poland | 3+1 (2-3-4-12~18 months) | Both | Post dose 3, post-booster | 3rd-gen ELISA (WHO reference ELISA) | 4, 6B, 9V, 14, 18C, 19F, 23F |
| 21 | Scott, 2011[54] | Phase III RCT | PCV7 | 2 | Infants | Kenya | 3+1 (1.5-2.5-3.5-9 months) or 3+1 (0-2.5-3.5-9 months) | Both | post dose 1, post dose 2, post dose 3, post-booster | 3rd-gen ELISA (WHO reference ELISA) | 4, 6B, 9V, 14, 18C, 19F, 23F |
| 22 | van den Bergh, 2011[55], van den Bergh, 2016[56], NCT00652951[57] | Phase III RCT, PCV7 as control | PCV7 | 1 | Infants | Netherlands | 3+1 (2-3-4-11~13 months) | Both | Post dose 3, post-booster | GSK 22F-ELISA | 4, 6B, 9V, 14, 18C, 19F, 23F |
| 23 | Kim, 2011[58], NCT00680914[59], NCT00911144[60] | Phase III RCT, PCV7 as control | PCV7 | 1 | Infants | South Korea | 3+1 (2-4-6-12~18 months) | Both | Post dose 3, post-booster | GSK 22F-ELISA | 4, 6B, 9V, 14, 18C, 19F, 23F |
| 24 | Leonardi, 2011[61], NCT00109343[62] | Phase III RCT | PCV7 | 2 | Infants | United States | 3+1 (2-4-6-12~15 months) | Both | Post-booster | 3rd-gen ELISA (Merck Pn ELISA) | 4, 6B, 9V, 14, 18C, 19F, 23F |
| 25 | Marshall, 2011[63] | Phase II RCT | PCV7 | 2 | Infants | United States | 3+1 (2-4-6-12~15 months) | Both | Post dose 3, post booster | GSK 22F-ELISA | 4, 6B, 9V, 14, 18C, 19F, 23F |
| 26 | Blatter, 2012[64], NCT00578175[65] | Phase III RCT | PCV7 | 3 | Infants | United States | 3+1 (2-4-6-12~14 months) | Both | Post-booster | GSK 22F-ELISA | 4, 6B, 9V, 14, 18C, 19F, 23F |
| 27 | Klein, 2012[66], NCT00474526[67] | Phase III RCT | PCV7 | 2 | Infants | United States, Colombia, Argentina | 3+1 (2-4-6-12 months) | Both | Post dose 3 | ELISA without detail | 4, 6B, 9V, 14, 18C, 19F, 23F |
| 28 | Tapiéro, 2013[68], Halperin et al, 2014[69] | Phase II RCT | PCV7 | 2 | Infants | Canada | 3+1 (2-4-6-12 months) | GMC | Post dose 3, post-booster | 3rd-gen ELISA (Merck Pn ELISA) | 4, 6B, 9V, 14, 18C, 19F, 23F |
| 29 | Vesikari, 2013[70], NCT00657709[71] | Phase III RCT | PCV7 | 2 | Infants | Finland, Czech Republic, Germany, Austria, Italy | 3+1 (2-4-6-12 months) | GMC | Post dose 3 | ELISA without detail | 4, 6B, 9V, 14, 18C, 19F, 23F |

| ID | Author & Year | Study design | Vaccine type | Study arm | Setting | | | Outcome reported | | | |
|---|---|---|---|---|---|---|---|---|---|---|---|
| | | | | | Population | Country/region | Schedule | Type† | Time points* | Assay | Serotype |
| 30 | van Westen, 2013[72], Rodenburg, 2010[73] | Phase III RCT | PCV7 | 2 | Infants | Netherlands | 3+1 (2-3-4-11 months) or 2+1 (2-4-11 months) or 2+0 (2-4 months) | Both | Post-booster | 3rd-gen ELISA (WHO reference ELISA) | 4, 6B, 9V, 14, 18C, 19F, 23F |
| 31 | Yetman, 2013[74], NCT00312858[75] | Phase IV RCT | PCV7 | 2 | Infants | United States | 3+1 (2-4-6-12~15 months) | Both | Post-booster | 3rd-gen ELISA (Merck Pn ELISA) | 4, 6B, 9V, 14, 18C, 19F, 23F |
| 32 | Prymula, 2014[76], Esposito, 2014[77] | Phase II RCT | PCV7 | 3 | Infants | Czech Republic, Italy, Hungary, Chile, Argentina | 3+1 (2-3-4-12 months) | Both | Post dose 3 | 3rd-gen ELISA (WHO reference ELISA) | 4, 6B, 9V, 14, 18C, 19F, 23F |
| 33 | López, 2017[78], NCT01444781[79] | Phase III RCT | PCV7 | 3 | Infants | Colombia, Costa Rica | 3+1 (2-4-6-12~24 months) | Both | Post dose 3, post booster | ELISA without detail | 4, 6B, 9V, 14, 18C, 19F, 23F |
| 34 | Zhao, 2022[80] | Phase III RCT, PCV7 as control | PCV7 | 1 | Infants | China | 3+1 (3-4-5-12~15 months) | Both | Post dose 3, post-booster | 3rd-gen ELISA (WHO reference ELISA) | 4, 6B, 9V, 14, 18C, 19F, 23F |
| 35 | FDA Package Insert: Prevnar for Study D118-P16[22] | Phase II RCT | PCV7 | 1 | Infants | United States | 3+1 (2-4-6-12~15 months) | GMC | Post dose 3 | 2nd-gen ELISA | 4, 6B, 9V, 14, 18C, 19F, 23F |
| 36 | EUCTR2007-004276-39[81] | Phase III RCT | PCV7 | 2 | Infants | Germany | 3+1 (2-3-4-13 months) | Both | Post booster | ELISA without detail | 4, 6B, 9V, 14, 18C, 19F, 23F |
| 37 | NCT01250756[82] | Phase IV RCT | PCV7 (subcutaneously) | 1 | Infants | Japan | 3+1 (3-4-5-12~15 months) | Both | Post dose 3, post booster | ELISA without detail | 4, 6B, 9V, 14, 18C, 19F, 23F |
| 38 | Thisyakorn, 2014[83] | Phase III RCT | PCV7 | 1 | Infants | Thailand | 3+1 (2-4-6-12~18 months) | GMC | Post-booster | 3rd-gen ELISA (WHO reference ELISA) | 4, 6B, 9V, 14, 18C, 19F, 23F |
| 39 | Grimprel, 2011[84], NCT00366678[85], NCT01026038[86] | Phase III RCT | PCV7 & PCV13 | 2 | Infants | France | 3+1 (2-3-4-12 months) | Both | Post dose 3, post-booster | 3rd-gen ELISA (WHO reference ELISA) | 4, 6B, 9V, 14, 18C, 19F, 23F, 1, 3, 5, 6A, 7F, 19A |
| 40 | Sobanjo-ter Meulen, 2015[87], EUCTR2009-015103-58-FI[88], NCT01215175[89] | Phase I RCT | PCV7 | 1 | Infants | United States, Finland | 3+1 (2-4-6-12~15 months) | Both | Post-booster | Pn ECL | 4, 6B, 9V, 14, 18C, 19F, 23F |
| 41 | Kieninger , 2010[90], NCT00366340[91], EUCTR2005-004770-24[92] | Phase III RCT | PCV7 & PCV13 | 2 | Infants | Germany | 3+1 (2-3-4-11~12 months) | Both | Post dose 3, post-booster | 3rd-gen ELISA (WHO reference ELISA) | 4, 6B, 9V, 14, 18C, 19F, 23F, 1, 3, 5, 6A, 7F, 19A |
| 42 | Esposito, 2010[93], NCT00366899[94], EUCTR2005-004771-38-IT[95], Rodgers, 2013[96] | Phase III RCT | PCV7 & PCV13 | 2 | Infants | Italy | 2+1 (3-5-11 months) | Both | Post dose 2, post-booster | 3rd-gen ELISA (WHO reference ELISA) | 4, 6B, 9V, 14, 18C, 19F, 23F, 1, 3, 5, 6A, 7F, 19A |
| 43 | Snape, 2010[97]; Rodgers, 2013[96], NCT00384059[98], | Phase III RCT | PCV7 & PCV13 | 2 | Infants | United Kingdom | 2+1 (2-4-12 months) | Both | Post dose 2, post-booster | 3rd-gen ELISA (WHO reference ELISA) | 4, 6B, 9V, 14, 18C, 19F, 23F, |

| ID | Author & Year | Study design | Vaccine type | Study arm | Setting | | | Outcome reported | | | |
|---|---|---|---|---|---|---|---|---|---|---|---|
| | | | | | Population | Country/region | Schedule | Type† | Time points* | Assay | Serotype |
| | EUCTR2005-005130-12[99] | | | | | | | | | | 1, 3, 5, 6A, 7F, 19A |
| 44 | Bryant, 2010[100], NCT00205803[101] | Phase I and II RCT | PCV7 & PCV13 | 2 | Infants | United States | 3+1 (2-4-6-12 months) | Both | Post dose 3, post-booster | 3rd-gen ELISA (WHO reference ELISA) | 4, 6B, 9V, 14, 18C, 19F, 23F, 1, 3, 5, 6A, 7F, 19A |
| 45 | Yeh, 2010[102], NCT00373958[103] | Phase III RCT | PCV7 & PCV13 | 2 | Infants | United States | 3+1 (2-4-6-12 months) | Both | Post dose 3, post booster | 3rd-gen ELISA (WHO reference ELISA) | 4, 6B, 9V, 14, 18C, 19F, 23F, 1, 3, 5, 6A, 7F, 19A |
| 46 | Weckx, 2012[104], NCT00676091[105] | Phase III RCT | PCV7 & PCV13 | 2 | Infants | Brazil | 3+1 (2-4-6-12 months) | Both | Post dose 3, post-booster | 3rd-gen ELISA (WHO reference ELISA) | 4, 6B, 9V, 14, 18C, 19F, 23F, 1, 3, 5, 6A, 7F, 19A |
| 47 | Huang, 2012[106], NCT00688870[107] | Phase III RCT | PCV7 & PCV13 | 2 | Infants | Taiwan | 3+1 (2-4-6-15 months) | Both | Post dose 3, post-booster | 3rd-gen ELISA (WHO reference ELISA) | 4, 6B, 9V, 14, 18C, 19F, 23F, 1, 3, 5, 6A, 7F, 19A |
| 48 | Amdekar, 2013[108] | Phase III RCT | PCV7 & PCV13 | 2 | Infants | India | 3+1 (1.5-2.5-3.5-12 months) | Both | Post dose 3, post-booster | 3rd-gen ELISA (WHO reference ELISA) | 4, 6B, 9V, 14, 18C, 19F, 23F, 1, 3, 5, 6A, 7F, 19A |
| 49 | Dagan, 2013[109], Juergens, 2014[110], Dagan, 2021[111] | Phase III RCT | PCV7 & PCV13 | 2 | Infants | Israel | 3+1 (2-4-6-12 months) | GMC | Post dose 3, post booster | 3rd-gen ELISA (WHO reference ELISA) | 4, 6B, 9V, 14, 18C, 19F, 23F, 1, 3, 5, 6A, 7F, 19A |
| 50 | Kim, 2013[112], NCT00689351[113] | Phase II RCT | PCV7 & PCV13 | 2 | Infants | South Korea | 3+1 (2-4-6-12 months) | Both | Post dose 3, post booster | 3rd-gen ELISA (WHO reference ELISA) | 4, 6B, 9V, 14, 18C, 19F, 23F, 1, 3, 5, 6A, 7F, 19A |
| 51 | Rodgers, 2013[96]; Diez-Domingo, 2013[114], NCT00368966[115] | Phase III RCT | PCV7 & PCV13 | 2 | Infants | Spain | 3+1 (2-4-6-15 months) | Both | Post dose 2, post dose 3, post-booster | 3rd-gen ELISA (WHO reference ELISA) | 4, 6B, 9V, 14, 18C, 19F, 23F, 1, 3, 5, 6A, 7F, 19A |
| 52 | Payton, 2013[116], NCT00444457[117] | Phase III RCT | PCV7 & PCV13 | 4 | Infants | United States | 3+1 (2-4-6-12 months) | Both | Post dose 3, post booster | 3rd-gen ELISA (WHO reference ELISA) | 4, 6B, 9V, 14, 18C, 19F, 23F, 1, 3, 5, 6A, 7F, 19A |
| 53 | Togashi, 2015[118], NCT01200368[119] | Phase III RCT | PCV7 & PCV13 (subcuta-neously) | 2 | Infants | Japan | 3+1 (3-4-5-12~15 months) | Both | Post dose 3, post-booster | 3rd-gen ELISA (WHO reference ELISA) | 4, 6B, 9V, 14, 18C, 19F, 23F, 1, 3, 5, 6A, 7F, 19A |
| 54 | Zhu, 2016[120], Zhu, 2019[121] | Phase III RCT | PCV7 & PCV13 | 4 | Infants | China | 3+1 (2-4-6-12 months or 3-4-5-12 months) or 2+1 (3-5-12 months), | Both | Post dose 3, post booster | 3rd-gen ELISA (WHO reference ELISA) | 4, 6B, 9V, 14, 18C, 19F, 23F, 1, 3, 5, 6A, 7F, 19A |

| ID | Author & Year | Study design | Vaccine type | Study arm | Setting | | | Outcome reported | | | |
|---|---|---|---|---|---|---|---|---|---|---|---|
| | | | | | Population | Country/region | Schedule | Type† | Time points* | Assay | Serotype |
| | | | | | | | PCV7 had 3+1 (3-4-5-12 months) only | | | | |
| 55 | NCT00452790[122] | Phase III RCT | PCV7 & PCV13 | 2 | Infants | India | 3+1 (1.5-2.5-3.5-12 months) | Both | Post dose 3, post booster | 3rd-gen ELISA (WHO reference ELISA) | 4, 6B, 9V, 14, 18C, 19F, 23F, 1, 3, 5, 6A, 7F, 19A |
| 56 | Gadzinowski, 2011[123], NCT00464945[124], EUCTR2006-006204-11[125] | Phase III RCT | PCV13 | 2 | Infants | Poland | 3+1 (2-3-4-12 months) | Both | Post dose 3 | 3rd-gen ELISA (WHO reference ELISA) | 4, 6B, 9V, 14, 18C, 19F, 23F, 1, 3, 5, 6A, 7F, 19A |
| 57 | Vanderkooi, 2012[126], NCT00475033[127] | Phase III RCT | PCV13 | 1 | Infants | Canada | 3+1 (2-4-6-12 months) | Both | Post dose 3, post-booster | 3rd-gen ELISA (WHO reference ELISA) | 4, 6B, 9V, 14, 18C, 19F, 23F, 1, 3, 5, 6A, 7F, 19A |
| 58 | Spijkerman, 2013[128], van Westen, 2018[129] | Phase IV RCT | PCV13 | 4 | Infants | Netherlands | 3+1 (2-4-6-11.5 months or 2-3-4-11.5 months) or 2+1 (3-5-11.5 months or 2-4-11.5 months) | Both | Post dose 2, post dose 3, post-booster | FMIA | 4, 6B, 9V, 14, 18C, 19F, 23F, 1, 3, 5, 6A, 7F, 19A |
| 59 | Rodgers, 2013[96], Martinón-Torres, 2012[130], NCT00474539[131], EUCTR2007-000304-32[132] | Phase III RCT | PCV13 | 1 | Infants | Spain | 3+1 (2-4-6-15 months) | Both | Post dose 2, post dose 3, post-booster | 3rd-gen ELISA (WHO reference ELISA) | 4, 6B, 9V, 14, 18C, 19F, 23F, 1, 3, 5, 6A, 7F, 19A |
| 60 | Gadzinowski, 2015[133], NCT00366548[134] | Phase III RCT | PCV13 | 1 | Infants | Poland | 3+1 (2-3-4-12 months) | Both | Post dose 3, post-booster | 3rd-gen ELISA (WHO reference ELISA) | 4, 6B, 9V, 14, 18C, 19F, 23F, 1, 3, 5, 6A, 7F, 19A |
| 61 | Iro, 2015[135] | Phase IV RCT | PCV13 | 2 | Infants | United Kingdom, Malta | 2+1 (2-4-12 months) | Both | Post dose 2, post-booster | FMIA | 4, 6B, 9V, 14, 18C, 19F, 23F, 1, 3, 5, 6A, 7F, 19A |
| 62 | Truck, 2016[136] | Phase III RCT | PCV13 | 1 | Infants | United Kingdom | 2+1 (2-4-12 months) | Both | Post-booster | 3rd-gen ELISA (WHO reference ELISA) | 4, 6B, 9V, 14, 18C, 19F, 23F, 1, 3, 5, 6A, 7F, 19A |
| 63 | Block, 2016[137] | Phase III RCT | PCV13 | 3 | Infants | United States, Canada | 3+1 (2-4-6-12 months) | Both | Post dose 3, post-booster | 3rd-gen ELISA (WHO reference ELISA) | 4, 6B, 9V, 14, 18C, 19F, 23F, 1, 3, 5, 6A, 7F, 19A |
| 64 | Prymula, 2017[138], NCT01204658[139] | Phase II RCT, PCV13 as control | PCV13 | 1 | Infants | Czech Republic, Germany, Poland, Sweden | 3+1 (2-3-4-12~15 months) | Both | Post dose 3, post-booster | GSK 22F-ELISA | 4, 6B, 9V, 14, 18C, 19F, 23F, |

| ID | Author & Year | Study design | Vaccine type | Study arm | Setting | | | Outcome reported | | | |
|---|---|---|---|---|---|---|---|---|---|---|---|
| | | | | | Population | Country/region | Schedule | Type† | Time points* | Assay | Serotype |
| | | | | | | | | | | | 1, 3, 5, 6A, 7F, 19A |
| 65 | Vesikari, 2017[140], NCT01248884[141], NCT01453998[141] | Phase III RCT | PCV13 | 3 | Infants | Dominican Republic, Finland | 3+1 (2-3-4-12~15 months) | Both | Post dose 3, post-booster | GSK 22F-ELISA | 4, 6B, 9V, 14, 18C, 19F, 23F, 1, 3, 5, 6A, 7F, 19A |
| 66 | Idoko, 2017[142], NCT01964716[143], EUCTR2012-000482-21[144] | Phase III RCT | PCV13 & PCV13 (multi-dose vials) | 2 | Infants | Gambia | 3+0 (2-3-4 months) | Both | Post dose 3 | 3rd-gen ELISA (WHO reference ELISA) | 4, 6B, 9V, 14, 18C, 19F, 23F, 1, 3, 5, 6A, 7F, 19A |
| 67 | Wysocki, 2017[145], NCT01392378[146], EUCTR2010-022303-22[147] | Phase IV RCT | PCV13 | 1 | Infants | Poland | 3+1 (2-3-4-12 months) | Both | Post dose 3, post booster | 3rd-gen ELISA (WHO reference ELISA) | 4, 6B, 9V, 14, 18C, 19F, 23F, 1, 3, 5, 6A, 7F, 19A |
| 68 | Cutland, 2018[148], NCT01939158[149] | Phase III RCT | PCV13 | 2 | Infants | Australia, Canada, Czech Republic, Panama, South Africa, Turkey | 3+1 (2-4-6-12~14 months) | Both | Post-booster | 3rd-gen ELISA (WHO reference ELISA) | 4, 6B, 9V, 14, 18C, 19F, 23F, 1, 3, 5, 6A, 7F, 19A |
| 69 | Prymula, 2018[150], EUCTR2012-001055-39[151], EUCTR2012-001042-18-ES[152] | Phase III RCT | PCV13 | 2 | Infants | Germany, Czech Republic | 3+1 (2-3-4-11~15 months) | Both | Post dose 3, post-booster | 3rd-gen ELISA (WHO reference ELISA) | 4, 6B, 9V, 14, 18C, 19F, 23F, 1, 3, 5, 6A, 7F, 19A |
| 70 | Goldblatt, 2018[153], EUCTR2015-000817-32[154] | Phase II RCT | PCV13 | 2 | Infants | United Kingdom | 2+1 (2-4-12 months) or 1+1 (3-12 months) | Both | Post dose 2, post-booster | 3rd-gen ELISA (WHO reference ELISA) | 4, 6B, 9V, 14, 18C, 19F, 23F, 1, 3, 5, 6A, 7F, 19A |
| 71 | Temple, 2019[155] | Phase II and III RCT | PCV13 | 1 | Infants | Vietnam | 2+1 (2-4-9.5 months) | Both | Post dose 1, post dose 2, post-booster | 3rd-gen ELISA (WHO reference ELISA) | 4, 6B, 9V, 14, 18C, 19F, 23F, 1, 3, 5, 6A, 7F, 19A |
| 72 | Moïsi, 2019[156] | Phase IV RCT | PCV13 | 2 | Infants | Burkina Faso | 2+1 (1.5-3.5-9 months) or 3+0 (1.5-2.5-3.5 months) | Both | Post dose 2, post-booster or post dose 3 | 3rd-gen ELISA (WHO reference ELISA) | 4, 6B, 9V, 14, 18C, 19F, 23F, 1, 3, 5, 6A, 7F, 19A |
| 73 | Carmona Martinez, 2019[157], NCT01616459[158] | Phase II RCT, PCV13 as control | PCV13 | 1 | Infants | Czech Republic, Germany, Poland, Spain | 3+1 (2-3-4-12~15 months) | Both | Post dose 3, post-booster | GSK 22F-ELISA | 4, 6B, 9V, 14, 18C, 19F, 23F, 1, 3, 5, 6A, 7F, 19A |
| 74 | Odutola, 2019[159], NCT01262872[160] | Phase II RCT, PCV13 as control | PCV13 | 1 | Infants | Gambia | 3+0 (2-3-4 months) | Both | Post dose 3 | GSK 22F-ELISA | 4, 6B, 9V, 14, 18C, 19F, 23F, 1, 3, 5, 6A, 7F, 19A |
| 75 | Klein, 2019[161], NCT01978093[162] | Phase III RCT, co-administered with Hib-MenCY-TT | PCV13 | 2 | Infants | United States | 3+1 (2-4-6-12~15 months) | Both | Post dose 3, post-booster | 3rd-gen ELISA (WHO reference ELISA) | 4, 6B, 9V, 14, 18C, 19F, 23F, 1, 3, 5, 6A, 7F, 19A |

| ID | Author & Year | Study design | Vaccine type | Study arm | Setting | | | Outcome reported | | | |
|---|---|---|---|---|---|---|---|---|---|---|---|
| | | | | | Population | Country/region | Schedule | Type† | Time points* | Assay | Serotype |
| 76 | Madhi, 2020[163], Mutsaerts, 2024[164] | Phase III RCT | PCV13 | 3 | Infants | South Africa | 2+1 (1.5-3.5-9 months) or 1+1 (3.5-9 months or 1.5-9 months) | Both | Post dose 1, post dose 2, post-booster | 3rd-gen ELISA (WHO reference ELISA with 007sp) | 4, 6B, 9V, 14, 18C, 19F, 23F, 1, 3, 5, 6A, 7F, 19A |
| 77 | Shin, 2020[165] | Phase II RCT | PCV13 | 1 | Infants | Thailand | 2+1 (2-4-10~13 months) | Both | Post dose 2, post-booster | 3rd-gen ELISA (WHO reference ELISA) | 4, 6B, 9V, 14, 18C, 19F, 23F, 1, 3, 5, 6A, 7F, 19A |
| 78 | Leach, 2021[166], Leach, 2022[167] | Phase IV RCT | PCV13 | 1 | Australian Aboriginal infants | Australia | 3+0 (2-4-6 months) | Both | Post dose 3 | 3rd-gen ELISA (WHO reference ELISA) | 4, 6B, 9V, 14, 18C, 19F, 23F, 1, 3, 5, 6A, 7F, 19A |
| 79 | Lalwani, 2021[168], NCT03548337[169] | Phase IV RCT | PCV13, PCV13 (multi-dose vials) | 2 | Infants | India | 3+1 (6-10-14 weeks-12 months) | Both | Post dose 3, post-booster | Pfizer dLIA | 4, 6B, 9V, 14, 18C, 19F, 23F, 1, 3, 5, 6A, 7F, 19A |
| 80 | Dhingra, 2021[170], NCT03205371[171] | Phase III RCT, co-administered with Hib-MenCY-TT | PCV13 | 2 | Infants | the Russian Federation | 2+1 (2-4-15~23 months) | Both | Post-booster | Pn ECL | 4, 6B, 9V, 14, 18C, 19F, 23F, 1, 3, 5, 6A, 7F, 19A |
| 81 | Wang, 2022[172] | Phase III RCT | PCV13 | 1 | Infants | China | 3+1 (2-4-6-13 months) | Both | Post dose 3, post-booster | 3rd-gen ELISA (WHO reference ELISA) | 4, 6B, 9V, 14, 18C, 19F, 23F, 1, 3, 5, 6A, 7F, 19A |
| 82 | Kawade, 2023[173] | Phase IV RCT | PCV13 | 3 | Infants | India | 1+1 (3.5-9 months) or 2+1 (1.5-3.5-9 months) or 3+0 (1.5-2.5-3.5 months) | Both | Post dose 1, post dose 2, post dose 3, post-booster | 3rd-gen ELISA (WHO reference ELISA) | 4, 6B, 9V, 14, 18C, 19F, 23F, 1, 3, 5, 6A, 7F, 19A |
| 83 | Sanchez, 2023[174] | Phase III RCT | PCV13 | 2 | Infants | Thailand | 3+1 (2-4-6-15~18 months) | Both | Post dose 3 | Pn ECL | 4, 6B, 9V, 14, 18C, 19F, 23F, 1, 3, 5, 6A, 7F, 19A |
| 84 | Rajan, 2023[175] | Phase III RCT | PCV13 | 2 | Infants | United Kingdom | 1+1 (3-12 months) | Both | Post dose 1, post booster | 3rd-gen ELISA (WHO reference ELISA) | 4, 6B, 9V, 14, 18C, 19F, 23F, 1, 3, 5, 6A, 7F, 19A |
| 85 | Simon, 2023[176], NCT03550313[177] | Phase II RCT of a complementary 7-valent PCV, PCV13 as control | PCV13 | 1 | Infants | United States | 3+1 (2-4-6-12 months) | Both | Post dose 3, post-booster | Pfizer dLIA | 4, 6B, 9V, 14, 18C, 19F, 23F, 1, 3, 5, 6A, 7F, 19A |
| 86 | Temple, 2023[178] | Phase II and III RCT | PCV13 | 2 | Infants | Vietnam | 0+1 (12 months) or 1+1 (2-12 months) | Both | Post dose 1, post-booster | 3rd-gen ELISA (WHO reference ELISA) | 4, 6B, 9V, 14, 18C, 19F, 23F, 1, 3, 5, 6A, 7F, 19A |

| ID | Author & Year | Study design | Vaccine type | Study arm | Setting | | | Outcome reported | | | |
|---|---|---|---|---|---|---|---|---|---|---|---|
| | | | | | Population | Country/region | Schedule | Type† | Time points* | Assay | Serotype |
| 87 | Xie, 2024[179] | Phase III RCT, PCV13 as control | PCV13 | 1 | Infants | China | 3+1 (2-4-6-12~15 months) | Both | Post dose 3, post-booster | 3rd-gen ELISA (WHO reference ELISA) | 4, 6B, 9V, 14, 18C, 19F, 23F, 1, 3, 5, 6A, 7F, 19A |
| 88 | Matur, 2024[180] | Phase III RCT, PCV13 as control | PCV13 | 1 | Infants | India | 3+0 (1.5-2.5-3.5 months) | Rate | Post dose 3 | 2nd-gen ELISA (WHO reference ELISA with a minor modification) | 4, 6B, 9V, 14, 18C, 19F, 23F, 1, 3, 5, 7F, 19A (without 6A) |
| 89 | Gallagher, 2024[181] | Phase IV RCT | PCV13 | 1 | Infants | Kenya | 2+1 (1.5-3.5-9~12 months) | Rate | Post dose 2 | 3rd-gen ELISA (WHO reference ELISA) | 4, 6B, 9V, 14, 18C, 19F, 23F, 1, 3, 5, 6A, 7F, 19A |
| 90 | Borys, 2024[182] | Phase I RCT, PCV13 as control | PCV13 | 1 | Infants | United States | 3+1 (2-4-6-12 months) | Rate | Post-booster | Pfizer dLIA | 4, 6B, 9V, 14, 18C, 19F, 23F, 1, 3, 5, 6A, 7F, 19A |
| 91 | NCT01090453[183] | Phase II RCT, co-administered with DTPa-IPV/Hib-MenC-TT | PCV13 | 2 | Infants | Canada, France, Germany | 2+1 (2-4-12 months) | Both | Post dose 2, post booster | GSK 22F-ELISA | 4, 6B, 9V, 14, 18C, 19F, 23F, 1, 3, 5, 6A, 7F, 19A |
| 92 | NCT03207750[184] | Phase III RCT, co-administered with human rotavirus vaccine | PCV13 | 2 | Infants | United States | 3+0 (2-4-6 months) | GMC | Post dose 3 | 3rd-gen ELISA (WHO reference ELISA) | 4, 6B, 9V, 14, 18C, 19F, 23F, 1, 3, 5, 6A, 7F, 19A |
| 93 | Bili, 2023[185], NCT03620162[186], EUCTR2018-001151-12[187] | Phase III RCT, PCV13 as control | PCV13 & PCV15 | 2 | Infants | USA, Puerto Rico, Thailand, Turkey | 3+1 (2-4-6-12~15 months) | Both | Post dose 3, post booster | Pn ECL | 4, 6B, 9V, 14, 18C, 19F, 23F, 1, 3, 5, 6A, 7F, 19A, 22F, 33F |
| 94 | NCT05408429[188], EUCTR2021-006624-41[189] | Phase III RCT, PCV13 as control | PCV13 | 1 | Infants | Hungary, Poland, Spain | 2+1 (2-4-12 months) | Both | Post booster | Pfizer dLIA | 4, 6B, 9V, 14, 18C, 19F, 23F, 1, 3, 5, 6A, 7F, 19A |
| 95 | Greenberg, 2018[190], NCT01215188[191], EUCTR2010-019775-29[192] | Phase II RCT, PCV13 as control | PCV13 & PCV15 | 2 | Infants | United States, Canada, Finland, Israel, and Spain | 3+1 (2-4-6-12 months) | GMC | Post dose 3, post-booster | Pn ECL | 4, 6B, 9V, 14, 18C, 19F, 23F, 1, 3, 5, 6A, 7F, 19A, 22F, 33F |
| 96 | Rupp, 2019[193], NCT02531373[194] | Phase I and II RCT, PCV13 as control | PCV13 & PCV15 | 3 | Infants | United States | 3+1 (2-4-6-12~15 months) | Both | Post dose 3, post booster | Pn ECL | 4, 6B, 9V, 14, 18C, 19F, 23F, 1, 3, 5, 6A, 7F, 19A, 22F, 33F |
| 97 | Platt, 2020[195], NCT02987972[196] | Phase II RCT, PCV13 as control | PCV13 & PCV15 | 3 | Infants | Finland, Spain, Israel, Denmark, Canada, the United States | 3+1 (2-4-6-12~15 months) | Both | Post dose 3, post booster | Pn ECL | 4, 6B, 9V, 14, 18C, 19F, 23F, 1, 3, 5, 6A, 7F, 19A, 22F, 33F |

| ID | Author & Year | Study design | Vaccine type | Study arm | Setting | | | Outcome reported | | | |
|---|---|---|---|---|---|---|---|---|---|---|---|
| | | | | | Population | Country/region | Schedule | Type† | Time points* | Assay | Serotype |
| 98 | Banniettis, 2022[197], EUCTR2018-003706-88[198], NCT03885934[199] | Phase III RCT, PCV13 as control | PCV13 & PCV15 | 2 | Infants | Finland, Malaysia, Poland, the Russian Federation, Thailand | 2+1 (9-10-12 months) | GMC | Post booster | Pn ECL | 4, 6B, 9V, 14, 18C, 19F, 23F, 1, 3, 5, 6A, 7F, 19A, 22F, 33F |
| 99 | Martinón-Torres, 2023[200], NCT04031846[201] | Phase III RCT, PCV13 as control | PCV13 & PCV15 | 2 | Infants | Australia, Belgium, Czech Republic, Estonia, Germany, Greece, Poland, Russian Federation, Spain | 2+1 (2-4-11~15 months) | Both | Post dose 2, post booster | Pn ECL | 4, 6B, 9V, 14, 18C, 19F, 23F, 1, 3, 5, 6A, 7F, 19A, 22F, 33F |
| 100 | Benfield, 2023[202], NCT04016714[203] | Phase III RCT, PCV13 as control | PCV13 & PCV15 | 2 | Infants | Denmark, Finland, Italy, Norway | 2+1 (3-5-12 months) | Both | Post dose 2, post-booster | Pn ECL | 4, 6B, 9V, 14, 18C, 19F, 23F, 1, 3, 5, 6A, 7F, 19A, 22F, 33F |
| 101 | Suzuki, 2023[204], NCT04384107[205] | Phase III RCT, PCV13 as control | PCV13 & PCV15 (subcuta-neously) | 2 | Infants | Japan | 3+1 (2-3-4-12~15 months) | Both | Post dose 3, post-booster | Pn ECL | 4, 6B, 9V, 14, 18C, 19F, 23F, 1, 3, 5, 6A, 7F, 19A, 22F, 33F |
| 102 | Ishihara, 2023[206], Wan, 2024[207], NCT03848065[208] | Phase I RCT, PCV13 as control | PCV13 (subcuta-neously) & PCV15 (both subcuta-neously and intramu-scularly) | 3 | Infants | Japan | 3+1 (3-4-5-12~15 months) | Both | Post dose 3, post-booster | Pn ECL | 4, 6B, 9V, 14, 18C, 19F, 23F, 1, 3, 5, 6A, 7F, 19A, 22F, 33F |
| 103 | Lupinacci, 2023[209], NCT03893448[210] | Phase III RCT, PCV13 as control | PCV13 & PCV15 | 2 | Infants | United States, Puerto Rico, Thailand, Turkey | 3+1 (2-4-6-15~18 months) | Both | Post dose 3, post booster | Pn ECL | 4, 6B, 9V, 14, 18C, 19F, 23F, 1, 3, 5, 6A, 7F, 19A, 22F, 33F |
| 104 | Senders, 2021[211], NCT03512288[212] | Phase II RCT, PCV13 as control | PCV13 & PCV20 | 2 | Infants | United States | 3+1 (2-4-6-12 months) | GMC | Post dose 3, post-booster | Pfizer dLIA | 4, 6B, 9V, 14, 18C, 19F, 23F, 1, 3, 5, 6A, 7F, 19A, 22F, 33F, 8, 10A, 11A, 12F,15B |
| 105 | Korbal, 2024[213], NCT04546425[214] | Phase III RCT, PCV13 as control | PCV13 & PCV20 | 2 | Infants | Australia, Czechia, Denmark, Estonia, Finland, Italy, Netherlands, Norway, Poland, Russian Federation, Slovakia | 2+1 (2-4-12 months) | Both | Post dose 2, post-booster | Pfizer dLIA | 4, 6B, 9V, 14, 18C, 19F, 23F, 1, 3, 5, 6A, 7F, 19A, 22F, 33F, 8, 10A, 11A, 12F,15B |
| 106 | Ishihara, 2024[215], NCT04530838[216] | Phase III RCT, PCV13 as control | PCV13 (subcuta-neously) & PCV20 (both | 3 | Infants | Japan | 3+1 (2-3-4-12~15 months) | Both | Post dose 3, post booster | Pfizer dLIA | 4, 6B, 9V, 14, 18C, 19F, 23F, 1, 3, 5, 6A, 7F, 19A, 22F, 33F, |

| ID | Author & Year | Study design | Vaccine type | Study arm | Setting | | | Outcome reported | | | |
|---|---|---|---|---|---|---|---|---|---|---|---|
| | | | | | Population | Country/region | Schedule | Type† | Time points* | Assay | Serotype |
| | | | subcuta-neously and intramu-scularly) | | | | | | | | 8, 10A, 11A, 12F,15B |
| 107 | Senders, 2024[217], NCT04382326[218], EUCTR2019-003305-10[219] | Phase III RCT, PCV13 as control | PCV13 & PCV20 | 2 | Infants | United States, Puerto Rico | 3+1 (2-4-6-12~15 months) | Both | Post dose 3, post booster | Pfizer dLIA | 4, 6B, 9V, 14, 18C, 19F, 23F, 1, 3, 5, 6A, 7F, 19A, 22F, 33F, 8, 10A, 11A, 12F,15B |
| 108 | Clarke, 2020[220], NCT02308540[221] | Phase I and II RCT, PCV13 as control | PCV10-SII & PCV13 | 3 | Infants | Gambia | 3+1 (2-3-4-10 months or 2-3-4-10~14 months) | Both | Post dose 3, post-booster | 3rd-gen ELISA (WHO reference ELISA) | 1, 5, 6A, 6B, 7F, 9V, 14, 19A, 19F, 23F |
| 109 | Clarke, 2021[222], NCT03197376[223] | Phase III RCT | PCV10-SII | 1 | Infants | Gambia | 3+1 (1.5-2.5-3.5-9 months) | Both | Post dose 3, post-booster | 3rd-gen ELISA (WHO reference ELISA) | 1, 5, 6A, 6B, 7F, 9V, 14, 19A, 19F, 23F |
| 110 | Adigweme, 2023[224], NCT03896477[225] | Phase III RCT | PCV10-SII & PCV13 | 2 | Infants | Gambia | 2+1 (1.5-3.5-9~18 months) | Both | Post dose 2, post-booster | 3rd-gen ELISA (WHO reference ELISA) | 1, 5, 6A, 6B, 7F, 9V, 14, 19A, 19F, 23F |
| Quasi-experimental studies | | | | | | | | | | | |
| 111 | Nurkka, 2001[226] | Phase II study, non-randomized study | PCV7 and PCV7 as primary PPSV23 as booster | 1 | Infants | Finland | 3+1 (2-4-6-15 months) | GMC | Post dose 3, post-booster | 1st-gen ELISA | 4, 6B, 9V, 14, 18C, 19F, 23F |
| 112 | Käyhty, 2005[227] | Single-arm, non-randomized study | PCV7 | 1 | Infants | Sweden | 2+1 (3-5-12 months) | GMC | Post dose 2, post-booster | 2nd-gen ELISA | 4, 6B, 9V, 14, 18C, 19F, 23F |
| 113 | Shao, 2006[228] | Single-arm, non-experimental study | PCV7 | 1 | Infants | Taiwan | 3+1 (2-4-6-15~20 months) | GMC | Post-booster | 2nd-gen ELISA | 4, 6B, 9V, 14, 18C, 19F, 23F |
| 114 | Kim, 2007[229] | Single-arm, non-experimental study | PCV7 | 1 | Infants | South Korea | 3+1 (2-4-6-12 months) | Both | Post dose 2, post dose 3 | 2nd-gen ELISA | 4, 6B, 9V, 14, 18C, 19F, 23F |
| 115 | Lee, 2009[230] | Single-arm, non-randomized study | PCV7 | 1 | Infants who monitored at Ewha Winans University Hospital | Korea | 3+0 (2-4-6 months) | Both | Post dose 3 | 3rd-gen ELISA (WHO reference ELISA) | 4, 6B, 9V, 14, 18C, 19F, 23F |
| 116 | Li, 2015[231] | Phase IV trial, open label, grouping based on age, single arm per age group | PCV7 | 2 | Infants | China | 3+1 (5-6-7-14 months) or 2+1 (10-11-15 months) | Both | Post-booster | 3rd-gen ELISA (WHO reference ELISA) | 4, 6B, 9V, 14, 18C, 19F, 23F |
| 117 | Togashi, 2013[232], NCT00574795[233] | Single-arm, non-experimental, open label study | PCV13 (subcuta-neously) | 1 | Infants | Japan | 3+1 (3-4-5-12~15 months) | Both | Post dose 3, post-booster | 3rd-gen ELISA (WHO reference ELISA) | 4, 6B, 9V, 14, 18C, 19F, 23F, 1, 3, 5, 6A, 7F, 19A |

| ID | Author & Year | Study design | Vaccine type | Study arm | Setting | | | Outcome reported | | | |
|---|---|---|---|---|---|---|---|---|---|---|---|
| | | | | | Population | Country/region | Schedule | Type† | Time points* | Assay | Serotype |
| 118 | Rodgers, 2013[96], Gutiérrez Brito, 2013[234], NCT00708682[235] | Single-arm, open-label study | PCV13 | 1 | Infants | Mexico | 3+1 (2-4-6-12 months) | Both | Post dose 2, post dose 3, post-booster | 3rd-gen ELISA (WHO reference ELISA) | 4, 6B, 9V, 14, 18C, 19F, 23F, 1, 3, 5, 6A, 7F, 19A |
| 119 | Singleton, 2013[236] | Phase III trial, open label, grouping based on age, single arm per age group | PCV13 | 1 | Alaska Native children | United States | 3+1 (2-4-6-12 months) | Rate | Post dose 3 | 3rd-gen ELISA (WHO reference ELISA) | 4, 6B, 9V, 14, 18C, 19F, 23F, 1, 3, 5, 6A, 7F, 19A |
| 120 | Wijmenga-Monsuur, 2015[237], van Westen, 2015[238], van Westen, 2018[129] | Non-randomized study phase IV trial | PCV13 | 1 | Infants | Netherlands | 3+1 (2-3-4-11 months) | Both | Post-booster | FMIA | 4, 6B, 9V, 14, 18C, 19F, 23F, 1, 3, 5, 6A, 7F, 19A |
| 121 | Martinón-Torres, 2015[239], Martinón-Torres, 2017[240], NCT01193335[241] | Phase IV trial, open label, grouping based on GA at birth, single arm per group | PCV13 | 1 | Infants | Poland, Spain | 3+1 (2-3-4-12 months) | Both | Post dose 3, post booster | 3rd-gen ELISA (WHO reference ELISA) | 4, 6B, 9V, 14, 18C, 19F, 23F, 1, 3, 5, 6A, 7F, 19A |
| 122 | Chu, 2023[242], NCT03574389[243] | Phase III trial, single-arm, open-label | PCV13 | 2 | Infants | China | 3+1 (2-4-6-12~15 months) or 2+1 (10-11-14 months) | Both | Post dose 3, post booster | Not mentioned | 4, 6B, 9V, 14, 18C, 19F, 23F, 1, 3, 5, 6A, 7F, 19A |
| 123 | Urbancikova, 2017[244] | Phase III trial, open label, grouping based on vaccine history | PCV13 | 2 | Infants | Czech Republic | 3+1 (2-3-4-12~15 months) or 2+1 (2-4-11~12 months) | Both | Post-booster | 3rd-gen ELISA (WHO reference ELISA) | 4, 6B, 9V, 14, 18C, 19F, 23F, 1, 3, 5, 6A, 7F, 19A |
| 124 | Maestri, 2024[245], NCT04633226[246] | Single-arm phase III trial | PCV15 | 1 | Infants | Korea | 3+1 (2-4-6-12~15 months) | Both | Post dose 3, post booster | Pn ECL | 4, 6B, 9V, 14, 18C, 19F, 23F, 1, 3, 5, 6A, 7F, 19A, 22F, 33F |
| Cohort studies | | | | | | | | | | | |
| 125 | O'Brien, 2000[247] | Prospective cohort study | PCV7 | 1 | SCD and matched healthy infants (SCD excluded) | United States | 3+0 (2-4-6 months) | GMC | post dose 3 | 2nd-gen ELISA | 4, 6B, 9V, 14, 18C, 19F, 23F |
| 126 | Esposito, 2005[248] | Prospective cohort study nested within a cluster-RCT | PCV7 | 1 | Preterm and term infants | Italy | 2+1 (3-5-11 months) | Rate | Post dose 2, post-booster | 2nd-gen ELISA | 4, 6B, 9V, 14, 18C, 19F, 23F |
| 127 | Osendarp, 2007[249] | Prospective cohort study nested within a cluster-RCT | PCV7 | 1 | Infants with or without zinc supplementation | Bangladesh | 3+0 (4-6-7 months) | GMC | Post dose 1, post dose 2, post dose 3 | 2nd-gen ELISA | 4, 6B, 9V, 14, 18C, 19F, 23F |
| 128 | Vesikari, 2010[250] | Prospective cohort study nested within a RCT | PCV7 | 2 | Infants | France, Germany | 3+1 (2-3-4/12-15 months) | Both | Post dose 3 | 3rd-gen ELISA (WHO reference ELISA) | 4, 6B, 9V, 14, 18C, 19F, 23F |

| ID | Author & Year | Study design | Vaccine type | Study arm | Setting | | | Outcome reported | | | |
|---|---|---|---|---|---|---|---|---|---|---|---|
| | | | | | Population | Country/region | Schedule | Type† | Time points* | Assay | Serotype |
| 129 | Moss, 2010[251], Moss, 2010[252] | Prospective cohort study | PCV7 | 2 | Healthy term infants | United Kingdom | 3+0 (2-3-4 months) or 3+1 (2-3-4-12 months) | Both | Post dose 3, post-booster | 3rd-gen ELISA (WHO reference ELISA) | 4, 6B, 9V, 14, 18C, 19F, 23F |
| 130 | Whelan, 2012[253] | Prospective cohort study | PCV7 | 1 | Children who were already receiving routine vaccinations as part of the national immunization schedule | Netherlands | 3+1 (2-3-4-11 months) | Both | Post-booster | 3rd-gen ELISA (WHO reference ELISA) | 4, 6B, 9V, 14, 18C, 19F, 23F |
| 131 | Jones, 2013[254] | Prospective cohort study | PCV7 | 1 | Healthy, HIV-unexposed infants | South Africa | 2+1 (1.5-3.5-9 months) | Both | Post dose 1, post dose 2, post-booster | 3rd-gen ELISA (WHO reference ELISA) | 4, 6B, 9V, 14, 18C, 19F, 23F |
| 132 | Madhi, 2013[255], Madhi et all. 2010[256], Madhi, 2020[257] | Prospective cohort study | PCV7 | 1 | Infants with various HIV status and healthy infant cohorts | South Africa | 3+1 (1.5-2.5-3.5-15~18 months) | Both | Post dose 1, post dose 2, post dose 3, post-booster | 3rd-gen ELISA (WHO reference ELISA) | 4, 6B, 9V, 14, 18C, 19F, 23F |
| 133 | Ladhani, 2015[258] | Prospective cohort study | PCV13 | 1 | Infants born at term who were immunised according to the national schedule | United Kingdom | 2+1 (2-4-12 months) | Both | Post dose 2 | 3rd-gen ELISA (WHO reference ELISA) | 4, 6B, 9V, 14, 18C, 19F, 23F, 1, 3, 5, 6A, 7F, 19A |
| 134 | Ladhani, 2015[259] | Prospective cohort study | PCV13 | 1 | Infants born at term who were immunised according to the national schedule | United Kingdom | 2+1 (2-4-12 months) | Both | Post dose 2 | 3rd-gen ELISA (WHO reference ELISA) | 4, 6B, 9V, 14, 18C, 19F, 23F, 1, 3, 5, 6A, 7F, 19A |
| 135 | Maertens, 2017[260] | Prospective cohort study nested within a cluster-randomized trials | PCV13 | 2 | Infants born to Tdap vaccinated mothers and controls | Belgium | 2+1 (2-4-12 months) | Both | Post dose 2, post-booster | 3rd-gen ELISA (WHO reference ELISA) | 4, 6B, 9V, 14, 18C, 19F, 23F, 1, 3, 5, 6A, 7F, 19A |
| 136 | Madhi, 2017[261] | Prospective cohort study nested within a phase Ib and II RCT | PCV13 | 1 | Infants born to GBS vaccinated mothers and controls | South Africa | 3+1 (1.5-2.5-3.5-9 months) | Both | Post dose 3, post booster | 3rd-gen ELISA (WHO reference ELISA) | 4, 6B, 9V, 14, 18C, 19F, 23F, 1, 3, 5, 6A, 7F, 19A |
| 137 | Zimmermann, 2020[262], Zimmermann, 2019[263], Zimmermann, 2019[264] | Prospective cohort study nested within a cluster-randomized trials | PCV13 | 2 | Infants randomised to receive neonatal BCG vaccination or no intervention | Australia | 3+0 (1.5-4-6 months) | Both | Post dose 3 | FMIA | 4, 6B, 9V, 14, 18C, 19F, 23F, 1, 3, 5, 6A, 7F, 19A |
| 138 | Perrett, 2020[265], Martinón-Torres, 2021[266], NCT02422264[267], NCT02853929[268] | Prospective cohort study nested within a cluster-randomized trials | PCV13 | 4 | Infants born to Tdap vaccinated mothers and controls | Australia, Canada, Czech Republic, Finland, Italy, Spain | 3+1 (2-4-6-11~18 months) or 2+1 (3-5-11~18 months) | Both | Post dose 2, post dose 3, post-booster | Pn ECL | 4, 6B, 9V, 14, 18C, 19F, 23F, 1, 3, 5, 6A, 7F, 19A |

Note: GMC = geometric mean concentration; 1st-gen ELISA = first-generation enzyme-linked immunosorbent assay; 2nd-gen ELISA = second-generation enzyme-linked immunosorbent assay; 3rd-gen ELISA = third-generation enzyme-linked immunosorbent assay; WHO reference ELISA = World Health Organization enzyme-linked immunosorbent assay; GSK 22F-ELISA = GlaxoSmithKline 22F inhibition enzyme-linked immunosorbent assay; MIA = multiplex immunoassay; Pfizer dLIA = Pfizer direct Luminex immunoassay; Pn ECL = pneumococcal electrochemiluminescence assay.

†: "GMC" refers to IgG GMCs measured at specific time points post-vaccination; "Rate" refers to the seroresponse rate; and "Both" indicates that both outcomes are reported.

*: 30 days (4–6 weeks) post each sampling time point.

### Table S2. Variable list and completeness analysis

| Variable name | Definition/details | Type | Completeness (N = 4449)* |
|---|---|---|---|
| title | Title of publication paper(s) | String | 4449 (100%) |
| Author_year | Author and publication year of paper(s) | String | 4449 (100%) |
| study_design_type | Study design type of this study arm | String | 4449 (100%) |
| study_sponsor | Sponsor of this study | String | 4421 (99·4%) |
| study_period | The period during which the study was conducted | String | 4449 (100%) |
| country | Country name where study was conducted | String | 4449 (100%) |
| ISO3 | Three-letter country codes | String | 4449 (100%) |
| WHO_region | Region of study sites based on WHO regions. The WHO regions are focused on public health administration and epidemiology. WHO divides the world based on health challenges, needs, and capacities to better coordinate global health initiatives and responses to diseases, including African Region (AFR), Region of the Americas (AMR), South-East Asian Region (SEAR), European Region (EUR), Eastern Mediterranean Region (EMR), and Western Pacific Region (WPR). | String | 4449 (100%) |
| Income_group | The World Bank classifies economies for analytical purposes into four income groups: low, lower-middle, upper-middle, and high income. | String | 4449 (100%) |
| sample_size | The sample size for immune response for certain vaccine, schedule, serotype and sampling time | Integer | 4449 (100%) |
| study_eligibility_minimum_age | The minimum age requirement to participant in the study | String | 4449 (100%) |
| study_eligibility_maximum_age | The maximum age requirement to participant in the study | String | 4400 (98.9%) |
| whe_vaccine_naive | Whether participant is naive to PCVs or not | Boolean | 4449 (100%) |
| percentage_vaccine_naive | The percentage of vaccine naive participants | Numeric | 4449 (100%) |
| percentage_female | The percentage of female participants | Numeric | 4199 (94·4%) |
| indigenous_status | Study conducted specifically for Indigenous population or not | Boolean | 4449 (100%) |
| ethnicity_or_race | The percentage of participants identifying their ethnicity as White | Boolean | 4449 (100%) |
| ethnicity_race_proportion | The percentage of participants identifying their race / ethnicity | String | 3311 (74·4%) |
| vaccine_type | Vaccine used througout the study in order | String | 4449 (100%) |
| administration_vaccine | Route of vaccine administration, given intramuscularly, or subcutaneously | String | 4449 (100%) |
| vaccine_schedule | Vaccine schedule | String | 4449 (100%) |
| vaccine_schedule_time | Timing of scheduled dose | String | 4449 (100%) |
| age_first_dose_primary | Timing of first dose of primay series | String | 4436 (99·7%) |
| age_last_dose_primary | Timing of last dose of primay series | Numeric | 4293 (96·5%) |
| vaccine_interval_primary | Time interval between primary series | Numeric | 4293 (96·5%) |
| timing_booster | Timing of booster dose | Numeric | 4449 (100%) |
| serotype | Serotype | String | 4449 (100%) |
| sampling_time | Time points of blood sample date post vaccination to get the antibody levels | String | 4449 (100%) |
| laboratory_assay | Assays used to measure IgG antibody | String | 4449 (100%) |
| results_mean | Mean value of IgG GMCs | String | 4338 (97·5%) |
| results_lower | Lower 95% confidence intervals for the IgG GMCs | Numeric | 4326 (97·2%) |
| results_upper | Upper 95% confidence intervals for the IgG GMCs | Numeric | 4326 (97·2%) |
| seropositive_num | Number of participants with pneumococcal serotype-specific IgG concentrations higher than predefined threshold | Numeric | 3535 (79·5%) |
| seropositive_rate | Percentage of participant with pneumococcal serotype-specific IgG concentrations higher than predefined threshold | Numeric | 3535 (79·5%) |

**Table S3. Risk of bias assessment according to the modified JBI tool for randomized controlled trial (RCT)**

| ID | Author & Year | Q3 | Q6 | Q8 | Q9 | Q10 | Modified Q11 | Q12 | Overall |
|---|---|---|---|---|---|---|---|---|---|
| 1 | Rennels, 1998[19] | Yes | Yes | Yes | Yes | No | Yes | Yes | High |
| 2 | Shinefield, 1999[20] | Yes | Yes | Yes | Yes | Yes | Yes | Yes | Low |
| 3 | Black, 2000[21], US FDA Package Insert: Prevnar for Prevnar Study D118-P8[22] | Unclear | Yes | Yes | Yes | No | Yes | Yes | High |
| 4 | Eskola, 2001[23], Ekström, 2005[24], Ekström, 2007[25] | Yes | Yes | Yes | Yes | Yes | Yes | Yes | Low |
| 5 | Tichmann-Schumann, 2005[26] | Unclear | Yes | Yes | Yes | Yes | Yes | Yes | Low |
| 6 | Scheifele, 2006[27], Scheifele, 2007[28] | Yes | Yes | Yes | Unclear | Yes | Yes | Yes | High |
| 7 | Knuf, 2006[29] | Yes | Yes | Yes | Yes | Yes | Yes | Yes | Low |
| 8 | Pichichero, 2007[30] | Yes | Yes | Yes | Yes | Yes | Yes | Yes | Low |
| 9 | O'Brien, 2007[31], Millar, 2007[32] | Yes | Yes | Yes | Yes | No | Yes | Yes | High |
| 10 | Li, 2008[33], NCT00488826[34], Li, 2016[35] | Yes | Yes | Yes | Yes | Yes | Yes | Yes | Low |
| 11 | Olivier, 2008[36] | Yes | Yes | Yes | Yes | No | No | Yes | High |
| 12 | Dennehy, 2008[37] | Yes | Yes | Yes | Yes | No | Yes | Yes | High |
| 13 | Trofa, 2008[38], NCT00197002[39] | Yes | Yes | Yes | Yes | Yes | Yes | Yes | Low |
| 14 | Vesikari, 2009[40], NCT00370396[41] | Yes | Yes | Yes | Yes | Yes | Yes | Yes | Low |
| 15 | Wysocki, 2009[42] | Yes | Yes | Yes | Yes | Yes | Yes | Yes | Low |
| 16 | Bermal, 2009[43], Bermal, 2011[44], NCT00344318[45], NCT00547248[46] | Yes | Yes | Yes | Yes | Yes | Yes | Yes | Low |
| 17 | Givon-Lavi, 2010[47], Dagan, 2010[48], Dagan, 2012[49], Dagan, 2018[50] | Yes | Yes | Yes | Yes | No | Yes | Yes | Moderate |
| 18 | Wysocki, 2010[51] | Yes | Yes | Yes | Yes | Yes | Yes | Yes | Low |
| 19 | Goldblatt, 2010[52] | Yes | Yes | Yes | Yes | Yes | Yes | Yes | Low |
| 20 | Grimprel, 2011[53] | Yes | Yes | Yes | Yes | Yes | Yes | Yes | Low |
| 21 | Scott, 2011[54] | Yes | Yes | Yes | Yes | Yes | Yes | Yes | Low |
| 22 | van den Bergh, 2011[55], van den Bergh, 2016[56], NCT00652951[57] | Yes | Yes | Yes | Yes | Yes | Yes | Yes | Low |
| 23 | Kim, 2011[58], NCT00680914[59], NCT00911144[60] | Yes | Yes | Yes | Yes | Yes | Yes | Yes | Low |
| 24 | Leonardi, 2011[61], NCT00109343[62] | Yes | Yes | Yes | Yes | Yes | Yes | Yes | Low |
| 25 | Marshall, 2011[63] | Yes | Yes | Yes | Yes | No | Yes | Yes | Low |
| 26 | Blatter, 2012[64], NCT00578175[65] | Yes | Yes | Yes | Yes | Yes | Yes | Yes | Low |
| 27 | Klein, 2012[66], NCT00474526[67] | Yes | Yes | Yes | Unclear | Yes | Yes | Yes | High |
| 28 | Tapiéro, 2013[68], Halperin et al, 2014[69] | Yes | Yes | Yes | Yes | Yes | Yes | Yes | Low |
| 29 | Vesikari, 2013[70], NCT00657709[71] | Yes | Yes | Yes | Unclear | Yes | No | Yes | High |
| 30 | van Westen, 2013[72], Rodenburg, 2010[73] | Yes | Yes | Yes | Yes | Yes | Yes | Yes | Low |
| 31 | Yetman, 2013[74], NCT00312858[75] | Yes | Yes | Yes | Yes | Yes | Yes | Yes | Low |
| 32 | Prymula, 2014[76], Esposito, 2014[77] | Yes | Yes | Yes | Yes | Yes | Yes | Yes | Low |
| 33 | López, 2017[78], NCT01444781[79] | Yes | Yes | Yes | Unclear | Yes | Yes | Yes | High |
| 34 | Zhao, 2022[80] | Yes | Yes | Yes | Yes | Yes | Yes | Yes | Low |
| 35 | FDA Package Insert: Prevnar for Study D118-P16[22] | Unclear | Yes | Yes | Yes | Unclear | Yes | Unclear | High |
| 36 | EUCTR2007-004276-39[81] | Yes | Yes | Yes | Unclear | Unclear | Yes | Yes | High |
| 37 | NCT01250756[82] | Yes | Yes | Yes | Unclear | Unclear | Yes | Yes | High |
| 38 | Thisyakorn, 2014[83] | Yes | Yes | Yes | Yes | Yes | Yes | Yes | Low |
| 39 | Grimprel, 2011[84], NCT00366678[85], NCT01026038[86] | Yes | Yes | Yes | Yes | Yes | Yes | Yes | Low |
| 40 | Sobanjo-ter Meulen, 2015[87], EUCTR2009-015103-58-FI[88], NCT01215175[89] | Yes | Yes | Yes | Yes | Yes | Yes | Yes | Low |
| 41 | Kieninger , 2010[90], NCT00366340[91], EUCTR2005-004770-24[92] | Yes | Yes | Yes | Yes | Yes | Yes | Yes | Low |
| 42 | Esposito, 2010[93], NCT00366899[94], EUCTR2005-004771-38-IT[95], Rodgers, 2013[96] | Yes | Yes | Yes | Yes | Yes | Yes | Yes | Low |
| 43 | Snape, 2010[97]; Rodgers, 2013[96], NCT00384059[98], EUCTR2005-005130-12[99] | Yes | Yes | Yes | Yes | Yes | Yes | Yes | Low |
| 44 | Bryant, 2010[100], NCT00205803[101] | Yes | Yes | Yes | Yes | Yes | Yes | Yes | Low |

| ID | Author & Year | Q3 | Q6 | Q8 | Q9 | Q10 | Modified Q11 | Q12 | Overall |
|---|---|---|---|---|---|---|---|---|---|
| 45 | Yeh, 2010[102], NCT00373958[103] | Yes | Yes | Yes | Yes | Yes | Yes | Yes | Low |
| 46 | Weckx, 2012[104], NCT00676091[105] | Yes | Yes | Yes | Yes | Yes | Yes | Yes | Low |
| 47 | Huang, 2012[106], NCT00688870[107] | Yes | Yes | Yes | Yes | Yes | Yes | Yes | Low |
| 48 | Amdekar, 2013[108] | Yes | Yes | Yes | Yes | No | Yes | Yes | Moderate |
| 49 | Dagan, 2013[109], Juergens, 2014[110], Dagan, 2021[111] | Yes | Yes | Yes | Yes | Yes | Yes | Yes | Low |
| 50 | Kim, 2013[112], NCT00689351[113] | Yes | Yes | Yes | Yes | Yes | Yes | Yes | Low |
| 51 | Rodgers, 2013[96]; Diez-Domingo, 2013[114], NCT00368966[115] | Yes | Yes | Yes | Yes | Yes | Yes | Yes | Low |
| 52 | Payton, 2013[116], NCT00444457[117] | Yes | Yes | Yes | Yes | Yes | Yes | Yes | Low |
| 53 | Togashi, 2015[118], NCT01200368[119] | Yes | Yes | Yes | Yes | Yes | Yes | Yes | Low |
| 54 | Zhu, 2016[120], Zhu, 2019[121] | Yes | Yes | Yes | Yes | Yes | Yes | Yes | Low |
| 55 | NCT00452790[122] | Yes | Yes | Yes | Yes | Yes | Yes | Yes | Low |
| 56 | Gadzinowski, 2011[123], NCT00464945[124], EUCTR2006-006204-11[125] | Yes | Yes | Yes | Yes | Yes | Yes | Yes | Low |
| 57 | Vanderkooi, 2012[126], NCT00475033[127] | Yes | Yes | Yes | Yes | Yes | Yes | Yes | Low |
| 58 | Spijkerman, 2013[128], van Westen, 2018[129] | Yes | Yes | Yes | Yes | Yes | Yes | Yes | Low |
| 59 | Rodgers, 2013[96], Martinón-Torres, 2012[130], NCT00474539[131], EUCTR2007-000304-32[132] | Yes | Yes | Yes | Yes | Yes | Yes | Yes | Low |
| 60 | Gadzinowski, 2015[133], NCT00366548[134] | Yes | Yes | Yes | Yes | Yes | Yes | Yes | Low |
| 61 | Iro, 2015[135] | Yes | Yes | Yes | Yes | No | Yes | Yes | Moderate |
| 62 | Truck, 2016[136] | Yes | Yes | Yes | Yes | Yes | Yes | Yes | Low |
| 63 | Block, 2016[137] | Yes | Yes | Yes | Yes | No | Yes | Yes | Moderate |
| 64 | Prymula, 2017[138], NCT01204658[139] | Yes | Yes | Yes | Yes | Yes | Yes | Yes | Low |
| 65 | Vesikari, 2017[140], NCT01248884[141], NCT01453998[141] | Yes | Yes | Yes | Yes | Yes | Yes | Yes | Low |
| 66 | Idoko, 2017[142], NCT01964716[143], EUCTR2012-000482-21[144] | Yes | Yes | Yes | Yes | Yes | Yes | Yes | Low |
| 67 | Wysocki, 2017[145], NCT01392378[146], EUCTR2010-022303-22[147] | Yes | Yes | Yes | Yes | Yes | No | Yes | Low |
| 68 | Cutland, 2018[148], NCT01939158[149] | Yes | Yes | Yes | Yes | Yes | Yes | Yes | Low |
| 69 | Prymula, 2018[150], EUCTR2012-001055-39[151], EUCTR2012-001042-18-ES[152] | Yes | Yes | Yes | Yes | Yes | Yes | Yes | Low |
| 70 | Goldblatt, 2018[153], EUCTR2015-000817-32[154] | Yes | Yes | Yes | Yes | Yes | Yes | Yes | Low |
| 71 | Temple, 2019[155] | Yes | Yes | Yes | Yes | Yes | Yes | Yes | Low |
| 72 | Moïsi, 2019[156] | Yes | Yes | Yes | Yes | Yes | No | Yes | Low |
| 73 | Carmona Martinez, 2019[157], NCT01616459[158] | Yes | Yes | Yes | Yes | Yes | Yes | Yes | Low |
| 74 | Odutola, 2019[159], NCT01262872[160] | Yes | Yes | Yes | Yes | Yes | Yes | Yes | Low |
| 75 | Klein, 2019[161], NCT01978093[162] | No | Yes | Yes | Yes | Yes | Yes | Yes | Moderate |
| 76 | Madhi, 2020[163], Mutsaerts, 2024[164] | Yes | Yes | Yes | Yes | Yes | No | Yes | Low |
| 77 | Shin, 2020[165] | Yes | Yes | Yes | Yes | Yes | Yes | Yes | Low |
| 78 | Leach, 2021[166], Leach, 2022[167] | Yes | Yes | Yes | Yes | Yes | Yes | Yes | Low |
| 79 | Lalwani, 2021[168], NCT03548337[169] | Yes | Yes | Yes | Yes | Yes | Yes | Yes | Low |
| 80 | Dhingra, 2021[170], NCT03205371[171] | Yes | Yes | Yes | Yes | Yes | Yes | Yes | Low |
| 81 | Wang, 2022[172] | Yes | Yes | Yes | Yes | Yes | Yes | Yes | Low |
| 82 | Kawade, 2023[173] | Yes | Yes | Yes | Yes | Yes | No | Yes | Low |
| 83 | Sanchez, 2023[174] | Yes | Yes | Yes | Yes | Yes | Yes | Yes | Low |
| 84 | Rajan, 2023[175] | Yes | Yes | Yes | Yes | Yes | No | Yes | Low |
| 85 | Simon, 2023[176], NCT03550313[177] | Yes | Yes | Yes | Yes | Yes | Yes | Yes | Low |
| 86 | Temple, 2023[178] | Yes | Yes | Yes | Yes | Yes | No | Yes | Low |
| 87 | Xie, 2024[179] | Yes | Yes | Yes | Yes | Yes | Yes | Yes | Low |
| 88 | Matur, 2024[180] | Yes | Yes | Yes | Yes | Yes | Yes | Yes | Low |
| 89 | Gallagher, 2024[181] | Yes | Yes | Yes | Yes | Yes | Yes | Yes | Low |
| 90 | Borys, 2024[182] | No | Yes | Yes | Yes | Yes | Yes | Yes | Moderate |

| ID | Author & Year | Q3 | Q6 | Q8 | Q9 | Q10 | Modified Q11 | Q12 | Overall |
|---|---|---|---|---|---|---|---|---|---|
| 91 | NCT01090453[183] | Yes | Yes | Yes | Yes | Yes | Yes | Yes | Low |
| 92 | NCT03207750[184] | Yes | Yes | Yes | Yes | Yes | Yes | Yes | Low |
| 93 | Bili, 2023[185], NCT03620162[186], EUCTR2018-001151-12[187] | Yes | Yes | Yes | Yes | Yes | Yes | Yes | Low |
| 94 | NCT05408429[188], EUCTR2021-006624-41[189] | Yes | Yes | Yes | Yes | Yes | Yes | Yes | Low |
| 95 | Greenberg, 2018[190], NCT01215188[191], EUCTR2010-019775-29[192] | Yes | Yes | Yes | Yes | Yes | Yes | Yes | Low |
| 96 | Rupp, 2019[193], NCT02531373[194] | No | Yes | Yes | Yes | No | Yes | Yes | High |
| 97 | Platt, 2020[195], NCT02987972[196] | Yes | Yes | Yes | Yes | Yes | Yes | Yes | Low |
| 98 | Banniettis, 2022[197], EUCTR2018-003706-88[198], NCT03885934[199] | Yes | Yes | Yes | Yes | Yes | Yes | Yes | Low |
| 99 | Martinón-Torres, 2023[200], NCT04031846[201] | Yes | Yes | Yes | Yes | Yes | Yes | Yes | Low |
| 100 | Benfield, 2023[202], NCT04016714[203] | Yes | Yes | Yes | Yes | Yes | Yes | Yes | Low |
| 101 | Suzuki, 2023[204], NCT04384107[205] | Yes | Yes | Yes | Yes | Yes | Yes | Yes | Low |
| 102 | Ishihara, 2023[206], Wan, 2024[207], NCT03848065[208] | Yes | Yes | Yes | Yes | Yes | Yes | Yes | Low |
| 103 | Lupinacci, 2023[209], NCT03893448[210] | Yes | Yes | Yes | Yes | Yes | Yes | Yes | Low |
| 104 | Senders, 2021[211], NCT03512288[212] | Yes | Yes | Yes | Yes | Yes | Yes | Yes | Low |
| 105 | Korbal, 2024[213], NCT04546425[214] | Yes | Yes | Yes | Yes | Yes | Yes | Yes | Low |
| 106 | Ishihara, 2024[215], NCT04530838[216] | Yes | Yes | Yes | Yes | Yes | Yes | Yes | Low |
| 107 | Senders, 2024[217], NCT04382326[218], EUCTR2019-003305-10[219] | Yes | Yes | Yes | Yes | Yes | Yes | Yes | Low |
| 108 | Clarke, 2020[220], NCT02308540[221] | Yes | Yes | Yes | Yes | Yes | Yes | Yes | Low |
| 109 | Clarke, 2021[222], NCT03197376[223] | Yes | Yes | Yes | Yes | Yes | Yes | Yes | Low |
| 110 | Adigweme, 2023[224], NCT03896477[225] | Yes | Yes | Yes | Yes | Yes | Yes | Yes | Low |

### Table S4. Risk of bias assessment according to the modified JBI tool for quasi experimental study

| ID | Author & Year | Q3 | Q4 | Q6 | Q7 | Q8 | Q9 | Overall |
|---|---|---|---|---|---|---|---|---|
| 111 | Nurkka, 2001[226] | Unclear | Yes | Yes | Yes | Yes | Yes | Moderate |
| 112 | Käyhty, 2005[227] | Single-arm | Single-arm | Single-arm | Yes | Yes | Yes | Low |
| 113 | Shao, 2006[228] | Single-arm | Single-arm | Single-arm | Yes | Yes | Yes | Low |
| 114 | Kim, 2007[229] | Single-arm | Single-arm | Single-arm | Yes | Yes | Yes | Low |
| 115 | Lee, 2009[230] | Single-arm | Single-arm | Single-arm | Yes | Yes | Yes | Low |
| 116 | Li, 2015[231] | Subjects were stratified into 4 groups based on age | Yes | Yes | Yes | Yes | Yes | Low |
| 117 | Togashi, 2013[232], NCT00574795[233] | Single-arm | Single-arm | Single-arm | Yes | Yes | Yes | Low |
| 118 | Rodgers, 2013[96], Gutiérrez Brito, 2013[234], NCT00708682[235] | Single-arm | Single-arm | Single-arm | Yes | Yes | Yes | Low |
| 119 | Singleton, 2013[236] | Subjects were stratified into 5 groups based on age | Yes | Yes | Yes | No | Yes | High |
| 120 | Wijmenga-Monsuur, 2015[237], van Westen, 2015[238], van Westen, 2018[129] | Subjects were stratified into 2 groups based on birth date | Yes | Yes | Yes | Yes | Yes | Low |
| 121 | Martinón-Torres, 2015[239], Martinón-Torres, 2017[240], NCT01193335[241] | Subjects were designated preterm or term based on GA at birth. | Yes | Yes | Yes | Yes | Yes | Low |
| 122 | Chu, 2023[242], NCT03574389[243] | Subjects were stratified into 4 groups based on age | Yes | Yes | Unclear | No | Yes | High |
| 123 | Urbancikova, 2017[244] | Yes | No | Yes | Yes | Yes | Yes | Low |
| 124 | Maestri, 2024[245], NCT04633226[246] | Single-arm | Single-arm | Single-arm | Yes | Yes | Yes | Low |

### Table S5. Risk of bias assessment according to the modified JBI tool for cohort study

| ID | Author & Year | Q1 | Q2 | Q7 | Q8 | Q9 | Q11 | Overall |
|---|---|---|---|---|---|---|---|---|
| 125 | O'Brien, 2000[247] | Yes | Yes | Yes | Yes | Yes | Yes | Low |
| 126 | Esposito, 2005[248] | Yes | Yes | Yes | Yes | Yes | Yes | Low |
| 127 | Osendarp, 2007[249] | Yes | Yes | Yes | Yes | Yes | Yes | Low |
| 128 | Vesikari, 2010[250] | Yes | Yes | Yes | Yes | Yes | Yes | Low |
| 129 | Moss, 2010[251], Moss, 2010[252] | Yes | Yes | Yes | Yes | No | Yes | Moderate |
| 130 | Whelan, 2012[253] | Yes | Yes | Yes | Yes | Yes | Yes | Low |
| 131 | Jones, 2013[254] | Single-arm | Single-arm | Yes | Yes | Yes | Yes | Low |
| 132 | Madhi, 2013[255], Madhi et all. 2010[256], Madhi, 2020[257] | No, but only HIV-uninfected infants born to HIV non-infected mothers were included | Yes | Yes | Yes | Yes | Yes | Low |
| 133 | Ladhani, 2015[258] | Single-arm | Single-arm | Yes | Yes | Yes | Partly yes, no details of method for calculating 95%CI | Moderate |
| 134 | Ladhani, 2015[259] | Single-arm | Single-arm | Yes | Yes | Yes | Partly yes, no details of method for calculating 95%CI | Moderate |
| 135 | Maertens, 2017[260] | Yes | Yes | Yes | Yes | Yes | Partly yes, no details of method for calculating 95%CI | Moderate |
| 136 | Madhi, 2017[261] | Yes | Yes | Yes | Yes | Yes | Yes | Low |
| 137 | Zimmermann, 2020[262], Zimmermann, 2019[263], Zimmermann, 2019[264] | Yes | Yes | Yes | Yes | No | Yes | Moderate |
| 138 | Perrett, 2020[265], Martinón-Torres, 2021[266], NCT02422264[267], NCT02853929[268] | Yes | Yes | Yes | Yes | Yes | Yes | Low |

#### Table S6. Distribution of included study arms by vaccine products and countries conducted

| Country* | Number of study arm by vaccine products | | | | |
|---|---|---|---|---|---|
| | PCV7 | PCV13 | PCV15 | PCV20 | PCV10-SII |
| Australia | - | 3 | - | - | - |
| Bangladesh | 1 | - | - | - | - |
| Belgium | - | 2 | - | - | - |
| Brazil | 1 | 1 | - | - | - |
| Burkina Faso | - | 2 | - | - | - |
| Canada | 5 | 1 | - | - | - |
| China | 8 | 8 | - | - | - |
| Czechia | - | 1 | - | - | - |
| Finland | 2 | - | - | - | - |
| France | 1 | 1 | - | - | - |
| Gambia (the) | - | 6 | - | - | 3 |
| Germany | 5 | 1 | - | - | - |
| India | 2 | 8 | - | - | - |
| Israel | 4 | 1 | - | - | - |
| Italy | 2 | 1 | - | - | - |
| Japan | 2 | 5 | 3 | 2 | - |
| Kenya | 2 | 1 | - | - | - |
| Korea (the Republic of) | 4 | 1 | 1 | - | - |
| Mexico | - | 1 | - | - | - |
| Multi-countries | 18 | 31 | 8 | 2 | - |
| Netherlands (the) | 4 | 5 | - | - | - |
| Philippines (the) | 1 | - | - | - | - |
| Poland | 3 | 4 | - | - | - |
| Russian Federation (the) | - | 2 | - | - | - |
| Slovakia | - | 1 | - | - | - |
| South Africa | 2 | 4 | - | - | - |
| Spain | 1 | 2 | - | - | - |
| Sweden | 1 | - | - | - | - |
| Thailand | 1 | 3 | - | - | - |
| United Kingdom of Great Britain and Northern Ireland (the) | 6 | 8 | - | - | - |
| United States of America (the) | 25 | 15 | 1 | 1 | - |
| Viet Nam | 1 | 2 | - | - | - |

*"Multi-countries" indicates study arm conducted across multiple sites involving more than one country.

#### Table S7. Detailed breakdown of countries involved in "Multi-countries" studies and corresponding number of study arms

| Country | Number of study arm by vaccine products | | | |
|---|---|---|---|---|
| | PCV7 | PCV13 | PCV15 | PCV20 |
| Argentina | 5 | - | - | - |
| Australia | - | 8 | 1 | 1 |
| Austria | 2 | - | - | - |
| Belgium | - | 2 | 1 | 1 |
| Canada | - | 13 | 3 | - |
| Chile | 3 | - | - | - |
| Colombia | 5 | - | - | - |
| Costa Rica | 3 | - | - | - |
| Czechia | 5 | 12 | 1 | 1 |
| Denmark | - | 3 | 3 | 1 |
| Dominican Republic (the) | - | 3 | - | - |
| Estonia | - | 2 | 1 | 1 |
| Finland | 4 | 11 | 4 | 1 |
| France | 6 | 2 | - | - |
| Germany | 6 | 7 | 1 | - |
| Greece | - | 1 | 1 | - |
| Hungary | 3 | 1 | - | - |
| Israel | - | 1 | 2 | - |
| Italy | 5 | 6 | 1 | 1 |
| Malaysia | - | 1 | 1 | - |
| Malta | - | 2 | - | - |
| Netherlands (the) | - | 1 | - | 1 |
| Norway | - | 2 | 1 | 1 |
| Panama | - | 2 | - | - |
| Poland | 4 | 7 | 2 | 1 |
| Puerto Rico | - | 3 | 2 | 1 |
| Russian Federation (the) | - | 3 | 2 | 1 |
| Slovakia | - | 1 | - | 1 |
| South Africa | - | 2 | - | - |
| Spain | 1 | 10 | 4 | - |

| Country | Number of study arm by vaccine products | | | |
|---|---|---|---|---|
| | PCV7 | PCV13 | PCV15 | PCV20 |
| Sweden | - | 1 | - | - |
| Thailand | - | 3 | 3 | - |
| Turkey | - | 4 | 2 | - |
| United Kingdom of Great Britain and Northern Ireland (the) | - | 2 | - | - |
| United States of America (the) | 3 | 7 | 3 | 1 |

**Table S8. Number of study arms reporting IgG geometric mean concentrations (IgG GMCs) and seroresponse rates post-childhood-schedule by vaccine product**

| Vaccine product | Num of study arms with reported IgG GMCs | Num of study arms with reported seroresponse rates |
|---|---|---|
| PCV7 | 47 | 23 |
| PCV13 | 77 | 58 |
| PCV15 | 9 | 3 |
| PCV20 | 4 | 3 |
| PCV10-SII | 3 | 1 |

**Table S9. Number of study arms reporting IgG geometric mean concentrations (IgG GMCs) and seroresponse rates post-childhood-schedule by vaccine product and schedule**

| Vaccine product | Vaccine schedule | Num of study arms with reported IgG GMCs | Num of study arms with reported seroresponse rates |
|---|---|---|---|
| PCV7 | 3+1 | 34 | 16 |
| | 2+1 | 7 | 4 |
| | 1+1 | - | - |
| | 3+0 | 6 | 3 |
| PCV13 | 3+1 | 46 | 26 |
| | 2+1 | 19 | 19 |
| | 1+1 | 7 | 7 |
| | 3+0 | 5 | 6 |
| PCV15 | 3+1 | 7 | 1 |
| | 2+1 | 2 | 2 |
| | 1+1 | - | - |
| | 3+0 | - | - |
| PCV20 | 3+1 | 3 | 2 |
| | 2+1 | 1 | 1 |
| | 1+1 | - | - |
| | 3+0 | - | - |
| PCV10-SII | 3+1 | 2 | - |
| | 2+1 | 1 | 1 |
| | 1+1 | - | - |
| | 3+0 | - | - |

**Table S10. Number of study arms reporting IgG geometric mean concentrations (IgG GMCs) and seroresponse rates post "3+1" vaccination schedule by vaccine product and WHO region**

| Vaccine product | WHO region | Num of study arms with reported IgG GMCs | Num of study arms with reported seroresponse rates |
|---|---|---|---|
| PCV7 | AFR | - | - |
| | AMR | 10 | 5 |
| | EUR | 13 | 6 |
| | SEAR | 3 | 2 |
| | WPR | 7 | 2 |
| | Multi | 1 | 1 |
| PCV13 | AFR | 3 | 1 |
| | AMR | 16 | 7 |
| | EUR | 10 | 7 |
| | SEAR | 4 | 4 |
| | WPR | 5 | 3 |
| | Multi | 8 | 4 |
| PCV15 | AFR | - | - |
| | AMR | - | - |
| | EUR | - | - |
| | SEAR | - | - |
| | WPR | 2 | 1 |
| | Multi | 5 | - |
| PCV20 | AFR | - | - |
| | AMR | 2 | 1 |
| | EUR | - | - |
| | SEAR | - | - |
| | WPR | 1 | 1 |
| | Multi | - | - |

| Vaccine product | WHO region | Num of study arms with reported IgG GMCs | Num of study arms with reported seroresponse rates |
|---|---|---|---|
| PCV10-SII | AFR | 2 | - |
| | AMR | - | - |
| | EUR | - | - |
| | SEAR | - | - |
| | WPR | - | - |
| | Multi | - | - |

***Abbreviations:*** AFR: African Region; AMR: Region of the Americas; EUR: European Region; SEAR: South-East Asia Region; WPR: Western Pacific Region; Multi: Multi regions.

**Table S11. Number of study arms reporting IgG geometric mean concentrations (IgG GMCs) and seroresponse rates post different timepoints by vaccine product**

| Vaccine product | Timepoint | Num of study arms with reported IgG GMCs | Num of study arms with reported seroresponse rates |
|---|---|---|---|
| PCV7 | Post 1-dose primary | 3 | 3 |
| | Post 2-dose primary | 11 | 11 |
| | Post 3-dose primary | 36 | 28 |
| | Post booster | 41 | 20 |
| PCV13 | Post 1-dose primary | 5 | 5 |
| | Post 2-dose primary | 21 | 23 |
| | Post 3-dose primary | 49 | 52 |
| | Post booster | 72 | 52 |
| PCV15 | Post 1-dose primary | - | - |
| | Post 2-dose primary | 2 | 2 |
| | Post 3-dose primary | 7 | 7 |
| | Post booster | 9 | 3 |
| PCV20 | Post 1-dose primary | - | - |
| | Post 2-dose primary | 1 | 1 |
| | Post 3-dose primary | 3 | 3 |
| | Post booster | 4 | 3 |
| PCV10-SII | Post 1-dose primary | - | - |
| | Post 2-dose primary | 1 | 1 |
| | Post 3-dose primary | 2 | 2 |
| | Post booster | 3 | 1 |

**Table S12. Summary of the meta-analysis results of anti-pneumococcal IgG geometric mean concentrations (GMCs, µg/mL) post-childhood-schedule by vaccine product and serotype**

| Serotype (ST) | IgG GMCs (95%CI) by vaccine product (µg/mL) | | | | |
|---|---|---|---|---|---|
| | PCV7 | PCV13 | PCV15 | PCV20 | PCV10-SII |
| PCV7 covered STs | | | | | |
| 4 | 4·02 (3·37, 4·79) | 3·73 (3·25, 4·30) | 1·56 (1·31, 1·86) | 5·32 (3·76, 7·52) | - |
| 6B | 6·92 (5·42, 8·84) | 7·13 (6·30, 8·07) | 5·63 (4·87, 6·52) | 4·73 (2·98, 7·52) | 10·76 (8·16, 14·20) |
| 9V | 3·45 (3·02, 3·93) | 3·23 (2·90, 3·60) | 2·73 (2·31, 3·24) | 4·38 (3·44, 5·59) | 2·47 (1·70, 3·60) |
| 14 | 10·55 (9·36, 11·90) | 9·47 (8·73, 10·28) | 6·75 (5·72, 7·97) | 6·69 (4·80, 9·33) | 7·49 (6·58, 8·52) |
| 18C | 3·10 (2·76, 3·49) | 3·00 (2·69, 3·36) | 2·65 (2·27, 3·10) | 3·74 (2·79, 5·02) | - |
| 19F | 4·54 (3·91, 5·27) | 8·07 (7·30, 8·92) | 4·72 (4·14, 5·37) | 6·71 (5·30, 8·51) | 8·09 (5·73, 11·43) |
| 23F | 4·90 (4·16, 5·76) | 4·17 (3·67, 4·73) | 2·23 (1·86, 2·67) | 4·64 (2·96, 7·27) | 4·54 (3·96, 5·21) |
| PCV13-non-PCV7 STs | | | | | |
| 1 | - | 4·26 (3·75, 4·85) | 1·73 (1·43, 2·09) | 2·07 (1·51, 2·82) | 6·76 (5·28, 8·65) |
| 3 | - | 0·98 (0·89, 1·08) | 1·08 (0·93, 1·25) | 0·84 (0·60, 1·17) | - |
| 5 | - | 3·00 (2·69, 3·34) | 2·85 (2·23, 3·64) | 2·38 (1·72, 3·29) | 1·64 (1·19, 2·26) |
| 6A | - | 7·98 (7·15, 8·90) | 4·18 (3·61, 4·82) | 10·73 (8·02, 14·36) | 7·49 (4·79, 11·72) |
| 7F | - | 5·21 (4·81, 5·64) | 3·76 (3·31, 4·27) | 4·52 (3·58, 5·71) | 6·48 (6·09, 6·90) |
| 19A | - | 8·17 (7·56, 8·83) | 4·84 (4·11, 5·71) | 5·17 (3·68, 7·25) | 6·13 (3·82, 9·82) |
| PCV15-non-PCV13 STs | | | | | |
| 22F | - | - | 8·13 (6·78, 9·75) | 11·89 (9·55, 14·82) | - |
| 33F | - | - | 4·67 (3·89, 5·62) | 7·47 (5·12, 10·90) | - |
| PCV20-non-PCV15 STs | | | | | |

| Serotype (ST) | IgG GMCs (95%CI) by vaccine product (μg/mL) | | | | |
|---|---|---|---|---|---|
| | PCV7 | PCV13 | PCV15 | PCV20 | PCV10-SII |
| 8 | - | - | - | 4·01 (3·08, 5·23) | - |
| 10A | - | - | - | 6·98 (5·15, 9·45) | - |
| 11A | - | - | - | 4·55 (3·51, 5·90) | - |
| 12F | - | - | - | 2·04 (1·72, 2·44) | - |
| 15B | - | - | - | 16·00 (12·31, 20·80) | - |

Heterogeneity for PCV7: $I^2$=98·7%, $\tau^2$=0·462, p <0·01; for PCV13: $I^2$=99·3%, $\tau^2$=0·572, p <0·01; for PCV15: $I^2$=99·4%, $\tau^2$=0·361, p <0·01; for PCV20: $I^2$=99·6%, $\tau^2$=0·508, p <0·01; for PCV10-SII: $I^2$=99·1%, $\tau^2$=0·354, p <0·01.

**Table S13. Summary of the meta-analysis results of anti-pneumococcal IgG seroresponse rates (%) post-childhood-schedule by vaccine product**

| Serotype (ST) | Seroresponse rate (95%CI) by vaccine product (%) | | | | |
|---|---|---|---|---|---|
| | PCV7 | PCV13 | PCV15 | PCV20 | PCV10-SII† |
| PCV7 covered STs | | | | | |
| 4 | 98% (97%-99%) | 99% (98%-99%) | 96% (95%-97%) | 98% (91%-100%) | - |
| 6B | 98% (96%-99%) | 98% (97%-99%) | 98% (96%-99%) | 99% (98%-99%) | 100% (98%-100%) |
| 9V | 99% (98%-99%) | 98% (98%-99%) | 99% (98%-100%) | 99% (98%-99%) | 99% (96%-100%) |
| 14 | 99% (98%-99%) | 99% (98%-99%) | 99% (99%-100%) | 98% (94%-100%) | 98% (96%-100%) |
| 18C | 99% (98%-99%) | 98% (98%-99%) | 99% (98%-100%) | 99% (98%-99%) | - |
| 19F | 98% (97%-99%) | 98% (98%-99%) | 100% (99%-100%) | 99% (98%-100%) | 100% (98%-100%) |
| 23F | 98% (97%-99%) | 98% (97%-98%) | 97% (96%-98%) | 98% (94%-99%) | 98% (95%-99%) |
| PCV13-non-PCV7 STs | | | | | |
| 1 | - | 99% (98%-99%) | 97% (95%-98%) | 97% (93%-98%) | 100% (98%-100%) |
| 3 | - | 91% (88%-94%) | 92% (91%-94%) | 84% (70%-92%) | - |
| 5 | - | 98% (98%-99%) | 99% (98%-100%) | 98% (97%-99%) | 98% (94%-99%) |
| 6A | - | 99% (98%-99%) | 99% (98%-99%) | 99% (98%-100%) | 98% (95%-99%) |
| 7F | - | 99% (99%-99%) | 100% (99%-100%) | 100% (99%-100%) | 100% (98%-100%) |
| 19A | - | 99% (99%-100%) | 99% (99%-100%) | 100% (99%-100%) | 100% (98%-100%) |
| PCV15-non-PCV13 STs | | | | | |
| 22F | - | - | 100% (99%-100%) | 99% (99%-100%) | - |
| 33F | - | - | 99% (98%-100%) | 99% (98%-100%) | - |
| PCV20-non-PCV15 STs | | | | | |
| 8 | - | - | - | 99% (99%-100%) | - |
| 10A | - | - | - | 98% (96%-99%) | - |
| 11A | - | - | - | 99% (98%-99%) | - |
| 12F | - | - | - | 97% (94%-98%) | - |
| 15B | - | - | - | 100% (99%-100%) | - |

†: Only 1 study arm available for PCV10-SII, no pooled estimate or heterogeneity.

Heterogeneity for PCV7: $I^2$=79·2%, $\tau^2$=0·972, p <0·01; for PCV13: $I^2$=84·7%, $\tau^2$=0·979, p <0·01; for PCV15: $I^2$=81·1%, $\tau^2$=0·595, p <0·01; for PCV20: $I^2$=95·2%, $\tau^2$=1·063, p <0·01.

**Table S14. Summary of the meta-analysis results of anti-pneumococcal IgG geometric mean concentrations (IgG GMCs, µg/mL) post-childhood-schedule by vaccine product and schedule**

| Serotype | IgG GMCs (95%CI) by vaccine schedule (µg/mL) | | | |
|---|---|---|---|---|
| | 3+1 | 2+1 | 1+1 | 3+0 |
| PCV7 | | | | |
| 4 | 4·19 (3·41, 5·15) | 5·68 (4·23, 7·63) | - | 2·08 (1·61, 2·68) |
| 6B | 9·43 (8·15, 10·91) | 6·34 (4·38, 9·17) | - | 1·22 (0·53, 2·80) |
| 9V | 3·76 (3·35, 4·22) | 4·19 (3·10, 5·66) | - | 1·58 (1·17, 2·14) |
| 14 | 11·25 (9·95, 12·72) | 13·04 (11·09, 15·33) | - | 5·50 (4·17, 7·25) |
| 18C | 3·32 (2·90, 3·81) | 2·87 (2·25, 3·68) | - | 2·22 (1·58, 3·11) |
| 19F | 5·01 (4·37, 5·74) | 5·81 (4·64, 7·28) | - | 1·89 (1·26, 2·85) |
| 23F | 5·93 (5·14, 6·83) | 4·50 (3·64, 5·57) | - | 1·67 (1·25, 2·24) |
| PCV13 | | | | |
| 4 | 3·51 (2·90, 4·26) | 3·47 (2·68, 4·49) | 6·05 (4·03, 9·10) | 4·33 (3·29, 5·70) |
| 6B | 9·07 (8·17, 10·07) | 6·75 (5·54, 8·23) | 3·12 (2·42, 4·00) | 2·64 (1·73, 4·04) |
| 9V | 3·10 (2·67, 3·60) | 3·45 (2·87, 4·14) | 4·27 (2·92, 6·24) | 2·60 (2·16, 3·14) |
| 14 | 9·74 (8·93, 10·64) | 9·40 (8·08, 10·93) | 12·69 (9·17, 17·56) | 5·06 (3·77, 6·79) |
| 18C | 3·19 (2·72, 3·74) | 2·86 (2·38, 3·44) | 2·44 (1·73, 3·44) | 2·89 (2·46, 3·39) |
| 19F | 7·67 (6·92, 8·50) | 8·22 (6·85, 9·86) | 15·99 (10·73, 23·82) | 4·90 (4·60, 5·23) |
| 23F | 4·89 (4·20, 5·68) | 3·76 (2·89, 4·90) | 2·76 (1·96, 3·89) | 2·44 (1·95, 3·06) |
| 1 | 3·82 (3·32, 4·40) | 4·08 (3·18, 5·23) | 11·51 (9·23, 14·36) | 3·52 (2·58, 4·80) |
| 3 | 0·96 (0·86, 1·07) | 0·89 (0·74, 1·06) | 1·10 (0·69, 1·76) | 1·39 (1·08, 1·79) |
| 5 | 3·24 (2·84, 3·70) | 2·76 (2·25, 3·39) | 3·26 (2·11, 5·03) | 1·76 (1·30, 2·37) |
| 6A | 8·74 (7·89, 9·68) | 8·39 (7·03, 10·01) | 8·67 (6·22, 12·06) | 2·39 (1·55, 3·69) |
| 7F | 5·76 (5·21, 6·36) | 4·76 (4·06, 5·56) | 4·46 (3·79, 5·26) | 3·64 (3·06, 4·34) |
| 19A | 8·64 (7·85, 9·50) | 7·44 (6·42, 8·62) | 10·20 (8·44, 12·32) | 5·17 (3·97, 6·72) |
| PCV15 | | | | |
| 4 | 1·64 (1·32, 2·04) | 1·35 (1·24, 1·47) | - | - |
| 6B | 6·07 (5·28, 6·99) | 4·36 (4·00, 4·76) | - | - |
| 9V | 2·95 (2·45, 3·55) | 2·14 (2·05, 2·23) | - | - |
| 14 | 7·26 (6·06, 8·70) | 5·30 (5·02, 5·60) | - | - |
| 18C | 2·87 (2·47, 3·34) | 2·02 (1·87, 2·18) | - | - |
| 19F | 4·94 (4·23, 5·77) | 4·08 (3·90, 4·28) | - | - |
| 23F | 2·48 (2·12, 2·89) | 1·55 (1·47, 1·63) | - | - |
| 1 | 1·88 (1·54, 2·30) | 1·29 (1·23, 1·34) | - | - |
| 3 | 1·16 (1·00, 1·34) | 0·84 (0·80, 0·89) | - | - |
| 5 | 3·15 (2·40, 4·14) | 2·02 (1·92, 2·13) | - | - |
| 6A | 4·52 (3·99, 5·11) | 3·16 (2·98, 3·34) | - | - |
| 7F | 4·04 (3·65, 4·47) | 2·93 (2·64, 3·25) | - | - |
| 19A | 4·89 (3·94, 6·08) | 4·71 (4·48, 4·95) | - | - |
| 22F | 9·00 (7·61, 10·64) | 6·02 (5·75, 6·30) | - | - |
| 33F | 5·22 (4·48, 6·07) | 3·35 (3·19, 3·51) | - | - |
| PCV20† | | | | |
| 4 | 5·80 (3·78, 8·90) | 4·11 (3·77, 4·48) | - | - |
| 6B | 5·74 (3·97, 8·31) | 2·64 (2·36, 2·95) | - | - |
| 9V | 4·65 (3·44, 6·30) | 3·68 (3·42, 3·97) | - | - |
| 14 | 7·62 (5·65, 10·27) | 4·52 (4·08, 5·00) | - | - |
| 18C | 4·17 (3·14, 5·55) | 2·71 (2·52, 2·93) | - | - |
| 19F | 6·91 (4·99, 9·58) | 6·19 (5·68, 6·75) | - | - |
| 23F | 5·59 (3·89, 8·06) | 2·64 (2·40, 2·91) | - | - |
| 1 | 2·21 (1·47, 3·30) | 1·71 (1·58, 1·84) | - | - |
| 3 | 0·88 (0·56, 1·39) | 0·72 (0·67, 0·78) | - | - |
| 5 | 2·64 (1·85, 3·77) | 1·74 (1·60, 1·89) | - | - |
| 6A | 11·95 (9·01, 15·85) | 7·75 (7·04, 8·53) | - | - |
| 7F | 4·88 (3·78, 6·30) | 3·61 (3·40, 3·84) | - | - |
| 19A | 5·41 (3·41, 8·59) | 4·51 (4·11, 4·94) | - | - |
| 22F | 12·92 (10·52, 15·88) | 9·27 (8·52, 10·08) | - | - |
| 33F | 7·87 (4·71, 13·17) | 6·37 (5·83, 6·95) | - | - |
| 8 | 4·17 (2·91, 5·98) | 3·57 (3·32, 3·83) | - | - |
| 10A | 7·87 (6·03, 10·26) | 4·86 (4·41, 5·36) | - | - |
| 11A | 4·86 (3·53, 6·69) | 3·74 (3·44, 4·07) | - | - |
| 12F | 2·12 (1·68, 2·67) | 1·86 (1·71, 2·01) | - | - |
| 15B | 17·13 (12·44, 23·60) | 13·09 (12·10, 14·15) | - | - |
| PCV10-SII‡ | | | | |
| 6B | 10·01 (6·63, 15·13) | 12·46 (11·07, 14·01) | - | - |
| 9V | 2·06 (1·53, 2·76) | 3·46 (3·08, 3·88) | - | - |
| 14 | 6·98 (6·25, 7·81) | 8·28 (6·97, 9·82) | - | - |
| 19F | 6·58 (5·40, 8·00) | 11·11 (9·70, 12·73) | - | - |
| 23F | 4·35 (3·61, 5·25) | 4·95 (4·28, 5·73) | - | - |
| 1 | 5·74 (5·29, 6·22) | 8·45 (7·54, 9·48) | - | - |
| 5 | 1·72 (0·99, 2·98) | 1·54 (1·38, 1·73) | - | - |
| 6A | 6·63 (3·45, 12·77) | 9·56 (8·26, 11·05) | - | - |
| 7F | 6·40 (5·93, 6·90) | 6·66 (5·96, 7·44) | - | - |
| 19A | 5·00 (3·00, 8·34) | 8·82 (7·65, 10·15) | - | - |

†: Only 1 study arm available for PCV20 (2+1 schedule), no pooled estimate or heterogeneity.
‡: Only 1 study arm available for PCV10-SII (2+1 schedule), no pooled estimate or heterogeneity.
Heterogeneity for PCV7 (3+1): $I^2$=98·7%, $\tau^2$=0·361, p <0·01; for PCV7 (2+1): $I^2$=97·6%, $\tau^2$=0·295, p <0·01; for PCV7 (3+0): $I^2$96·7%, $\tau^2$=0·417, p <0·01; for PCV13 (3+1): $I^2$=99·4%, $\tau^2$=0·576, p <0·01; for PCV13 (2+1): $I^2$=99·3%, $\tau^2$=0·538, p <0·01; for PCV13 (1+1): $I^2$=98·6%, $\tau^2$=0·736, p <0·01; for PCV13 (3+0): $I^2$=97·8%, $\tau^2$=0·229, p <0·01; for PCV15 (3+1): $I^2$=99·2%, $\tau^2$=0·350, p <0·01; for PCV15 (2+1): $I^2$=99·6%, $\tau^2$=0·329, p <0·01; for PCV20 (3+1): $I^2$=99·6%, $\tau^2$=0·509, p <0·01; for PCV10-SII (3+1): $I^2$=99·1%, $\tau^2$=0·329, p <0·01.

**Table S15. Summary of the meta-analysis results of anti-pneumococcal IgG seroresponse rates (%) post-childhood-schedule by vaccine product and schedule**

| Serotype | Seroresponse rate (95%CI) by vaccine schedule (%) | | | |
|---|---|---|---|---|
| | 3+1 | 2+1 | 1+1 | 3+0 |
| PCV7 | | | | |
| 4 | 98% (97%-99%) | 100% (98%-100%) | - | 98% (94%-99%) |
| 6B | 99% (98%-99%) | 97% (86%-99%) | - | 79% (17%-98%) |
| 9V | 99% (98%-100%) | 98% (95%-99%) | - | 94% (88%-98%) |
| 14 | 99% (98%-100%) | 99% (97%-100%) | - | 96% (92%-98%) |
| 18C | 99% (98%-99%) | 100% (98%-100%) | - | 98% (95%-99%) |
| 19F | 99% (98%-99%) | 98% (96%-99%) | - | 95% (91%-98%) |
| 23F | 99% (98%-99%) | 98% (97%-99%) | - | 92% (70%-98%) |
| PCV13 | | | | |
| 4 | 99% (98%-99%) | 98% (97%-99%) | 100% (99%-100%) | 98% (96%-99%) |
| 6B | 99% (99%-100%) | 98% (97%-99%) | 96% (93%-98%) | 90% (82%-94%) |
| 9V | 99% (98%-99%) | 98% (97%-99%) | 98% (96%-99%) | 97% (95%-98%) |
| 14 | 99% (99%-99%) | 99% (98%-99%) | 98% (94%-99%) | 98% (97%-99%) |
| 18C | 99% (98%-99%) | 98% (97%-99%) | 98% (96%-99%) | 97% (94%-98%) |
| 19F | 99% (98%-99%) | 99% (98%-99%) | 98% (96%-99%) | 98% (97%-98%) |
| 23F | 99% (98%-99%) | 97% (96%-98%) | 96% (94%-98%) | 93% (88%-96%) |
| 1 | 99% (98%-99%) | 99% (98%-99%) | 99% (98%-100%) | 97% (93%-99%) |
| 3 | 92% (87%-95%) | 89% (83%-93%) | 90% (79%-95%) | 97% (88%-99%) |
| 5 | 99% (98%-99%) | 98% (97%-99%) | 99% (97%-100%) | 94% (91%-96%) |
| 6A | 99% (99%-100%) | 99% (98%-99%) | 97% (95%-99%) | 96% (91%-98%) |
| 7F | 99% (99%-100%) | 99% (99%-100%) | 99% (98%-100%) | 99% (96%-100%) |
| 19A | 100% (99%-100%) | 99% (98%-100%) | 100% (99%-100%) | 99% (99%-100%) |
| PCV15* | | | | |
| 4 | 100% (92%-100%) | 96% (95%-97%) | - | - |
| 6B | 100% (92%-100%) | 98% (95%-100%) | - | - |
| 9V | 100% (92%-100%) | 99% (97%-100%) | - | - |
| 14 | 100% (92%-100%) | 100% (98%-100%) | - | - |
| 18C | 100% (92%-100%) | 99% (97%-100%) | - | - |
| 19F | 100% (92%-100%) | 100% (99%-100%) | - | - |
| 23F | 100% (92%-100%) | 97% (96%-98%) | - | - |
| 1 | 100% (92%-100%) | 97% (95%-98%) | - | - |
| 3 | 98% (87%-100%) | 92% (91%-94%) | - | - |
| 5 | 100% (92%-100%) | 99% (98%-100%) | - | - |
| 6A | 100% (92%-100%) | 99% (98%-99%) | - | - |
| 7F | 100% (92%-100%) | 100% (99%-100%) | - | - |
| 19A | 100% (92%-100%) | 99% (98%-100%) | - | - |
| 22F | 100% (92%-100%) | 100% (99%-100%) | - | - |
| 33F | 100% (92%-100%) | 99% (98%-100%) | - | - |
| PCV20† | | | | |
| 4 | 97% (79%-100%) | 99% (98%-100%) | - | - |
| 6B | 99% (98%-100%) | 98% (97%-99%) | - | - |
| 9V | 99% (93%-100%) | 99% (98%-100%) | - | - |
| 14 | 99% (98%-99%) | 97% (95%-98%) | - | - |
| 18C | 99% (98%-99%) | 99% (98%-100%) | - | - |
| 19F | 99% (97%-100%) | 100% (99%-100%) | - | - |
| 23F | 98% (92%-100%) | 96% (94%-98%) | - | - |
| 1 | 97% (88%-99%) | 97% (95%-98%) | - | - |
| 3 | 84% (59%-95%) | 83% (79%-86%) | - | - |
| 5 | 99% (95%-100%) | 98% (97%-99%) | - | - |
| 6A | 100% (99%-100%) | 99% (97%-100%) | - | - |
| 7F | 100% (99%-100%) | 100% (99%-100%) | - | - |
| 19A | 100% (99%-100%) | 100% (99%-100%) | - | - |
| 22F | 100% (99%-100%) | 99% (98%-100%) | - | - |
| 33F | 100% (99%-100%) | 99% (97%-99%) | - | - |
| 8 | 100% (99%-100%) | 99% (98%-100%) | - | - |
| 10A | 99% (94%-100%) | 98% (96%-99%) | - | - |
| 11A | 99% (97%-100%) | 98% (97%-99%) | - | - |
| 12F | 97% (91%-99%) | 97% (95%-98%) | - | - |
| 15B | 100% (99%-100%) | 99% (98%-100%) | - | - |
| PCV10-SII‡ | | | | |

| Serotype | Seroresponse rate (95%CI) by vaccine schedule (%) | | | |
|---|---|---|---|---|
| | 3+1 | 2+1 | 1+1 | 3+0 |
| 6B | - | 100% (98%-100%) | - | - |
| 9V | - | 99% (96%-100%) | - | - |
| 14 | - | 98% (96%-100%) | - | - |
| 19F | - | 100% (98%-100%) | - | - |
| 23F | - | 98% (95%-99%) | - | - |
| 1 | - | 100% (98%-100%) | - | - |
| 5 | - | 98% (94%-99%) | - | - |
| 6A | - | 98% (95%-99%) | - | - |
| 7F | - | 100% (98%-100%) | - | - |
| 19A | - | 100% (98%-100%) | - | - |

*: Only 1 study arm available for PCV15 (3+1 schedule), no pooled estimate or heterogeneity.
†: Only 1 study arm available for PCV20 (2+1 schedule), no pooled estimate or heterogeneity.
‡: Only 1 study arm available for PCV10-SII (2+1 schedule), no pooled estimate or heterogeneity.
Heterogeneity for PCV7 (3+1): $I^2$=61·5%, $\tau^2$=0·732, p <0·01; for PCV7 (2+1): $I^2$=35·9%, $\tau^2$=0·192, p <0·01; for PCV7 (3+0): $I^2$=87·1%, $\tau^2$=1·284, p <0·01; for PCV13 (3+1): $I^2$=85·0%, $\tau^2$=0·916, p <0·01; for PCV13 (2+1): $I^2$=83·2%, $\tau^2$=0·858, p <0·01; for PCV13 (1+1): $I^2$=63·3%, $\tau^2$=0·755, p <0·01; for PCV13 (3+0): $I^2$=90·3%, $\tau^2$=0·952, p <0·01; for PCV15 (2+1): $I^2$=87·0%, $\tau^2$=0·939, p <0·01; for PCV20 (3+1): $I^2$=95·6%, $\tau^2$=1·286, p <0·01.

**Table S16. Summary of the meta-analysis results of anti-pneumococcal IgG geometric mean concentrations (IgG GMCs, µg/mL) post "3+1" vaccination schedule by vaccine product and region**

| Serotype | IgG GMCs (95%CI) by WHO region (µg/mL) | | | | | |
|---|---|---|---|---|---|---|
| | Africa | Americas | Europe | South East Asia | Western Pacific | Multi-regions |
| PCV7† | | | | | | |
| 4 | - | 2·29 (1·69, 3·11) | 4·25 (3·78, 4·78) | 5·76 (5·35, 6·20) | 8·91 (6·03, 13·16) | 2·80 (1·90, 4·00) |
| 6B | - | 9·27 (7·62, 11·26) | 7·27 (5·89, 8·97) | 10·73 (9·16, 12·57) | 15·52 (12·16, 19·82) | 7·00 (4·50, 10·90) |
| 9V | - | 3·19 (2·92, 3·49) | 3·41 (2·81, 4·13) | 3·89 (2·86, 5·29) | 5·38 (4·25, 6·82) | 5·90 (3·60, 9·90) |
| 14 | - | 7·87 (6·67, 9·29) | 11·60 (10·12, 13·28) | 12·24 (9·87, 15·16) | 17·48 (14·80, 20·64) | 10·80 (6·10, 18·80) |
| 18C | - | 2·89 (2·44, 3·42) | 2·74 (2·35, 3·18) | 3·01 (2·80, 3·23) | 6·02 (4·67, 7·77) | 3·90 (2·70, 5·80) |
| 19F | - | 3·86 (3·49, 4·26) | 4·39 (3·87, 4·98) | 5·72 (5·28, 6·20) | 9·12 (6·66, 12·48) | 4·20 (2·80, 6·10) |
| 23F | - | 4·84 (4·21, 5·56) | 4·80 (4·08, 5·64) | 6·21 (4·54, 8·51) | 11·27 (9·57, 13·27) | 7·10 (4·60, 10·90) |
| PCV13 | | | | | | |
| 4 | 4·54 (3·61, 5·70) | 2·84 (2·04, 3·96) | 3·76 (3·18, 4·45) | 7·71 (5·05, 11·79) | 7·08 (4·21, 11·89) | 2·08 (1·70, 2·55) |
| 6B | 13·97 (10·43, 18·72) | 8·41 (6·83, 10·35) | 8·60 (7·37, 10·04) | 12·04 (11·02, 13·15) | 12·93 (11·19, 14·96) | 6·87 (6·22, 7·60) |
| 9V | 3·89 (3·05, 4·96) | 2·52 (1·89, 3·36) | 2·60 (2·18, 3·11) | 5·23 (2·78, 9·84) | 5·28 (3·97, 7·02) | 3·01 (2·45, 3·70) |
| 14 | 9·44 (6·74, 13·22) | 7·98 (7·16, 8·91) | 10·10 (9·43, 10·81) | 11·19 (10·16, 12·33) | 15·53 (10·60, 22·76) | 9·44 (7·95, 11·21) |
| 18C | 5·75 (4·37, 7·56) | 2·84 (2·12, 3·79) | 2·46 (1·96, 3·10) | 4·04 (2·48, 6·58) | 6·70 (4·53, 9·91) | 2·93 (2·59, 3·32) |
| 19F | 10·42 (7·79, 13·93) | 6·50 (5·65, 7·48) | 7·78 (6·66, 9·08) | 10·74 (8·07, 14·29) | 11·32 (8·05, 15·93) | 6·29 (5·10, 7·77) |
| 23F | 8·85 (5·10, 15·33) | 4·70 (3·74, 5·91) | 3·74 (3·27, 4·29) | 6·62 (4·00, 10·96) | 10·33 (7·56, 14·10) | 3·26 (2·67, 4·00) |
| 1 | 6·06 (4·97, 7·40) | 2·99 (2·46, 3·63) | 4·02 (3·42, 4·72) | 4·70 (4·27, 5·17) | 9·32 (7·51, 11·56) | 2·60 (2·22, 3·04) |
| 3 | 2·45 (1·98, 3·03) | 0·89 (0·75, 1·06) | 0·90 (0·75, 1·09) | 1·21 (0·88, 1·66) | 1·57 (1·39, 1·77) | 0·70 (0·66, 0·75) |
| 5 | 2·64 (2·26, 3·08) | 2·80 (2·21, 3·54) | 3·03 (2·41, 3·80) | 4·37 (3·45, 5·54) | 6·12 (4·11, 9·12) | 2·89 (2·28, 3·66) |
| 6A | 11·42 (8·05, 16·21) | 8·88 (7·35, 10·73) | 6·96 (6·30, 7·68) | 12·48 (7·59, 20·51) | 11·08 (8·60, 14·27) | 7·55 (6·63, 8·60) |
| 7F | 9·32 (7·85, 11·06) | 5·04 (4·23, 6·01) | 5·15 (4·57, 5·80) | 6·23 (4·63, 8·37) | 9·06 (6·83, 12·03) | 5·28 (4·95, 5·63) |
| 19A | 11·70 (8·04, 17·02) | 6·78 (5·97, 7·71) | 9·77 (8·81, 10·85) | 13·46 (12·43, 14·57) | 12·13 (9·92, 14·84) | 7·28 (6·27, 8·46) |
| PCV15 | | | | | | |
| 4 | - | - | - | - | 2·41 (1·46, 3·98) | 1·43 (1·29, 1·59) |
| 6B | - | - | - | - | 7·47 (6·27, 8·90) | 5·72 (4·95, 6·61) |

| Serotype | IgG GMCs (95%CI) by WHO region (µg/mL) | | | | | |
|---|---|---|---|---|---|---|
| | Africa | Americas | Europe | South East Asia | Western Pacific | Multi-regions |
| 9V | - | - | - | - | 4·12 (2·64, 6·43) | 2·62 (2·43, 2·83) |
| 14 | - | - | - | - | 10·22 (7·93, 13·16) | 6·45 (5·85, 7·10) |
| 18C | - | - | - | - | 3·79 (3·21, 4·48) | 2·64 (2·34, 2·99) |
| 19F | - | - | - | - | 6·04 (4·90, 7·43) | 4·61 (3·91, 5·44) |
| 23F | - | - | - | - | 3·08 (2·59, 3·67) | 2·32 (1·97, 2·75) |
| 1 | - | - | - | - | 2·64 (1·93, 3·62) | 1·67 (1·44, 1·94) |
| 3 | - | - | - | - | 1·48 (1·19, 1·84) | 1·07 (0·95, 1·21) |
| 5 | - | - | - | - | 4·61 (2·35, 9·06) | 2·74 (2·24, 3·35) |
| 6A | - | - | - | - | 5·65 (4·73, 6·76) | 4·26 (3·80, 4·77) |
| 7F | - | - | - | - | 4·56 (3·70, 5·61) | 3·93 (3·51, 4·39) |
| 19A | - | - | - | - | 6·22 (3·90, 9·91) | 4·48 (3·59, 5·60) |
| 22F | - | - | - | - | 11·20 (7·67, 16·35) | 8·25 (7·35, 9·27) |
| 33F | - | - | - | - | 6·22 (5·34, 7·25) | 4·88 (4·08, 5·84) |
| PCV20[ǂ] | | | | | | |
| 4 | - | 5·18 (2·76, 9·71) | - | - | 7·31 (6·51, 8·20) | - |
| 6B | - | 5·03 (3·19, 7·91) | - | - | 7·50 (6·58, 8·55) | - |
| 9V | - | 4·34 (2·73, 6·90) | - | - | 5·38 (4·81, 6·02) | - |
| 14 | - | 6·95 (4·62, 10·44) | - | - | 9·19 (8·10, 10·43) | - |
| 18C | - | 4·38 (2·74, 6·99) | - | - | 3·81 (3·38, 4·30) | - |
| 19F | - | 6·22 (4·03, 9·58) | - | - | 8·56 (7·66, 9·56) | - |
| 23F | - | 4·87 (3·20, 7·40) | - | - | 7·39 (6·49, 8·42) | - |
| 1 | - | 1·97 (1·10, 3·51) | - | - | 2·78 (2·47, 3·12) | - |
| 3 | - | 0·80 (0·39, 1·62) | - | - | 1·08 (0·96, 1·21) | - |
| 5 | - | 2·52 (1·40, 4·53) | - | - | 2·94 (2·59, 3·33) | - |
| 6A | - | 11·10 (7·33, 16·82) | - | - | 13·92 (12·43, 15·59) | - |
| 7F | - | 4·90 (3·15, 7·62) | - | - | 4·85 (4·34, 5·42) | - |
| 19A | - | 4·47 (2·79, 7·16) | - | - | 7·92 (7·06, 8·89) | - |
| 22F | - | 12·38 (9·00, 17·03) | - | - | 14·21 (12·61, 16·00) | - |
| 33F | - | 6·63 (3·39, 12·95) | - | - | 11·13 (9·99, 12·39) | - |
| 8 | - | 3·54 (2·79, 4·48) | - | - | 5·88 (5·23, 6·62) | - |
| 10A | - | 7·83 (4·95, 12·38) | - | - | 8·02 (7·02, 9·16) | - |
| 11A | - | 4·47 (2·79, 7·15) | - | - | 5·78 (5·14, 6·50) | - |
| 12F | - | 1·87 (1·75, 1·99) | - | - | 2·69 (2·36, 3·06) | - |
| 15B | - | 15·19 (10·45, 22·10) | - | - | 21·83 (19·53, 24·41) | - |
| PCV10-SII | | | | | | |
| 6B | 10·01 (6·63, 15·13) | - | - | - | - | - |
| 9V | 2·06 (1·53, 2·76) | - | - | - | - | - |
| 14 | 6·98 | - | - | - | - | - |

| Serotype | IgG GMCs (95%CI) by WHO region (µg/mL) | | | | | |
|---|---|---|---|---|---|---|
| | Africa | Americas | Europe | South East Asia | Western Pacific | Multi-regions |
| | (6·25, 7·81) | | | | | |
| 19F | 6·58 (5·40, 8·00) | - | - | - | - | - |
| 23F | 4·35 (3·61, 5·25) | - | - | - | - | - |
| 1 | 5·74 (5·29, 6·22) | - | - | - | - | - |
| 5 | 1·72 (0·99, 2·98) | - | - | - | - | - |
| 6A | 6·63 (3·45, 12·77) | - | - | - | - | - |
| 7F | 6·40 (5·93, 6·90) | - | - | - | - | - |
| 19A | 5·00 (3·00, 8·34) | - | - | - | - | - |

†: Only 1 study arm available for PCV7 (Multi-regions), no pooled estimate or heterogeneity.
‡: Only 1 study arm available for PCV20 (Western Pacific Region), no pooled estimate or heterogeneity.
Heterogeneity for PCV7 (Americas): $I^2$=98·5%, $\tau^2$=0·303, p <0·01; PCV7 (Europe): $I^2$=98·3%, $\tau^2$=0·274, p <0·01; PCV7 (South-East Asia): $I^2$=98·1%, $\tau^2$=0·241, p <0·01; PCV7 (Western Pacific): $I^2$=98·5%, $\tau^2$=0·272, p <0·01; for PCV13 (Africa): $I^2$=94·2%, $\tau^2$=0·309, p <0·01; for PCV13 (Americas): $I^2$=99·4%, $\tau^2$=0·580, p <0·01; PCV13 (Europe): $I^2$=99·3%, $\tau^2$=0·496, p <0·01; PCV13 (South-East Asia): $I^2$=99·0%, $\tau^2$=0·508, p <0·01; PCV13 (Western Pacific): $I^2$=99·5%, $\tau^2$=0·424, p <0·01; PCV13 (Multi-region): $I^2$=99·4%, $\tau^2$=0·515, p <0·01; for PCV15(Western Pacific): $I^2$=95·9%, $\tau^2$=0·306, p <0·01; PCV15 (Multi-region): $I^2$=99·4%, $\tau^2$=0·334, p <0·01; for PCV20 (Americas): $I^2$=99·6%, $\tau^2$=0·514, p <0·01; for PCV10-SII (Africa): $I^2$=99·1%, $\tau^2$=0·329, p <0·01.

**Table S17. Summary of the meta-analysis results of anti-pneumococcal IgG seroresponse rates (%) post "3+1" vaccination schedule by vaccine product and region**

| Serotype | IgG GMCs (95%CI) by WHO region (µg/mL)* | | | | | |
|---|---|---|---|---|---|---|
| | Africa | Americas | Europe | South East Asia | Western Pacific | Multi-regions |
| PCV7* | | | | | | |
| 4 | - | 96% (94%-98%) | 99% (99%-100%) | 100% (96%-100%) | 100% (99%-100%) | 85% (65%-97%) |
| 6B | - | 99% (98%-100%) | 98% (97%-99%) | 99% (98%-100%) | 100% (99%-100%) | 77% (55%-92%) |
| 9V | - | 99% (98%-100%) | 99% (98%-100%) | 99% (98%-100%) | 100% (99%-100%) | 85% (65%-97%) |
| 14 | - | 99% (98%-100%) | 99% (99%-100%) | 99% (98%-100%) | 100% (98%-100%) | 85% (65%-97%) |
| 18C | - | 98% (97%-99%) | 99% (97%-99%) | 99% (97%-100%) | 100% (99%-100%) | 85% (65%-97%) |
| 19F | - | 99% (98%-99%) | 99% (97%-99%) | 98% (96%-99%) | 100% (99%-100%) | 85% (65%-97%) |
| 23F | - | 99% (98%-99%) | 99% (98%-99%) | 99% (98%-100%) | 99% (95%-100%) | 85% (65%-97%) |
| PCV13† | | | | | | |
| 4 | 100% (95%-100%) | 98% (95%-99%) | 99% (99%-100%) | 100% (98%-100%) | 100% (99%-100%) | 98% (96%-99%) |
| 6B | 99% (92%-100%) | 100% (99%-100%) | 99% (99%-100%) | 98% (95%-99%) | 100% (99%-100%) | 99% (98%-100%) |
| 9V | 99% (92%-100%) | 98% (97%-99%) | 99% (99%-100%) | 99% (98%-100%) | 100% (98%-100%) | 99% (96%-100%) |
| 14 | 99% (92%-100%) | 100% (99%-100%) | 100% (99%-100%) | 98% (96%-99%) | 100% (99%-100%) | 100% (98%-100%) |
| 18C | 97% (90%-100%) | 99% (98%-99%) | 98% (97%-99%) | 99% (97%-100%) | 100% (99%-100%) | 99% (98%-100%) |
| 19F | 94% (86%-98%) | 99% (98%-99%) | 98% (97%-99%) | 98% (97%-99%) | 100% (99%-100%) | 100% (99%-100%) |
| 23F | 98% (91%-100%) | 98% (98%-99%) | 99% (98%-99%) | 98% (97%-99%) | 98% (97%-99%) | 99% (98%-99%) |
| 1 | 97% (90%-100%) | 97% (96%-98%) | 99% (98%-100%) | 99% (97%-99%) | 99% (99%-100%) | 99% (97%-100%) |
| 3 | 99% (92%-100%) | 87% (76%-93%) | 88% (82%-93%) | 95% (88%-98%) | 99% (98%-99%) | 80% (77%-83%) |
| 5 | 97% (90%-100%) | 98% (97%-99%) | 99% (98%-99%) | 100% (98%-100%) | 100% (99%-100%) | 98% (94%-100%) |
| 6A | 96% (88%-99%) | 100% (99%-100%) | 99% (98%-100%) | 98% (91%-99%) | 100% (99%-100%) | 100% (98%-100%) |
| 7F | 99% (92%-100%) | 100% (99%-100%) | 99% (98%-100%) | 99% (98%-100%) | 99% (98%-100%) | 100% (98%-100%) |

| Serotype | IgG GMCs (95%CI) by WHO region (μg/mL)[*] | | | | | |
|---|---|---|---|---|---|---|
| | Africa | Americas | Europe | South East Asia | Western Pacific | Multi-regions |
| 19A | 98% (92%-100%) | 100% (99%-100%) | 100% (99%-100%) | 99% (98%-100%) | 100% (99%-100%) | 100% (99%-100%) |
| PCV15[ǂ] | | | | | | |
| 4 | - | - | - | - | 100% (92%-100%) | - |
| 6B | - | - | - | - | 100% (92%-100%) | - |
| 9V | - | - | - | - | 100% (92%-100%) | - |
| 14 | - | - | - | - | 100% (92%-100%) | - |
| 18C | - | - | - | - | 100% (92%-100%) | - |
| 19F | - | - | - | - | 100% (92%-100%) | - |
| 23F | - | - | - | - | 100% (92%-100%) | - |
| 1 | - | - | - | - | 100% (92%-100%) | - |
| 3 | - | - | - | - | 98% (87%-100%) | - |
| 5 | - | - | - | - | 100% (92%-100%) | - |
| 6A | - | - | - | - | 100% (92%-100%) | - |
| 7F | - | - | - | - | 100% (92%-100%) | - |
| 19A | - | - | - | - | 100% (92%-100%) | - |
| 22F | - | - | - | - | 100% (92%-100%) | - |
| 33F | - | - | - | - | 100% (92%-100%) | - |
| PCV20[#] | | | | | | |
| 4 | - | 99% (98%-100%) | - | - | 92% (87%-95%) | - |
| 6B | - | 99% (98%-100%) | - | - | 100% (98%-100%) | - |
| 9V | - | 98% (97%-99%) | - | - | 100% (98%-100%) | - |
| 14 | - | 99% (98%-100%) | - | - | 100% (98%-100%) | - |
| 18C | - | 99% (98%-100%) | - | - | 100% (98%-100%) | - |
| 19F | - | 99% (98%-99%) | - | - | 100% (98%-100%) | - |
| 23F | - | 97% (96%-98%) | - | - | 100% (97%-100%) | - |
| 1 | - | 94% (92%-96%) | - | - | 99% (96%-100%) | - |
| 3 | - | 74% (70%-77%) | - | - | 92% (87%-95%) | - |
| 5 | - | 98% (97%-99%) | - | - | 100% (97%-100%) | - |
| 6A | - | 100% (99%-100%) | - | - | 100% (98%-100%) | - |
| 7F | - | 100% (99%-100%) | - | - | 100% (98%-100%) | - |
| 19A | - | 100% (99%-100%) | - | - | 100% (98%-100%) | - |
| 22F | - | 100% (99%-100%) | - | - | 100% (98%-100%) | - |
| 33F | - | 100% (99%-100%) | - | - | 100% (98%-100%) | - |
| 8 | - | 100% (99%-100%) | - | - | 100% (98%-100%) | - |
| 10A | - | 98% (96%-99%) | - | - | 100% (97%-100%) | - |
| 11A | - | 99% (98%-99%) | - | - | 100% (98%-100%) | - |
| 12F | - | 95% (93%-97%) | - | - | 99% (96%-100%) | - |
| 15B | - | 100% | - | - | 100% | - |

| Serotype | IgG GMCs (95%CI) by WHO region (μg/mL)* | | | | | |
|---|---|---|---|---|---|---|
| | Africa | Americas | Europe | South East Asia | Western Pacific | Multi-regions |
| | | (99%-100%) | | | (98%-100%) | |

*No Seroresponse rate data were reported for PCV10-SII study.
*: Only 1 study arm available for PCV7 (Multi-regions), no pooled estimate or heterogeneity.
†: Only 1 study arm available for PCV13 (African Region), no pooled estimate or heterogeneity.
‡: Only 1 study arm available for PCV15 (Western Pacific Region), no pooled estimate or heterogeneity.
#: Only 1 study arm available for PCV20 (Western Pacific Region) and PCV20 (Americas), no pooled estimate or heterogeneity.
Heterogeneity for PCV7 (Americas): $I^2$=57·2%, $\tau^2$=0·292, p <0·01; PCV7 (Europe): $I^2$=0%, $\tau^2$=0, p = 0·63; PCV7 (South-East Asia): $I^2$=0, $\tau^2$=0, p = 0·73; PCV7 (Western Pacific): $I^2$=21·8%, $\tau^2$=0·102, p = 0·22; for PCV13 (Americas): $I^2$=89·1%, $\tau^2$=1·007, p <0·01; PCV13 (Europe): $I^2$=83·4%, $\tau^2$=0·963, p <0·01; PCV13 (South-East Asia): $I^2$=60·9%, $\tau^2$=0·333, p <0·01; PCV13 (Western Pacific): $I^2$=12·7%, $\tau^2$=0·084, p = 0·25; PCV13 (Multi-region): $I^2$=85·5%, $\tau^2$=1·435, p <0·01.

**Table S18. Summary of the meta-analysis results of anti-pneumococcal IgG geometric mean concentrations (IgG GMCs, μg/mL) by vaccine product and timepoint**

| Serotype | IgG GMC (95%CI) by vaccine schedule (μg/mL) | | | |
|---|---|---|---|---|
| | Post 1-primary dose | Post 2-primary dose | Post 3-primary dose | Post booster |
| PCV7 | | | | |
| 4 | 0·68 (0·45, 1·02) | 2·66 (1·47, 4·79) | 3·25 (2·64, 3·99) | 4·41 (3·68, 5·28) |
| 6B | 0·16 (0·13, 0·19) | 0·47 (0·33, 0·66) | 1·99 (1·59, 2·49) | 8·83 (7·66, 10·17) |
| 9V | 0·40 (0·31, 0·53) | 1·93 (1·26, 2·94) | 2·37 (2·00, 2·81) | 3·83 (3·44, 4·26) |
| 14 | 0·97 (0·85, 1·12) | 3·38 (2·57, 4·45) | 6·30 (5·24, 7·58) | 11·54 (10·38, 12·83) |
| 18C | 0·54 (0·38, 0·77) | 1·70 (1·12, 2·59) | 2·69 (2·32, 3·13) | 3·24 (2·87, 3·66) |
| 19F | 1·05 (0·82, 1·33) | 3·54 (2·59, 4·82) | 3·44 (2·91, 4·07) | 5·13 (4·55, 5·78) |
| 23F | 0·23 (0·18, 0·29) | 0·78 (0·55, 1·11) | 2·18 (1·91, 2·50) | 5·66 (4·98, 6·42) |
| PCV13 | | | | |
| 4 | 0·56 (0·32, 0·98) | 1·84 (1·44, 2·37) | 2·14 (1·82, 2·51) | 3·70 (3·19, 4·29) |
| 6B | 0·14 (0·09, 0·21) | 0·38 (0·29, 0·51) | 1·75 (1·45, 2·11) | 7·63 (6·82, 8·55) |
| 9V | 0·29 (0·22, 0·37) | 1·35 (1·08, 1·68) | 1·68 (1·49, 1·90) | 3·29 (2·93, 3·68) |
| 14 | 0·62 (0·56, 0·69) | 3·76 (2·98, 4·74) | 5·26 (4·48, 6·18) | 9·88 (9·15, 10·66) |
| 18C | 0·47 (0·36, 0·60) | 1·43 (1·20, 1·71) | 1·99 (1·77, 2·25) | 3·02 (2·68, 3·40) |
| 19F | 0·75 (0·61, 0·92) | 4·41 (3·73, 5·22) | 2·92 (2·59, 3·29) | 8·38 (7·57, 9·26) |
| 23F | 0·14 (0·09, 0·21) | 0·68 (0·54, 0·84) | 1·57 (1·38, 1·79) | 4·33 (3·81, 4·93) |
| 1 | 0·81 (0·57, 1·16) | 2·08 (1·68, 2·58) | 2·28 (1·98, 2·63) | 4·33 (3·78, 4·95) |
| 3 | 0·77 (0·65, 0·92) | 0·75 (0·60, 0·94) | 0·92 (0·81, 1·05) | 0·95 (0·86, 1·05) |
| 5 | 0·45 (0·38, 0·54) | 1·10 (0·90, 1·34) | 1·49 (1·31, 1·71) | 3·11 (2·79, 3·47) |
| 6A | 0·24 (0·14, 0·44) | 1·40 (1·17, 1·68) | 2·27 (1·96, 2·63) | 8·65 (7·94, 9·42) |
| 7F | 0·76 (0·52, 1·12) | 2·53 (2·13, 3·01) | 3·08 (2·78, 3·41) | 5·35 (4·93, 5·79) |
| 19A | 0·46 (0·36, 0·59) | 2·29 (1·88, 2·77) | 2·70 (2·35, 3·10) | 8·43 (7·81, 9·10) |
| PCV15 | | | | |
| 4 | - | 1·57 (1·28, 1·93) | 1·71 (1·12, 2·60) | 1·56 (1·31, 1·86) |
| 6B | - | 0·43 (0·39, 0·47) | 1·74 (1·38, 2·19) | 5·63 (4·87, 6·52) |
| 9V | - | 1·38 (1·10, 1·73) | 1·97 (1·45, 2·66) | 2·73 (2·31, 3·24) |
| 14 | - | 4·65 (3·24, 6·67) | 5·92 (4·39, 7·99) | 6·75 (5·72, 7·97) |
| 18C | - | 1·18 (1·12, 1·23) | 1·66 (1·31, 2·09) | 2·65 (2·27, 3·10) |
| 19F | - | 2·71 (2·56, 2·87) | 2·50 (1·82, 3·43) | 4·72 (4·14, 5·37) |
| 23F | - | 0·73 (0·68, 0·78) | 1·47 (1·19, 1·81) | 2·23 (1·86, 2·67) |
| 1 | - | 1·34 (1·26, 1·43) | 1·58 (1·12, 2·25) | 1·73 (1·43, 2·09) |
| 3 | - | 0·98 (0·79, 1·22) | 1·35 (0·89, 2·04) | 1·08 (0·93, 1·25) |
| 5 | - | 1·01 (0·79, 1·28) | 1·87 (1·43, 2·43) | 2·85 (2·23, 3·64) |
| 6A | - | 0·66 (0·61, 0·70) | 1·60 (1·28, 1·99) | 4·18 (3·61, 4·82) |
| 7F | - | 1·86 (1·54, 2·23) | 2·57 (2·39, 2·76) | 3·76 (3·31, 4·27) |
| 19A | - | 1·71 (1·61, 1·81) | 1·84 (1·24, 2·73) | 4·84 (4·11, 5·71) |
| 22F | - | 2·96 (2·59, 3·38) | 5·45 (4·44, 6·70) | 8·13 (6·78, 9·75) |
| 33F | - | 0·30 (0·28, 0·33) | 1·73 (1·50, 1·99) | 4·67 (3·89, 5·62) |
| PCV20* | | | | |
| 4 | - | 0·55 (0·50, 0·61) | 1·20 (0·74, 1·96) | 5·32 (3·76, 7·52) |
| 6B | - | 0·03 (0·03, 0·04) | 0·53 (0·39, 0·72) | 4·73 (2·98, 7·52) |
| 9V | - | 0·45 (0·40, 0·51) | 1·15 (0·85, 1·55) | 4·38 (3·44, 5·59) |
| 14 | - | 1·05 (0·94, 1·18) | 2·53 (2·03, 3·14) | 6·69 (4·80, 9·33) |
| 18C | - | 0·69 (0·62, 0·77) | 1·46 (1·29, 1·64) | 3·74 (2·79, 5·02) |
| 19F | - | 2·21 (2·04, 2·40) | 2·05 (1·50, 2·81) | 6·71 (5·30, 8·51) |
| 23F | - | 0·13 (0·12, 0·15) | 1·00 (0·76, 1·30) | 4·64 (2·96, 7·27) |
| 1 | - | 0·57 (0·52, 0·62) | 0·92 (0·71, 1·20) | 2·07 (1·51, 2·82) |
| 3 | - | 0·41 (0·38, 0·45) | 0·55 (0·28, 1·09) | 0·84 (0·60, 1·17) |
| 5 | - | 0·34 (0·30, 0·38) | 0·84 (0·65, 1·09) | 2·38 (1·72, 3·29) |
| 6A | - | 0·45 (0·40, 0·52) | 1·93 (1·62, 2·32) | 10·73 (8·02, 14·36) |
| 7F | - | 1·02 (0·94, 1·10) | 1·77 (1·46, 2·16) | 4·52 (3·58, 5·71) |
| 19A | - | 0·67 (0·61, 0·74) | 1·13 (0·55, 2·34) | 5·17 (3·68, 7·25) |
| 22F | - | 2·25 (2·06, 2·45) | 4·19 (3·67, 4·78) | 11·89 (9·55, 14·82) |

| Serotype | IgG GMC (95%CI) by vaccine schedule (μg/mL) | | | |
|---|---|---|---|---|
| | Post 1-primary dose | Post 2-primary dose | Post 3-primary dose | Post booster |
| 33F | - | 0·31 (0·28, 0·34) | 1·74 (1·37, 2·19) | 7·47 (5·12, 10·90) |
| 8 | - | 1·62 (1·51, 1·74) | 2·31 (1·62, 3·29) | 4·01 (3·08, 5·23) |
| 10A | - | 0·16 (0·14, 0·18) | 0·98 (0·47, 2·07) | 6·98 (5·15, 9·45) |
| 11A | - | 1·62 (1·50, 1·75) | 2·41 (1·11, 5·26) | 4·55 (3·51, 5·90) |
| 12F | - | 0·15 (0·13, 0·17) | 0·69 (0·53, 0·90) | 2·04 (1·72, 2·44) |
| 15B | - | 3·33 (3·00, 3·70) | 5·57 (4·33, 7·17) | 16·00 (12·31, 20·80) |
| PCV10-SII[‡] | | | | |
| 6B | - | 1·82 (1·48, 2·23) | 1·34 (1·05, 1·72) | 10·76 (8·16, 14·20) |
| 9V | - | 1·93 (1·74, 2·16) | 1·20 (0·99, 1·46) | 2·47 (1·70, 3·60) |
| 14 | - | 4·03 (3·50, 4·64) | 5·18 (4·91, 5·47) | 7·49 (6·58, 8·52) |
| 19F | - | 5·45 (4·94, 6·01) | 4·24 (3·85, 4·66) | 8·09 (5·73, 11·43) |
| 23F | - | 2·21 (1·92, 2·56) | 1·58 (1·51, 1·66) | 4·54 (3·96, 5·21) |
| 1 | - | 3·63 (3·32, 3·98) | 3·61 (2·54, 5·15) | 6·76 (5·28, 8·65) |
| 5 | - | 1·19 (1·10, 1·28) | 1·84 (1·46, 2·31) | 1·64 (1·19, 2·26) |
| 6A | - | 1·19 (1·00, 1·41) | 1·00 (0·95, 1·06) | 7·49 (4·79, 11·72) |
| 7F | - | 3·46 (3·09, 3·88) | 2·58 (1·92, 3·48) | 6·48 (6·09, 6·90) |
| 19A | - | 1·75 (1·57, 1·96) | 1·63 (1·56, 1·71) | 6·13 (3·82, 9·82) |

[*]: Only 1 study arm available for PCV20 (Post 2-primary), no pooled estimate or heterogeneity.

[‡]: Only 1 study arm available for PCV10-SII (Post 2-primary), no pooled estimate or heterogeneity.

Heterogeneity for PCV7 (post 1-primary): $I^2$=98·4%, $\tau^2$=0·476, p <0·01; for PCV7 (post 2-primary): $I^2$=99·0%, $\tau^2$=0·918, p <0·01; for PCV7 (post 3-primary): $I^2$=98·7%, $\tau^2$=0·410, p <0·01; for PCV7 (post booster): $I^2$=98·6%, $\tau^2$=0·348, p <0·01; for PCV13 (post 1-primary): $I^2$=98·1%, $\tau^2$=0·469, p <0·01; for PCV13 (post 2-primary): $I^2$=99·2%, $\tau^2$=0·660, p <0·01; for PCV13 (post 3-primary): $I^2$=99·2%, $\tau^2$=0·400, p <0·01; for PCV13 (post booster): $I^2$=99·3%, $\tau^2$=0·582, p <0·01; for PCV15 (post 2-primary): $I^2$=99·5%, $\tau^2$=0·511, p <0·01; for PCV15 (post 3-primary): $I^2$=98·9%, $\tau^2$=0·314, p <0·01; for PCV15 (post booster): $I^2$=99·4%, $\tau^2 =$ 0.361, p <0.01; for PCV20 (post 3-primary): $I^2 =$ 99.4%, $\tau^2 =$ 0.462, p <0.01; for PCV20 (post booster): $I^2$=99·6%, $\tau^2$=0·508, p <0·01; for PCV10-SII (post 3-primary): $I^2$=99·7%, $\tau^2$=0·298, p <0·01; for PCV10-SII (post booster): $I^2$=99·1%, $\tau^2$=0·354, p <0·01.

**Table S19. Summary of the meta-analysis results of anti-pneumococcal IgG seroresponse rates (%) by vaccine product and timepoint**

| Serotype | Seroresponse rate (95%CI) by vaccine schedule (%) | | | |
|---|---|---|---|---|
| | Post 1-primary dose | Post 2-primary dose | Post 3-primary dose | Post booster |
| PCV7 | | | | |
| 4 | 75% (58%-86%) | 96% (94%-97%) | 98% (98%-99%) | 99% (97%-99%) |
| 6B | 23% (19%-27%) | 61% (48%-72%) | 89% (85%-93%) | 99% (98%-99%) |
| 9V | 56% (49%-62%) | 94% (91%-96%) | 98% (97%-98%) | 99% (98%-100%) |
| 14 | 84% (73%-91%) | 96% (94%-97%) | 97% (96%-98%) | 99% (98%-100%) |
| 18C | 67% (54%-78%) | 93% (88%-96%) | 97% (97%-98%) | 99% (98%-99%) |
| 19F | 91% (81%-96%) | 96% (95%-98%) | 97% (96%-98%) | 99% (98%-99%) |
| 23F | 31% (20%-44%) | 77% (67%-84%) | 94% (92%-95%) | 99% (98%-99%) |
| PCV13 | | | | |
| 4 | 68% (46%-84%) | 96% (94%-97%) | 97% (96%-98%) | 99% (98%-99%) |
| 6B | 8% (4%-18%) | 55% (46%-64%) | 90% (87%-92%) | 99% (98%-99%) |
| 9V | 35% (27%-44%) | 92% (88%-94%) | 96% (95%-97%) | 99% (98%-99%) |
| 14 | 74% (70%-78%) | 96% (94%-97%) | 98% (97%-98%) | 99% (99%-99%) |
| 18C | 64% (51%-75%) | 91% (88%-93%) | 96% (96%-97%) | 99% (98%-99%) |
| 19F | 85% (77%-91%) | 98% (97%-99%) | 98% (97%-98%) | 99% (98%-99%) |
| 23F | 12% (8%-16%) | 72% (66%-77%) | 91% (90%-93%) | 98% (97%-98%) |
| 1 | 76% (70%-82%) | 96% (95%-97%) | 97% (96%-98%) | 99% (98%-99%) |
| 3 | 84% (78%-88%) | 87% (79%-92%) | 90% (87%-93%) | 90% (87%-93%) |
| 5 | 60% (54%-67%) | 90% (87%-93%) | 94% (92%-95%) | 99% (98%-99%) |
| 6A | 25% (10%-50%) | 88% (85%-91%) | 96% (94%-97%) | 99% (98%-99%) |
| 7F | 79% (67%-87%) | 98% (97%-99%) | 99% (98%-99%) | 99% (99%-99%) |
| 19A | 61% (50%-71%) | 96% (94%-97%) | 98% (98%-99%) | 99% (99%-100%) |
| PCV15 | | | | |
| 4 | - | 96% (90%-98%) | 97% (96%-98%) | 96% (95%-97%) |
| 6B | - | 58% (55%-61%) | 90% (85%-94%) | 98% (96%-99%) |
| 9V | - | 92% (85%-96%) | 97% (96%-98%) | 99% (98%-100%) |
| 14 | - | 97% (96%-98%) | 98% (97%-99%) | 99% (99%-100%) |
| 18C | - | 93% (91%-94%) | 97% (96%-98%) | 99% (98%-100%) |
| 19F | - | 98% (97%-99%) | 98% (95%-100%) | 100% (99%-100%) |
| 23F | - | 77% (74%-80%) | 92% (89%-95%) | 97% (96%-98%) |
| 1 | - | 96% (94%-98%) | 97% (96%-97%) | 97% (95%-98%) |
| 3 | - | 95% (91%-98%) | 95% (93%-96%) | 92% (91%-94%) |
| 5 | - | 89% (79%-94%) | 96% (95%-97%) | 99% (98%-100%) |
| 6A | - | 75% (71%-79%) | 94% (86%-98%) | 99% (98%-99%) |
| 7F | - | 98% (97%-99%) | 99% (98%-99%) | 100% (99%-100%) |
| 19A | - | 95% (93%-97%) | 97% (94%-99%) | 99% (99%-100%) |
| 22F | - | 97% (94%-98%) | 99% (98%-99%) | 100% (99%-100%) |

| Serotype | Seroresponse rate (95%CI) by vaccine schedule (%) | | | |
|---|---|---|---|---|
| | Post 1-primary dose | Post 2-primary dose | Post 3-primary dose | Post booster |
| 33F | - | 49% (46%-52%) | 89% (83%-94%) | 99% (98%-100%) |
| PCV20* | | | | |
| 4 | - | 69% (65%-72%) | 89% (76%-95%) | 98% (91%-100%) |
| 6B | - | 21% (17%-24%) | 86% (82%-89%) | 99% (98%-99%) |
| 9V | - | 60% (56%-64%) | 88% (80%-93%) | 99% (98%-99%) |
| 14 | - | 79% (75%-82%) | 94% (92%-96%) | 98% (94%-100%) |
| 18C | - | 71% (67%-75%) | 93% (91%-94%) | 99% (98%-99%) |
| 19F | - | 94% (92%-96%) | 98% (95%-99%) | 99% (98%-100%) |
| 23F | - | 24% (20%-27%) | 82% (74%-88%) | 98% (94%-99%) |
| 1 | - | 71% (67%-74%) | 87% (78%-92%) | 97% (93%-98%) |
| 3 | - | 58% (54%-62%) | 77% (37%-95%) | 84% (70%-92%) |
| 5 | - | 63% (59%-67%) | 88% (80%-93%) | 98% (97%-99%) |
| 6A | - | 60% (55%-64%) | 94% (92%-95%) | 99% (98%-100%) |
| 7F | - | 88% (85%-90%) | 97% (93%-99%) | 100% (99%-100%) |
| 19A | - | 92% (90%-94%) | 98% (96%-99%) | 100% (99%-100%) |
| 22F | - | 94% (92%-96%) | 98% (97%-99%) | 99% (99%-100%) |
| 33F | - | 47% (43%-51%) | 90% (86%-93%) | 99% (98%-100%) |
| 8 | - | 96% (95%-98%) | 99% (95%-100%) | 99% (99%-100%) |
| 10A | - | 29% (25%-33%) | 78% (59%-90%) | 98% (96%-99%) |
| 11A | - | 94% (92%-96%) | 97% (83%-99%) | 99% (98%-99%) |
| 12F | - | 30% (27%-34%) | 75% (65%-83%) | 97% (94%-98%) |
| 15B | - | 94% (92%-96%) | 98% (97%-99%) | 100% (99%-100%) |
| PCV10-SII‡ | | | | |
| 6B | - | 85% (79%-89%) | 84% (71%-92%) | 100% (98%-100%) |
| 9V | - | 95% (91%-97%) | 95% (94%-96%) | 99% (96%-100%) |
| 14 | - | 99% (96%-100%) | 99% (98%-99%) | 98% (96%-100%) |
| 19F | - | 100% (98%-100%) | 98% (97%-99%) | 100% (98%-100%) |
| 23F | - | 96% (92%-98%) | 93% (91%-96%) | 98% (95%-99%) |
| 1 | - | 100% (97%-100%) | 100% (99%-100%) | 100% (98%-100%) |
| 5 | - | 97% (93%-99%) | 98% (98%-99%) | 98% (94%-99%) |
| 6A | - | 83% (78%-88%) | 82% (80%-83%) | 98% (95%-99%) |
| 7F | - | 100% (97%-100%) | 98% (96%-99%) | 100% (98%-100%) |
| 19A | - | 98% (95%-99%) | 94% (91%-97%) | 100% (98%-100%) |

*: Only 1 study arm available for PCV20 (Post 2-primary), no pooled estimate or heterogeneity.

‡: Only 1 study arm available for PCV10-SII (Post 2-primary), no pooled estimate or heterogeneity.

Heterogeneity for PCV7 (post 1-primary): $I^2$=96·8%, $\tau^2$=1·531, p <0·01; for PCV7 (post 2-primary): $I^2$=94·6%, $\tau^2$=1·665, p <0·01; for PCV7 (post 3-primary): $I^2$=82·7%, $\tau^2$=0·924, p <0·01; for PCV7 (post booster): $I^2$=98·6%, $\tau^2$=0·348, p <0·01; for PCV13 (post 1-primary): $I^2$=97·3%, $\tau^2$=2·097, p <0·01; for PCV13 (post 2-primary): $I^2$=95·7%, $\tau^2$=1·727, p <0·01; for PCV13 (post 3-primary): $I^2$=91·0%, $\tau^2$=1·199, p <0·01; for PCV13 (post booster): $I^2$=99·3%, $\tau^2$=0·582, p <0·01; for PCV15 (post 2-primary): $I^2$=98·6%, $\tau^2$=1·650, p <0·01; for PCV15 (post 3-primary): $I^2$=88·8%, $\tau^2$=0·610, p <0·01; for PCV15 (post booster): $I^2$=99·4%, $\tau^2$=0·361, p <0·01; for PCV20 (post 3-primary): $I^2$=99·4%, $\tau^2$=0·462, p <0·01; for PCV20 (post booster): $I^2$=97·0%, $\tau^2$=1·334, p <0·01; for PCV10-SII (post 3-primary): $I^2$=99·7%, $\tau^2$=0·298, p <0·01; for PCV10-SII (post booster): $I^2$=97·7%, $\tau^2$=1·462, p <0·01.

### Supplementary figures

#### Figure S1. Temporal distribution of study arms based on study start year

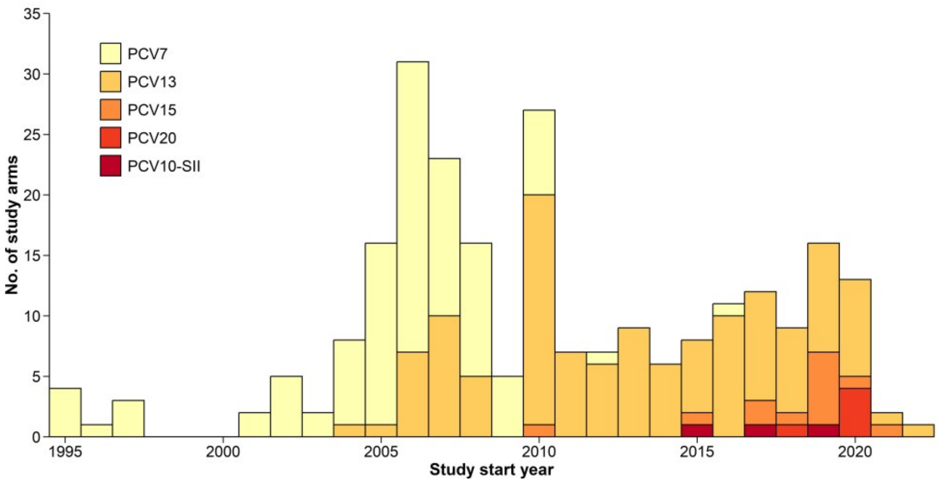

#### Figure S2. World map of study sites by number of study arms

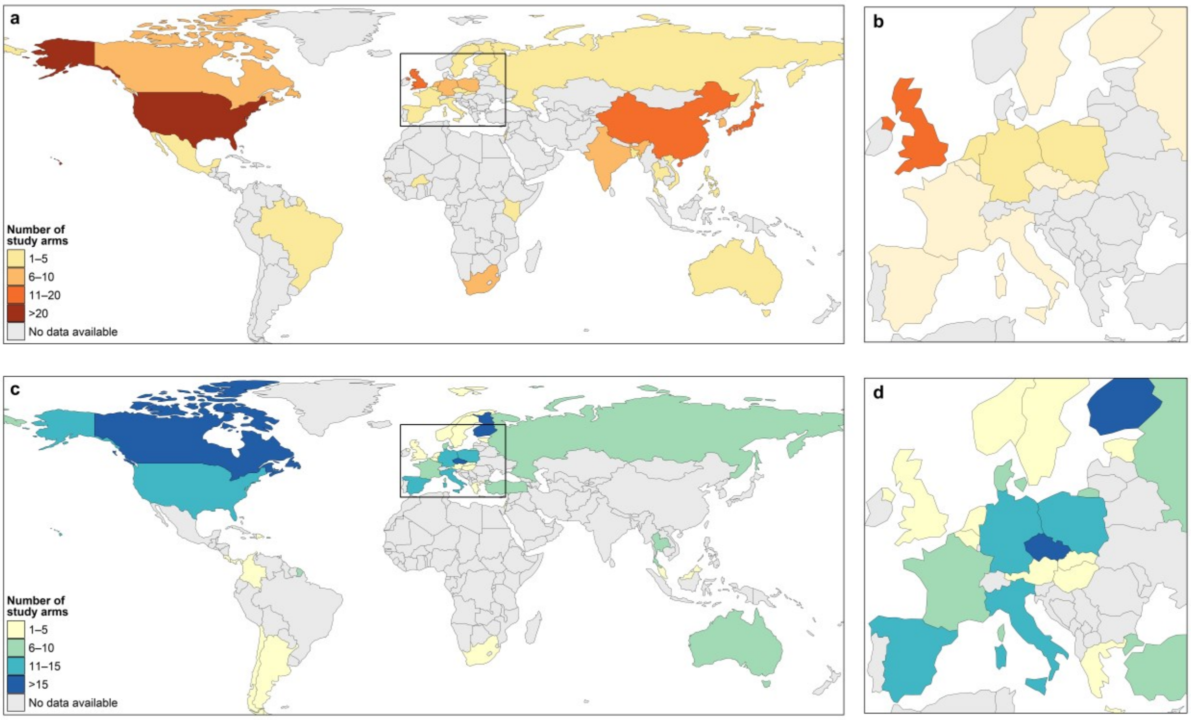

(a) Global distribution of single-country study arms by country. (b) Zoomed-in view of (a) showing single-country study arms in Europe. (c) Global distribution of multi-country study arms by country. (d) Zoomed-in view of (c) showing multi-country study arms in Europe.

***Note***: "multi-country" indicates study arms conducted across multiple sites involving more than one country. In multi-country studies, each study arm was counted for every country involved. In contrast, study arms conducted in a single country were counted only once.

**Figure S3. Pneumococcal post-childhood-schedule IgG GMCs (µg/mL) by serotype and vaccine product for all vaccine-covered serotypes**

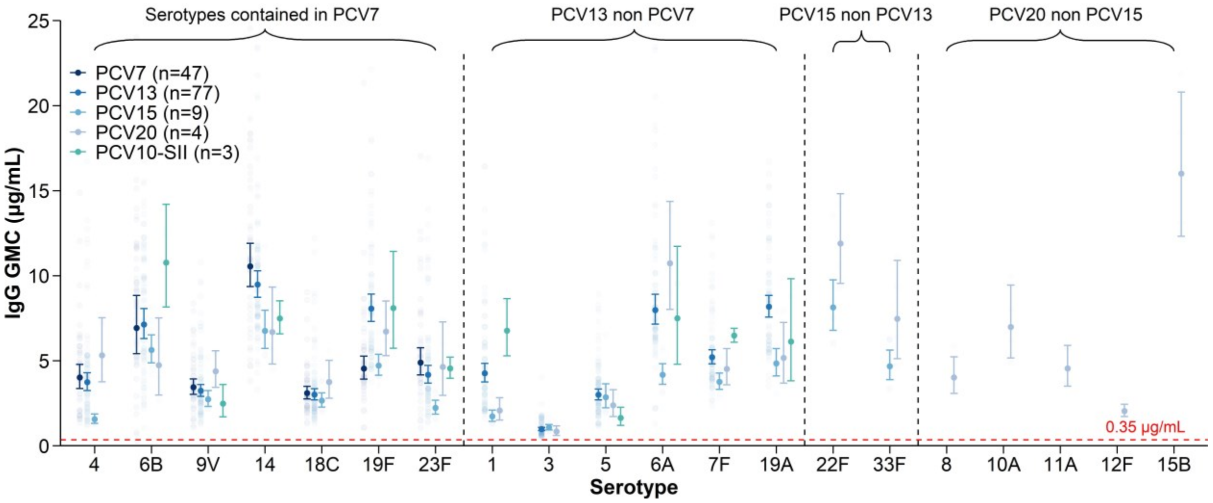

IgG GMCs post-childhood-schedule with different vaccine products.
***Note***: Numbers in parentheses following each vaccine name in the legend indicate the number of study arms included in the meta-analysis contributing to the pooled estimates for that vaccine.

**Figure S4. Pneumococcal post-childhood-schedule IgG GMCs (µg/mL) by serotype and vaccine product for different assays**

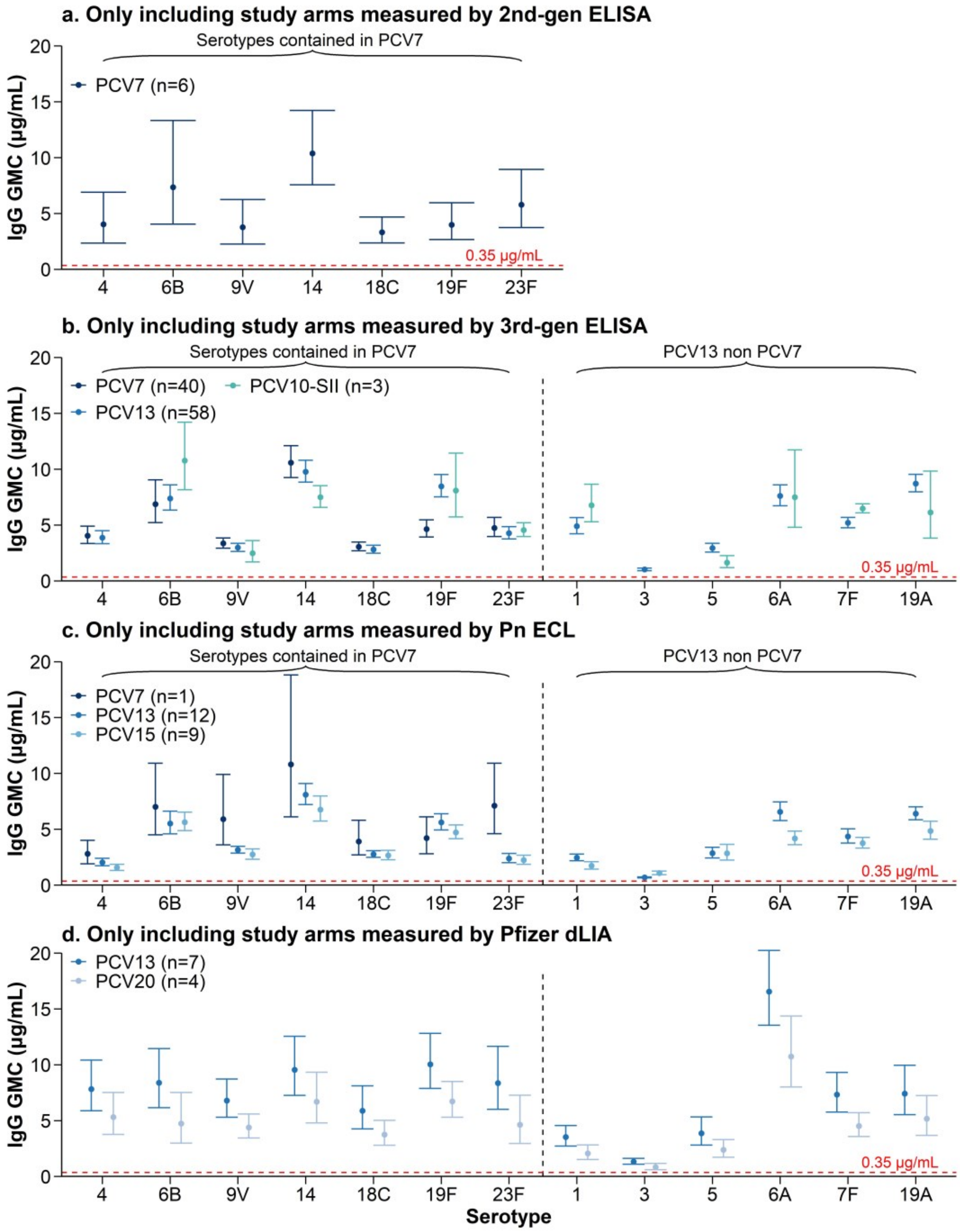

IgG GMCs post-childhood-schedule with different vaccine products measured by different laboratory methods. Panel (a) Study arms used second-generation ELISA to measure IgG response, (b) Study arms used third-generation ELISA (WHO reference ELISA) to measure IgG response, (c) Study arms used Pn ECL to measure IgG response, and (d) Study arms used Pfizer dLIA to measure IgG response.

***Note***: Numbers in parentheses following each vaccine name in the legend indicate the number of study arms included in the meta-analysis contributing to the pooled estimates for that vaccine.

**Figure S5. Pneumococcal post-childhood-schedule seroresponse rates (%)by serotype and vaccine product**

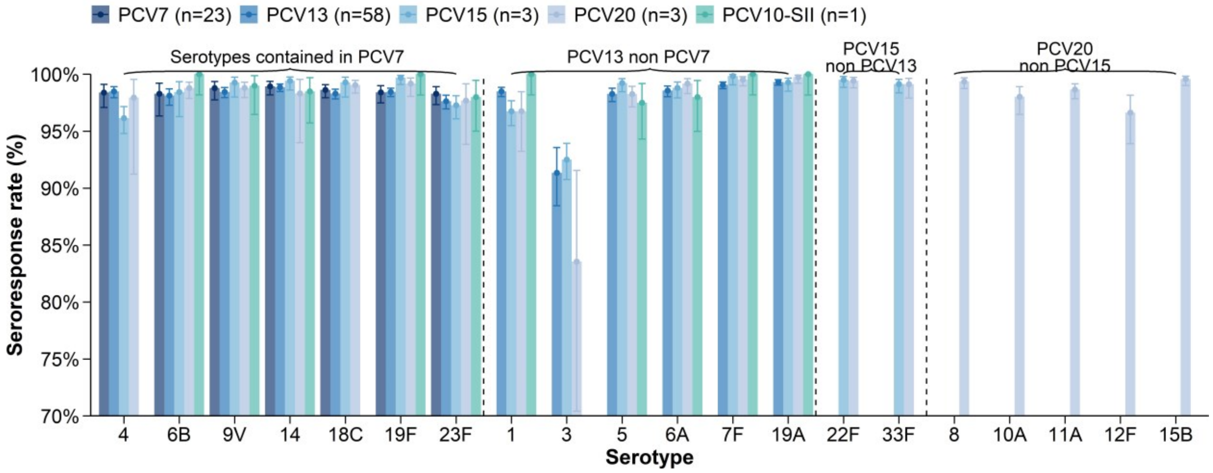

Seroresponse rates post-childhood-schedule with different vaccine products. The seroresponse rate indicates the proportion of participants achieving a predefined antibody response threshold. The y-axes in all panels are restricted to the range of 70–100% to highlight differences across vaccines.

***Note***: Numbers in parentheses following each vaccine name in the legend indicate the number of study arms included in the meta-analysis contributing to the pooled estimates for that vaccine.

**Figure S6. Pneumococcal post-childhood-schedule IgG GMC (µg/mL) by serotype, vaccine product and vaccine schedule**

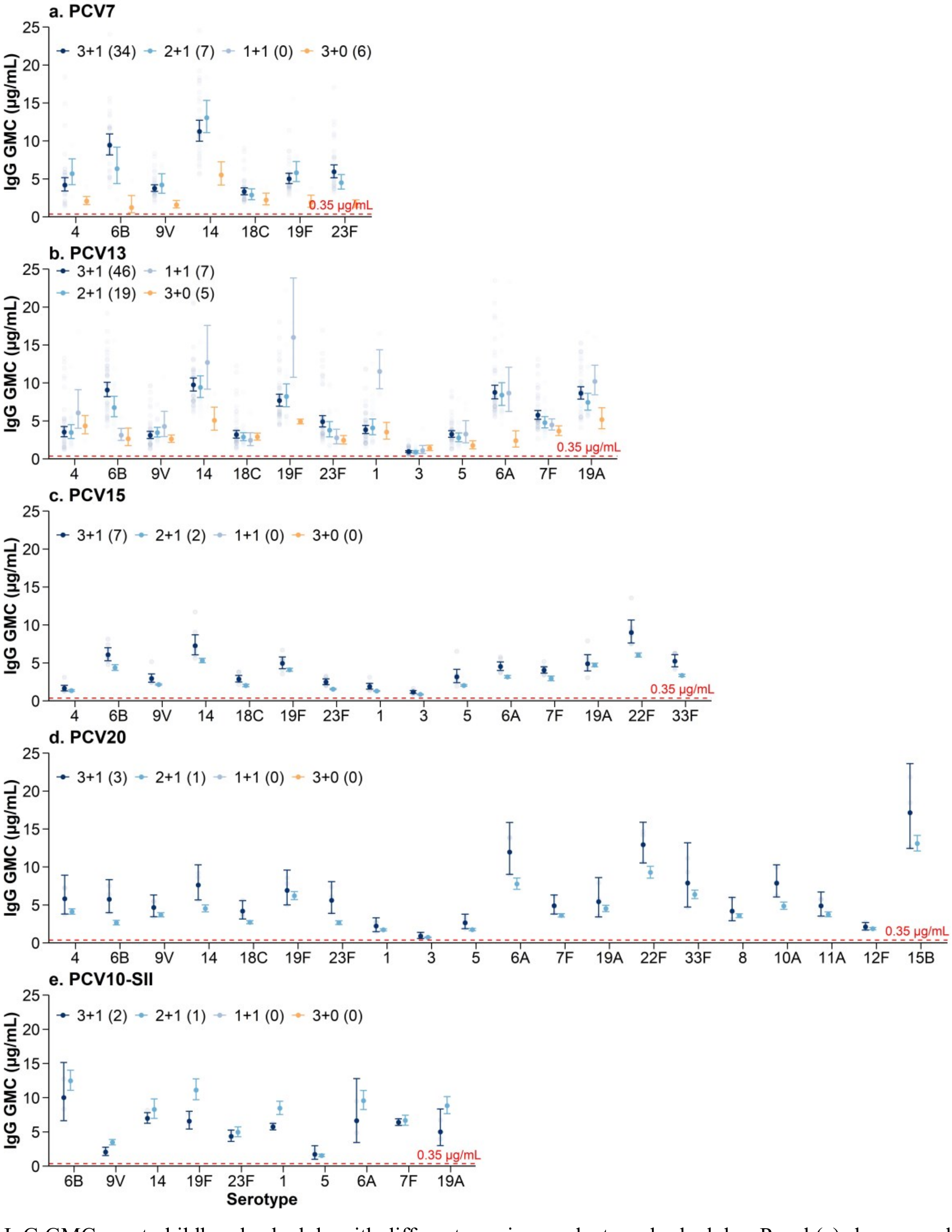

IgG GMCs post-childhood-schedule with different vaccine products and schedules. Panel (a) shows results for PCV7, (b) for PCV13, (c) for PCV15, (d) for PCV20, and (e) for PCV10-SII.

***Note***: Numbers in parentheses following each vaccine name in the legend indicate the number of study arms included in the meta-analysis contributing to the pooled estimates for that vaccine.

**Figure S7. Pneumococcal post-childhood-schedule seroresponse rates (%) by serotype, vaccine product and vaccine schedule**

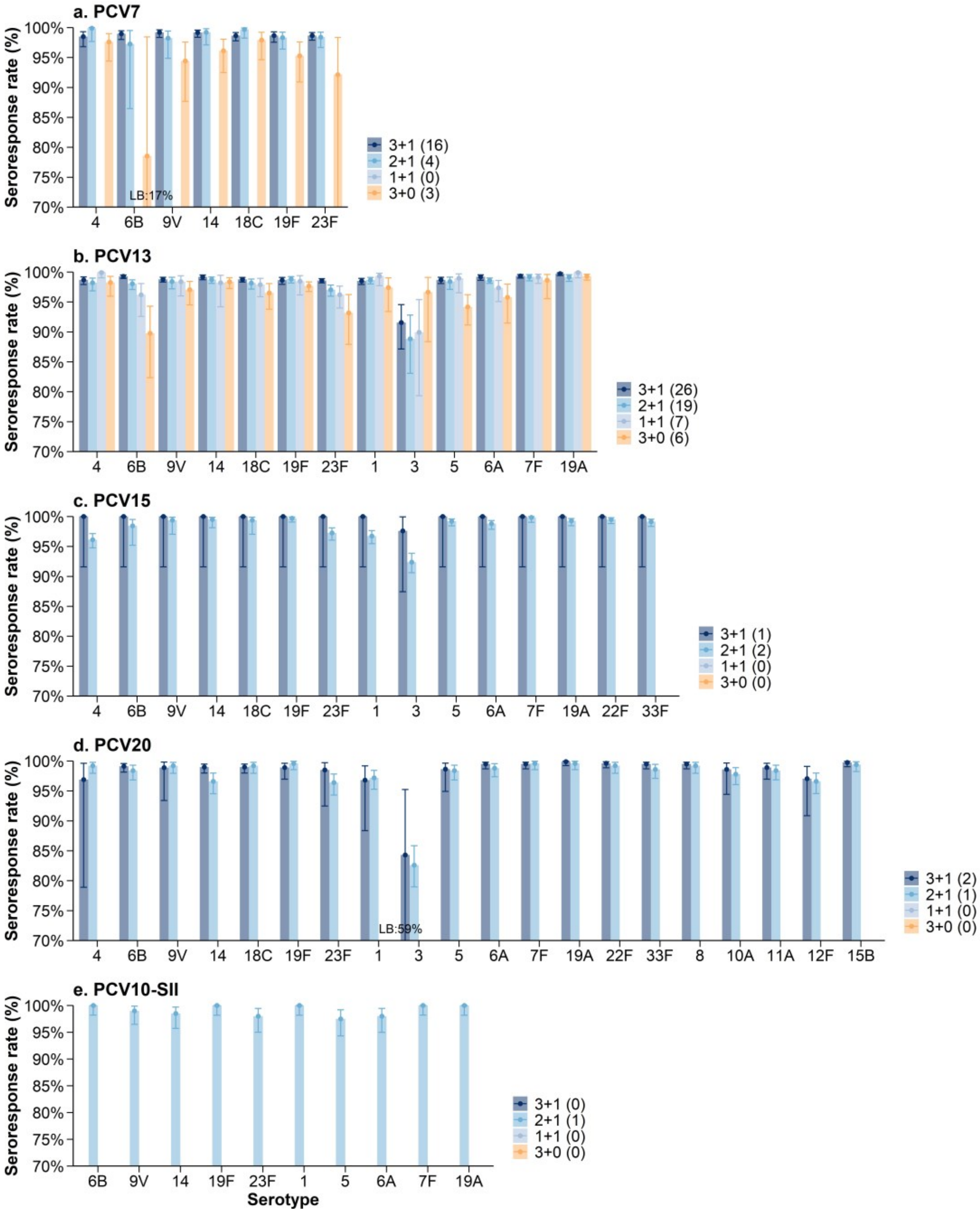

Seroresponse rates post-childhood-schedule with different vaccine products and schedules. Panel (a) shows results for PCV7, (b) for PCV13, (c) for PCV15, (d) for PCV20, and (e) for PCV10-SII. The seroresponse rate indicates the proportion of participants achieving a predefined antibody response threshold. The y-axes in all panels are restricted to the range of 80–100% to highlight differences across vaccines. LB denotes lower bound. ***Note***: Numbers in parentheses following each vaccine name in the legend indicate the number of study arms included in the meta-analysis contributing to the pooled estimates for that vaccine.

**Figure S8. Pneumococcal IgG GMC (µg/mL) post "3+1" vaccination schedule by serotype, vaccine product and region**

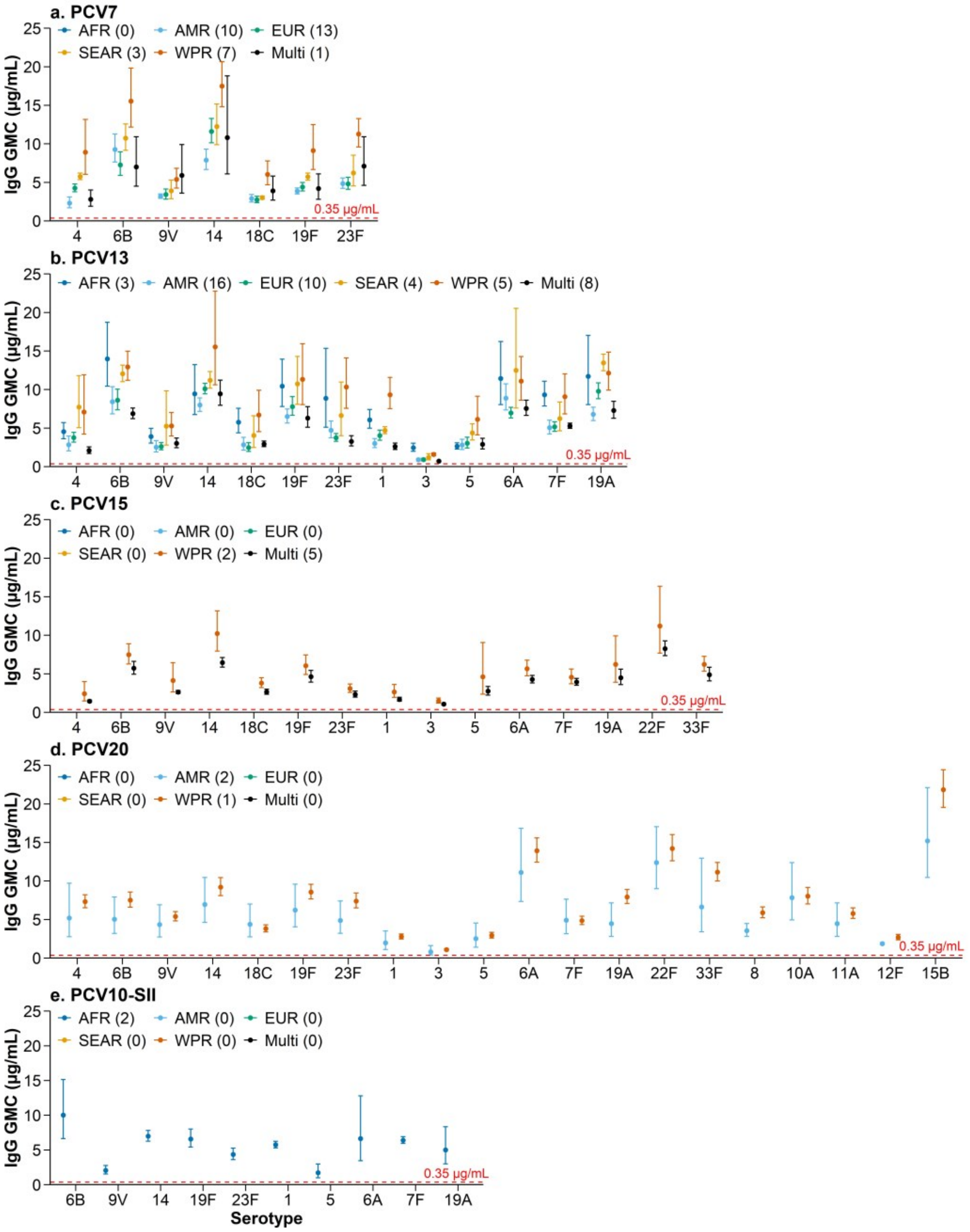

IgG GMCs post-childhood-schedule with different vaccine products using a 3+1 schedule, stratified by WHO regions. Panel (a) shows results for PCV7, (b) for PCV13, (c) for PCV15, (d) for PCV20, and (e) for PCV10-SII.

***Note***: "Multi region" indicates study arms conducted across multiple sites involving more than one WHO region. Numbers in parentheses following each vaccine name in the legend indicate the number of study arms included in the meta-analysis contributing to the pooled estimates for that vaccine.

***Abbreviations:*** AFR: African Region; AMR: Region of the Americas; EUR: European Region; SEAR: South-East Asia Region; WPR: Western Pacific Region; Multi: Multi-regions.

**Figure S9. Pneumococcal seroresponse rates (%) post "3+1" vaccination schedule by serotype, vaccine product and region**

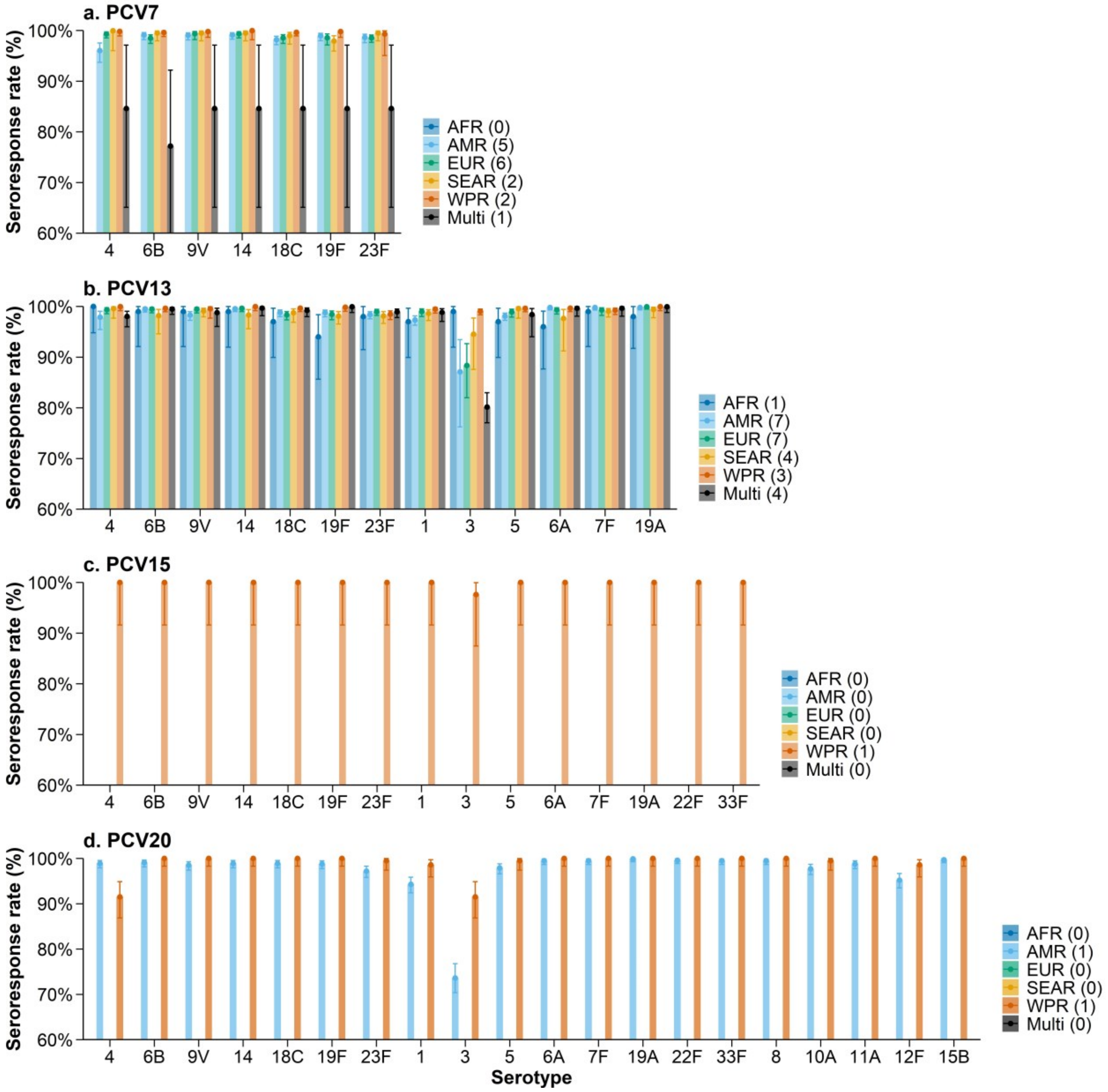

Seroresponse rates post-childhood-schedule with different vaccine products using a 3+1 schedule, stratified by WHO regions. Panel (a) shows results for PCV7, (b) for PCV13, (c) for PCV15, and (d) for PCV20. The seroresponse rate indicates the proportion of participants achieving a predefined antibody response threshold. The y-axes in all panels are restricted to the range of 80–100% to highlight differences across vaccines.

***Note***: "Multi region" indicates study arms conducted across multiple sites involving more than one WHO region. Numbers in parentheses following each vaccine name in the legend indicate the number of study arms included in the meta-analysis contributing to the pooled estimates for that vaccine. No seroresponse rate data were reported for PCV10-SII study.

***Abbreviations:*** AFR: African Region; AMR: Region of the Americas; EUR: European Region; SEAR: South-East Asia Region; WPR: Western Pacific Region; Multi: Multi regions.

**Figure S10. Serotype-specific pneumococcal IgG GMCs (μg/mL) by vaccine product and timepoint**

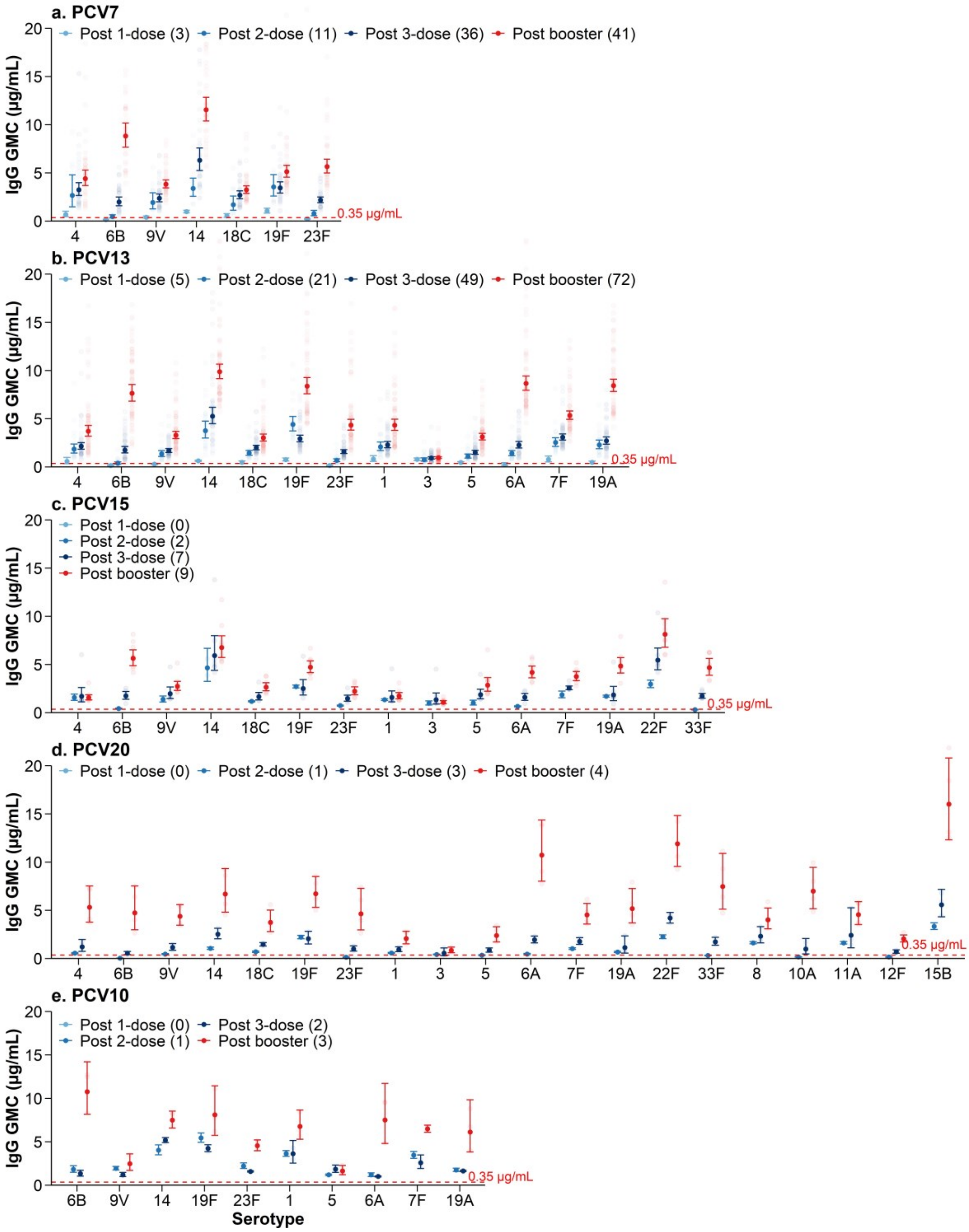

IgG GMCs post PCV vaccination by different vaccine products and timepoints. Panel (a) shows results for PCV7, (b) for PCV13, (c) for PCV15, (d) for PCV20, and (e) for PCV10-SII.
***Note***: Numbers in parentheses following each vaccine name in the legend indicate the number of study arms included in the meta-analysis contributing to the pooled estimates for that vaccine.

**Figure S11. Serotype-specific pneumococcal seroresponse rates (%) by vaccine product and timepoint**

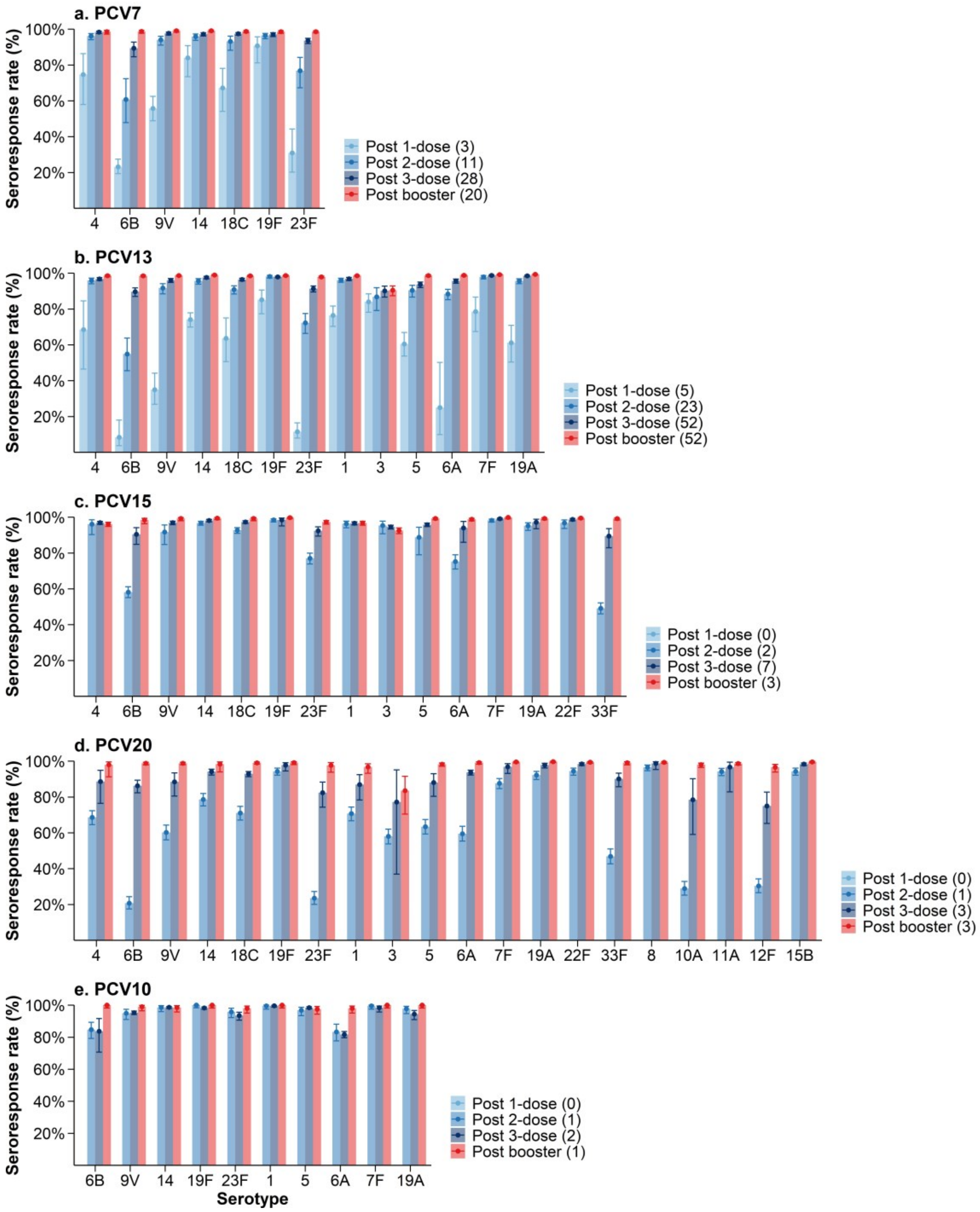

Seroresponse rates post PCV vaccination by different vaccine products and timepoints. Panel (a) shows results for PCV7, (b) for PCV13, (c) for PCV15, (d) for PCV20, and (e) for PCV10-SII. The seroresponse rate indicates the proportion of participants achieving a predefined antibody response threshold.
***Note***: Numbers in parentheses following each vaccine name in the legend indicate the number of study arms included in the meta-analysis contributing to the pooled estimates for that vaccine.

**Figure S12. Sensitivity analysis of pneumococcal post-childhood-schedule IgG GMCs (µg/mL) by serotype and vaccine product (c.f. Fig 2 and Fig S3 which excludes these studies)**

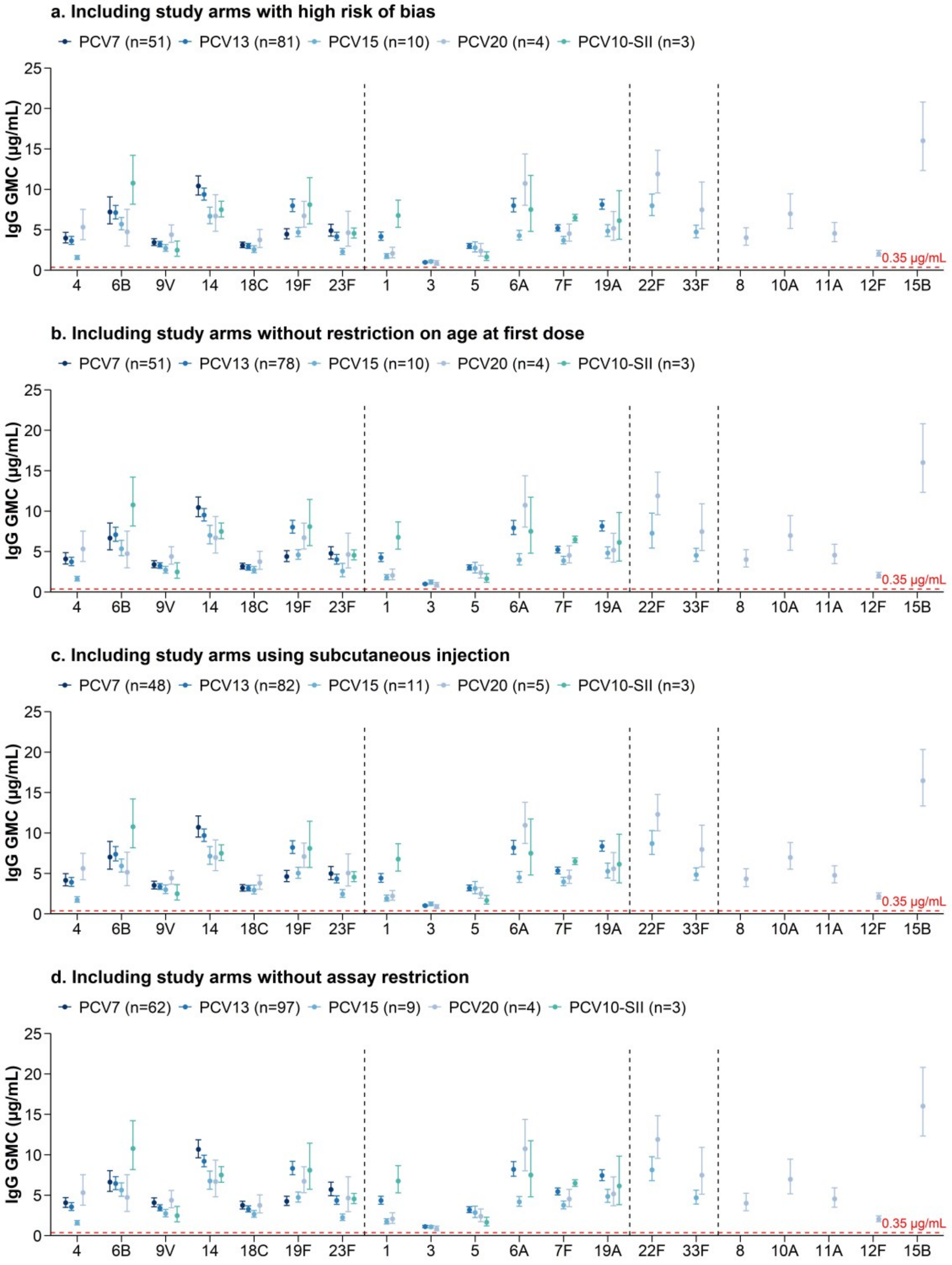

a

***Note***: Numbers in parentheses following each vaccine name in the legend indicate the number of study arms included in the meta-analysis contributing to the pooled estimates for that vaccine.

**Figure S13. Sensitivity analysis of pneumococcal post-childhood-schedule seroresponse rates (%) by serotype and vaccine product (c.f. Fig S5 which excludes these studies)**

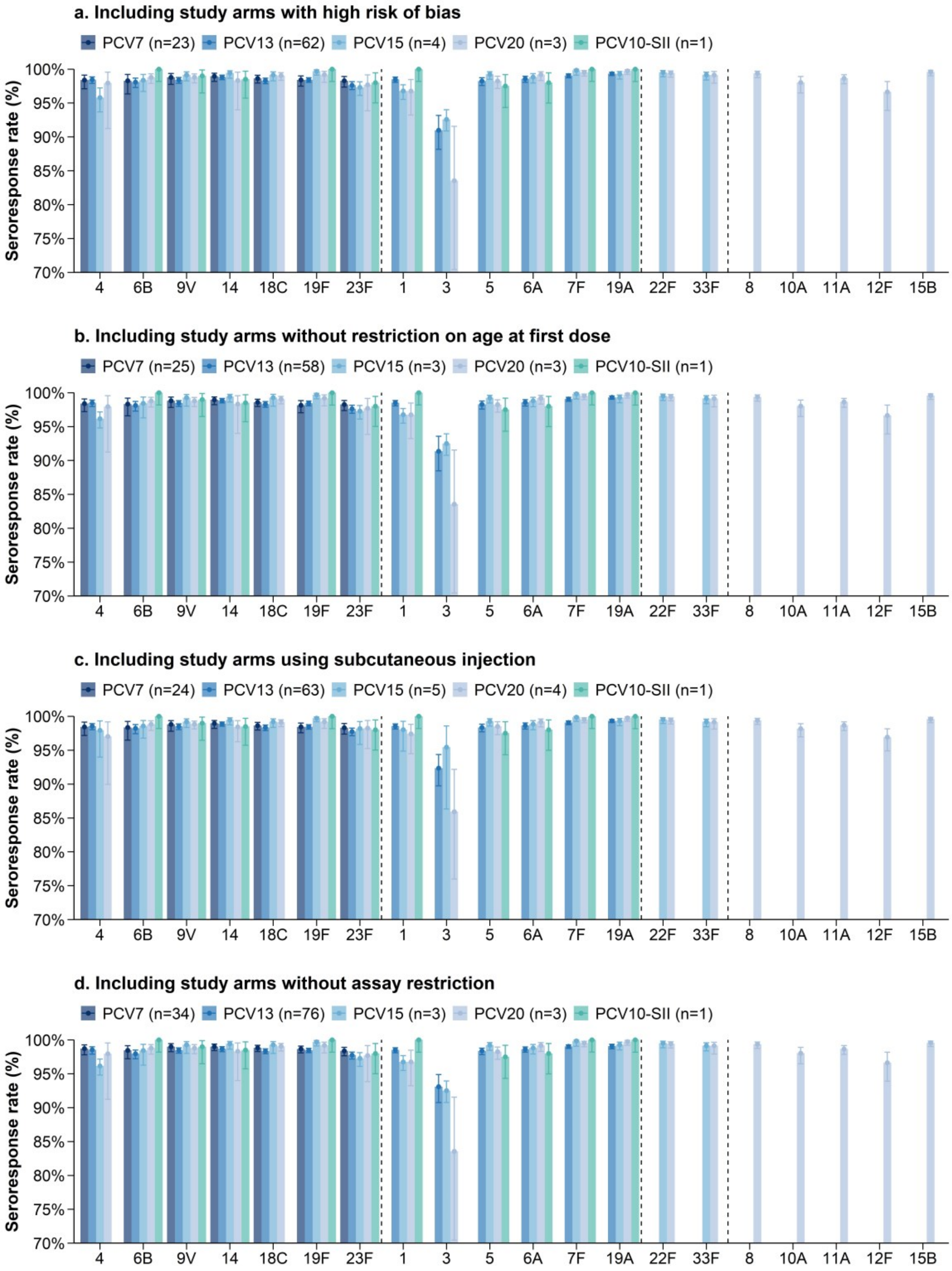

The seroresponse rate indicates the proportion of participants achieving a predefined antibody response threshold. The y-axes in all panels are restricted to the range of 70–100% to highlight differences across vaccines.

### Figure S14. The funnel plot analysis for PCV7

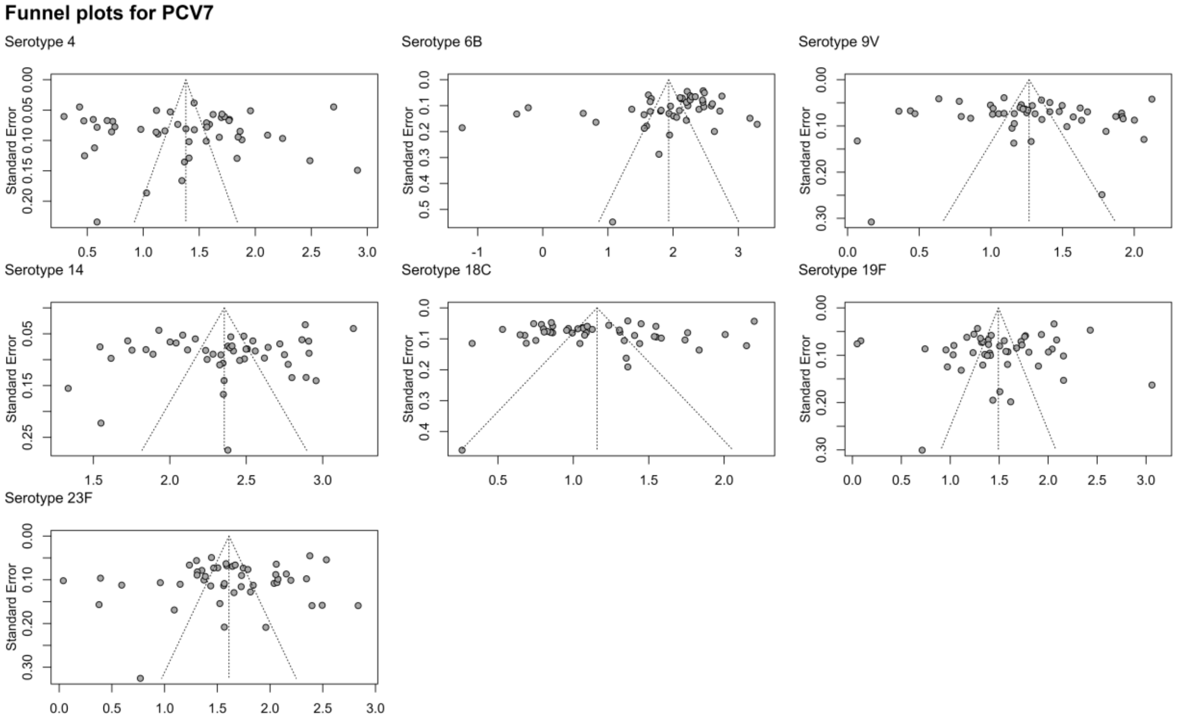

### Figure S15. The funnel plot analysis for PCV13

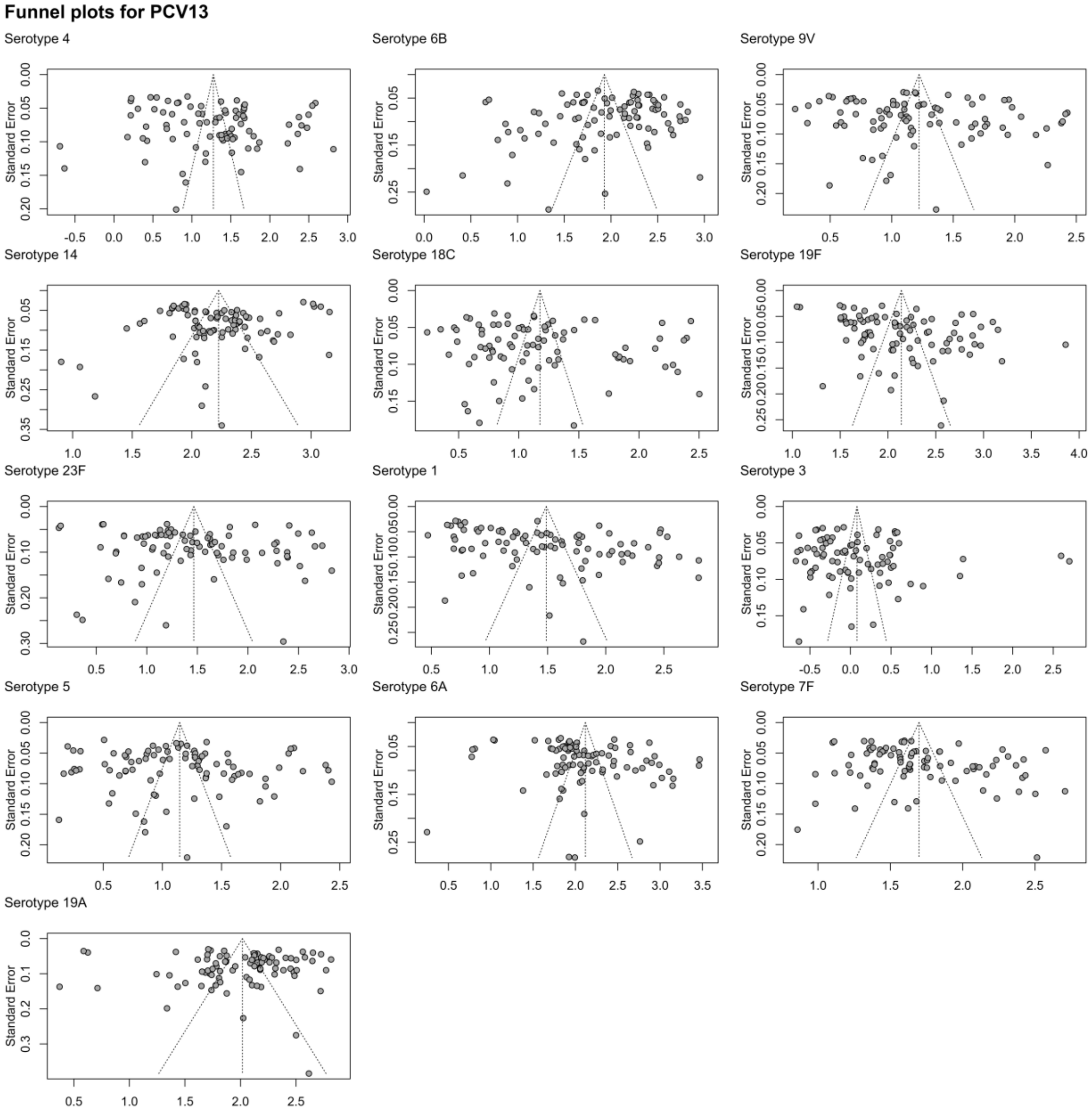

5. World Health Organization. Training manual for enzyme linked immunosorbent assay for the quantitation of streptococcus pneumoniae serotype specific igg (pn ps elisa) (89sf version). 2004.

6. World Health Organization. Training manual for enzyme linked immunosorbent assay for the quantitation of streptococcus pneumoniae serotype specific igg (pn ps elisa) (007sp version). 2011.

22. U.S. Food and Drug Administration. Prevnar. 2018.
